# Does a humoral correlate of protection exist for SARS-CoV-2? A systematic review

**DOI:** 10.1101/2022.01.21.22269667

**Authors:** Julie Perry, Selma Osman, James Wright, Melissa Richard-Greenblatt, Sarah A Buchan, Manish Sadarangani, Shelly Bolotin

## Abstract

**Background:** A correlate of protection (CoP) is an immunological marker associated with protection against infection. A CoP can be used to determine whether an individual is protected from infection, evaluate candidate vaccines, guide vaccination dosing intervals and policy, and understand population-level immunity against a pathogen. Despite an urgent need, a CoP for SARS-CoV-2 is currently undefined, leaving an evidence gap for informing public health policy and adapting it appropriately as new variants of concern emerge. The objective of this study was to systematically review and assess the evidence for a humoral SARS-CoV-2 CoP.

**Methods and Findings:** We searched OVID MEDLINE, EMBASE, Global Health, Biosis Previews and Scopus from inception to January 4, 2022 and pre-prints (using NIH iSearch COVID-19 portfolio) from inception to December 31, 2021, for studies describing SARS-CoV-2 re-infection or breakthrough infection with associated antibody measures. Two reviewers independently extracted study data and performed quality assessment. Twenty-five studies were included in our systematic review. Several studies reported re-infection or breakthrough cases that occurred in the presence of robust antibody levels. Studies that compared aggregate antibody concentrations from individuals who experienced re-infection or breakthrough compared to those who remained protected did not always find differences that were statistically significant. However, several studies found an inverse relationship between antibody levels and infection incidence, risk, or viral load, and a correlation between antibody levels and vaccine efficacy (VE). Estimates of the contribution of antibody levels to VE varied from 48.5% to 94.2%, suggesting that both humoral immunity and other immune components contribute to protection. Only two studies estimated a quantitative CoP. For Ancestral SARS-CoV-2, these included 154 (95% confidence interval (CI) 42, 559) anti-S binding antibody units/mL (BAU/mL), and 28.6% (95% CI 19.2, 29.2%) of the mean convalescent antibody level following infection. One study reported a CoP for the Alpha (B.1.1.7) variant of concern of 171 (95% CI 57, 519) BAU/mL. As of our search date, no studies reported an Omicron-specific CoP.

**Conclusions:** The reviewed literature was limited by a wide variation in assay methodology and antibody targets. Few studies reported SARS-CoV-2 lineage. The studies included in our review suggest that if it exists, a SARS-CoV-2 CoP is likely relative, where higher antibody levels decrease the risk of infection, but do not eliminate it completely. More work is urgently needed in this area to establish a SARS-CoV-2 CoP and guide policy as the pandemic continues.

## Introduction

As the COVID-19 pandemic progresses, our understanding of immunity against SARS-CoV-2 continues to evolve. Both previous infection and vaccination against SARS-CoV-2 appear to provide protection against infection and severe disease (1, 2), but the mechanism and durability of that protection remains unclear (3, 4). Humoral and cellular immunity likely both contribute to protection (5, 6), but it is uncertain whether a correlate of protection (CoP) for SARS-CoV-2 exists, and if so, whether it is easily quantifiable using a diagnostic laboratory test. Without a CoP, serological testing can confirm previous infection or vaccination, but not immunity, leaving an evidence gap in public health policy particularly as new variants of concern emerge.

A CoP is an immunological marker associated with protection from an infectious agent following infection or vaccination (7). Some CoPs are mechanistic, indicating that they are directly responsible for protection. Other CoPs are non-mechanistic or surrogate, and although not directly responsible for protection, can be used in substitute of the true correlate even if it is unknown (8, 9). A CoP can be absolute, where protection against disease is certain above a threshold, or relative, where higher levels of a biomarker correspond to more protection. However, for relative CoPs, even high levels are not protective in some instances (6). Some correlates vary by endpoint (e.g. symptomatic infection or severe disease), or are only applicable to a specific endpoint (9). The majority of CoPs described are humoral and used in a surrogate manner, as these antibodies are easier to detect in clinical laboratory settings than components of cellular immunity (10).

Elucidating a CoP for SARS-CoV-2 is a critical priority for improving our understanding of the extent and duration of protection against infection for individuals and populations. At the individual level, a CoP would provide clear immunological vaccine trial endpoints, and therefore may provide a pathway to licensure for new vaccines (10). If measurable using a diagnostic test, a CoP would enable determination of individual immunity, which is particularly important for immunocompromised individuals (11, 12) and individuals whose immunity levels have waned (13). At the population level, a CoP may enhance the utility of serosurveys, by enabling the assessment the level of protection within a community (10).

The search for a SARS-CoV-2 CoP is further complicated by the emergence of variants of concern (VOCs). Sera from previously infected and/or vaccinated individuals have reduced neutralizing ability against VOCs including Beta (B.1.351), Delta (B.1.617.2) and Omicron (B.1.1.529) (14–16), with the latter showing the greatest extent of immune evasion of all VOCs thus far (17). This complicates the search for a CoP, and raises the possibility that a SARS-CoV-2 CoP may be VOC-specific.

With this in mind, and considering that an easily measurable CoP would most likely be humoral and not cellular, we performed a systematic review to assess the evidence for a humoral CoP for SARS-CoV-2.

## Methods

### Data Sources and Searches

We searched the OVID MEDLINE database for peer-reviewed articles published from database inception to December 31, 2021, and the EMBASE, Global Health, Biosis Previews and Scopus databases from inception to January 4, 2022. We used the NIH iSearch COVID-19 Portfolio tool to search for preprint articles published from database inception to December 31, 2021. In our search strategy, we focused on studies reporting either re-infection or breakthrough infection following vaccination, since both allow an evaluation of humoral immune protection. All search terms used are reported in Supplementary Table 1. We also searched reference lists for suitable articles, and requested article recommendations from experts in the field.

### Study Selection

One reviewer screened titles and abstracts using Distiller SR (Ottawa, Ontario, Canada). Studies passed title and abstract screening if their abstracts discussed re-infection with SARS-CoV-2 or breakthrough infection following vaccination with COVID-19 vaccine; mentioned antibody measures specific to SARS-CoV-2; or mentioned a correlate or threshold of protection against SARS-CoV-2. We excluded studies that focused on immunocompromised populations or animal models.

Two reviewers screened full texts of articles that met title/abstract screening criteria using defined re-infection and breakthrough infection criteria (Table 1). During full-text screening, we included studies reporting a quantitative CoP against SARS-CoV-2, and studies reporting re-infection or breakthrough infection according to our definitions along with associated pre-infection measures for any antibody isotype. If these studies reported aggregate antibody measures (i.e. geometric mean titres (GMT)) we required them to include summary statistics (i.e. statistical significance testing or 95% confidence intervals (95% CI)) to permit the determination of statistically significant differences between groups. We also included studies that correlated antibody levels to vaccine efficacy (VE) or effectiveness, but only if they provided statistical summary measures (e.g. a correlation co-efficient describing the relationship between antibody level and VE), or if they correlated an antibody concentration to a VE of 100% (i.e. absolute protection). We only included studies written English or French. We calculated a Cohen’s Kappa to assess inter-rater agreement for full-text screening. Discrepancies were resolved through discussion or using additional reviewers as needed.

**Table 1:**
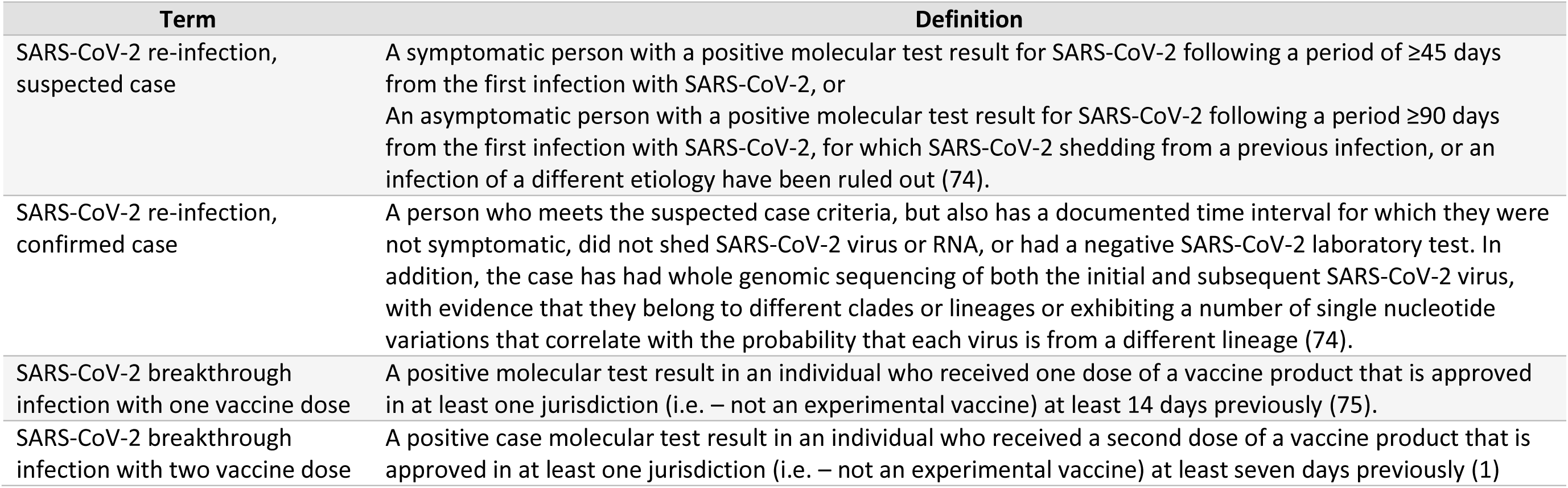
Definitions applied to determine cases of re-infection and breakthrough in this systematic review.

| Term | Definition |
| --- | --- |
| SARS-CoV-2 re-infection, suspected case | A symptomatic person with a positive molecular test result for SARS-CoV-2 following a period of $\geq 45$ days from the first infection with SARS-CoV-2, or<br>An asymptomatic person with a positive molecular test result for SARS-CoV-2 following a period $\geq 90$ days from the first infection with SARS-CoV-2, for which SARS-CoV-2 shedding from a previous infection, or an infection of a different etiology have been ruled out (74). |
| SARS-CoV-2 re-infection, confirmed case | A person who meets the suspected case criteria, but also has a documented time interval for which they were not symptomatic, did not shed SARS-CoV-2 virus or RNA, or had a negative SARS-CoV-2 laboratory test. In addition, the case has had whole genomic sequencing of both the initial and subsequent SARS-CoV-2 virus, with evidence that they belong to different clades or lineages or exhibiting a number of single nucleotide variations that correlate with the probability that each virus is from a different lineage (74). |
| SARS-CoV-2 breakthrough infection with one vaccine dose | A positive molecular test result in an individual who received one dose of a vaccine product that is approved in at least one jurisdiction (i.e. – not an experimental vaccine) at least 14 days previously (75). |
| SARS-CoV-2 breakthrough infection with two vaccine dose | A positive case molecular test result in an individual who received a second dose of a vaccine product that is approved in at least one jurisdiction (i.e. – not an experimental vaccine) at least seven days previously (1) |

### Data extraction and Quality Assessment

Two reviewers extracted data in duplicate from articles that met full-text screening criteria. We extracted data from figures using WebPlotDigitizer (18). We summarized and synthesized the data, stratifying the included studies by whether they described re-infection or breakthrough infection. We explored the possibility of meta-analyzing our results.

We used the National Institutes of Health National Heart, Lung and Blood Institute (NIH NHLBI) Study Quality Assessment tools to assess the quality of each study using the corresponding tools specific for each study design (19), and adapted it by adding questions to customize the tool for this study. Studies correlating VE to antibody levels were evaluated using the Cohort and Cross Sectional Tool.

### Data Synthesis and Analysis

We reported our results using the Preferred Reporting Items for Systematic Reviews and Meta-Analyses (PRISMA) 2020 (20). Recognizing that that the immune response following natural infection and vaccination may differ, we grouped studies involving re-infection separately from studies examining breakthrough infection for analysis.

## Results

Our literature search identified 11,803 records for screening (Figure 1). After de-duplication, we screened 4,919 peer-reviewed studies, 783 preprint studies and 16 studies identified through expert recommendations and scanning of article reference lists. After title/abstract screening and full-text screening, for which our Kappa was 1.0, we included 30 articles in our review. However, only 25 articles passed quality assessment. Of these, 14 described SARS-CoV-2 re-infection along with individual or aggregate humoral measures (2, 21–33), and 11 studies described SARS-CoV-2 breakthrough infection following vaccination or statistical modelling to explore associations between VE and antibody levels (34–44) (Table 2). Only two studies estimated a SARS-CoV-2 antibody CoP, both using statistical modelling methods (38, 39).

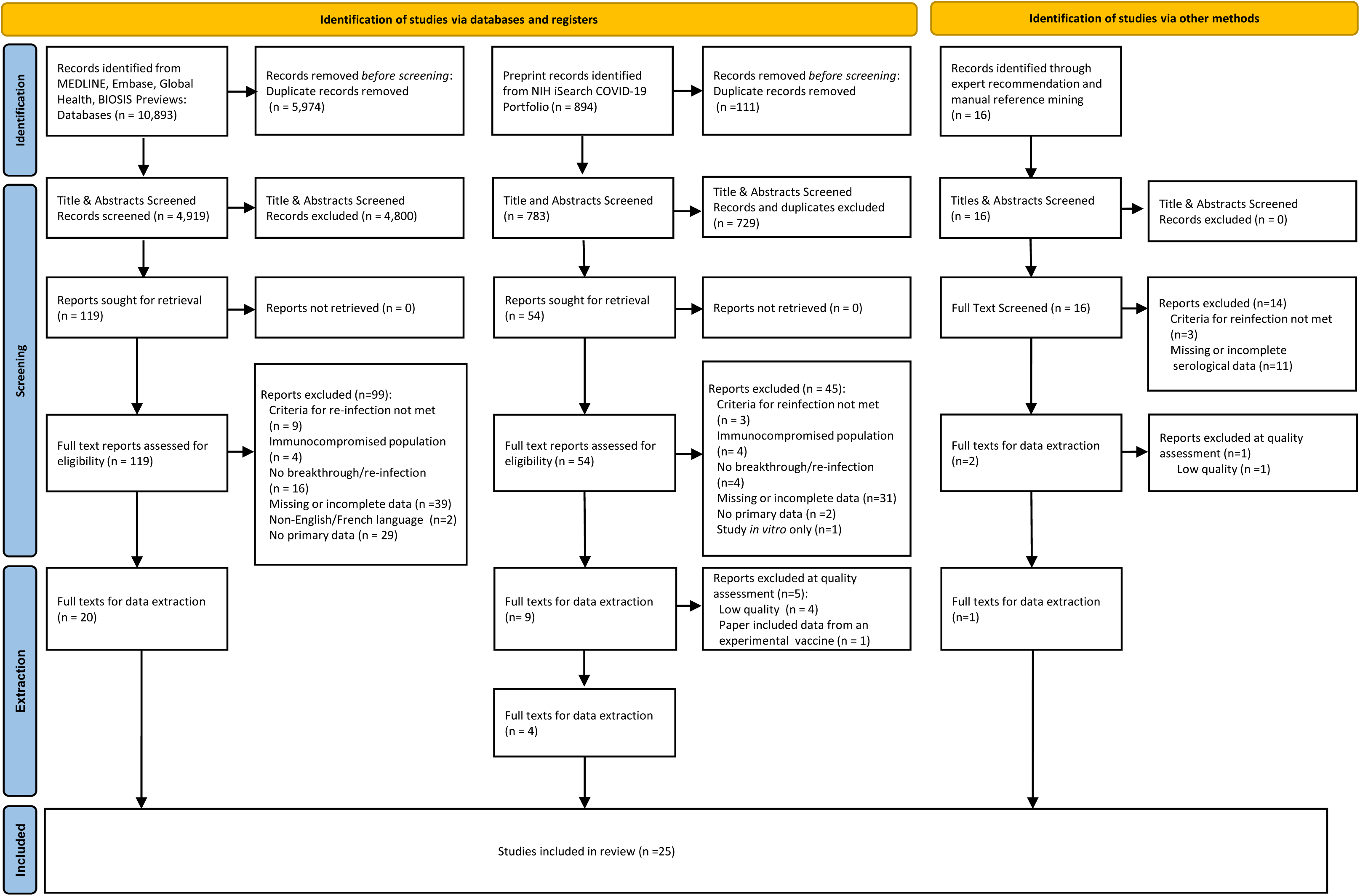

**Table 2:**
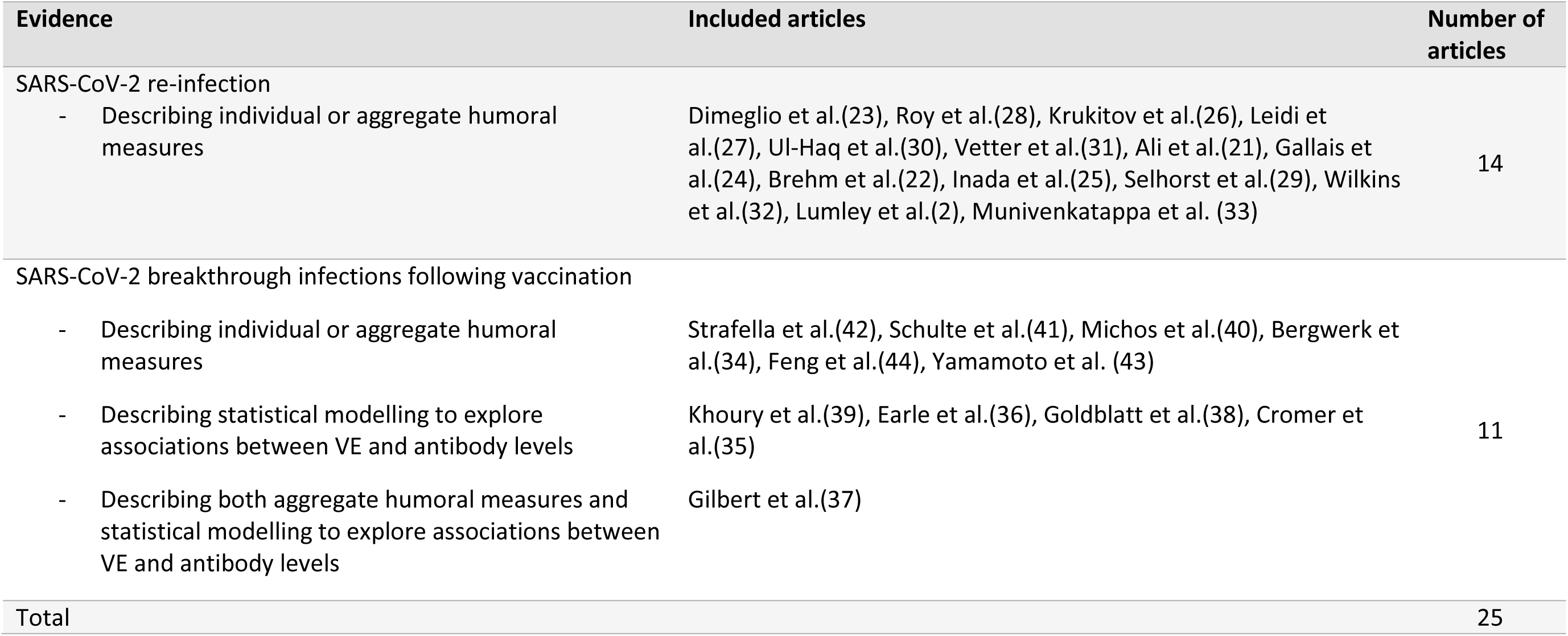
Summary of articles included in this review following re-infection and breakthrough infection definition screening, and types of evidence they describe.

### Studies describing SARS-CoV-2 re-infection

Fourteen studies met our SARS-CoV-2 re-infection definition and provided pre-infection antibody values (Table 3). These included seven cohort studies (2, 21, 23, 24, 26, 27, 32), and seven case reports (22, 25, 28–31, 33). Most study populations were healthcare workers, patients, or long term care home residents (2, 21–24, 26, 29–33). The remaining studies described individuals from the general population (25, 27, 28). Although not always reported, specimen collection occurred between 14 days and seven months after initial infection (22, 31) and between 4 days and seven months before re-infection (26, 32). Antibody test results included various commercial and laboratory developed enzyme-linked immunosorbent assays (ELISAs) targeting anti-spike (anti-S), anti-receptor binding domain (anti-RBD) and anti-nucleocapsid (anti-N) antibodies, as well as neutralization assays. No study utilized the World Health Organization (WHO) International Standard (IS), which was developed to enable the comparison of serological data from different platforms (45). Only three papers reported on the SARS-CoV-2 lineage of the re-infection (22, 29, 31). No studies reported serological measures preceding re-infection with VOCs.

**Table 3:** Articles describing SARS-CoV-2 re-infection along with individual or aggregate humoral measures^#^

| First author, publication year (study country) | Study design, population | Number of reinfections reported | Lineage of first infection, reinfection | Time from first infection to most recent antibody test before re-infection* (days) | Antibody assay, target isotype (cut-off) | Pre reinfection antibody level* | Time from most recent antibody test* to re-infection (days) | Statistical association |
| --- | --- | --- | --- | --- | --- | --- | --- | --- |
| **Inada, 2020 (Japan)** | Case report, general public | 1 | Not provided | 94 | Laboratory developed Anti-S IgG ELISA (cut-off not provided) | 15.6 OD ratio | 11 | None reported |
|  |  |  |  | 94 | Laboratory developed neutralization assay, IgG specific | 50 µg/mL | 11 | None reported |
| **Roy, 2021 (Not Reported)** | Case report, general public | 1 | Not provided | 150 (5 months) | LIASON SARS-CoV-2 S1/S2 IgG test kit (DiaSorin Inc., Saluggia, Italy) (>15.0) | 48 AU/ml | 47 | None reported |
| **Dimeglio, 2021 (France)** | Cohort, HCW | 5 | Not provided | Not provided | Quantitative ELISA (Wantai Biological Pharmacy Enterprise Co, Ltd, China); Total Ab; anti-Spike | Range: 1.5-385.8 S/Co | Not provided (serology performed a median of 167 IQR (156–172) days apart) | None reported |
|  |  |  |  | Not provided | Neutralization test – assay not provided | Range: 0-64 S/CO | Not provided (serology performed a median of 167 days apart) | None reported |
| **Leidi, 2021**<br>**(Switzerland)** | Cohort,<br>general<br>public | 5 | Not<br>provided | Not provided | Euroimmun ELISA,<br>(Euroimmun Lubeck,<br>Germany); IgG; anti-S (cut-off: $\geq 0.5$) | Range: 0.58-2<br>ratio | Range: 34-185 | None reported |
| **Lumley, 2021**<br>**(England)** | Cohort,<br>HCW | 3 | Not<br>provided | 50-112 days<br>for HCW2; Not<br>provided for<br>HCW1 and<br>HCW3 | ELISA (LDT); IgG; Anti-S (cut-off not provided) | Range: 0.34-10.5 million<br>units | Range: 61-179 | IRR of 0.11 (95%<br>CI 0.03, 0.44, p<br>= 0.002) in<br>seropositive<br>healthcare<br>workers<br>compared to<br>seronegative<br>healthcare<br>workers |
|  |  |  |  | 50-112 days<br>for HCW2; Not<br>provided for<br>HCW1 and<br>HCW3 | ELISA (LDT); IgG; Anti-N (cut-off not provided) | Range: 0-7.5<br>arbitrary units | Range: 10-179 | IRR of 0.11 (95%<br>CI 0.03, 0.45, p<br>= 0.002) in<br>seropositive<br>healthcare<br>workers<br>compared to<br>seronegative<br>healthcare<br>workers |
| **Ul-Haq, 2020**<br>**(Pakistan)** | Case<br>report,<br>HCW | 1 | Not<br>provided | 15 | Assay information not<br>provided, cut off of $\geq 1$ | 1.97 | 133 | None reported |
| **Vetter, 2021**<br>**(Switzerland)** | Case<br>report,<br>HCW | 1 | Re-infection<br>lineage<br>different<br>than first<br>infection, | 35 | Euroimmun Anti-S IgG<br>(Euroimmun, Lubeck,<br>Germany) (cut-off not<br>provided) | 2.16 UI/l | 169 | None reported |
|  |  |  |  | 35 | Elecsys/Roche (Basel,<br>Switzerland), Total anti-RBD<br>(0.8 U/ml) | 21.6 U/ml | 169 |  |
|  |  |  | but both<br>clade 20A | 35 | Elecsys/Roche (Basel,<br>Switzerland), Total anti-N<br>(cut-off not provided) | 128 COI | 169 |  |
|  |  |  |  | 35 | PRNT/neutralization assay<br>90% | 14.1 (1/<br>(inferred to<br>mean 1/14.1) | 169 |  |
| **Ali, 2020 (Iraq)** | Cohort,<br>patients<br>admitted<br>to hospital | 17** | Not<br>provided | Not provided | IgG Anti-N (PishTaz Teb<br>Diagnostic, Tehran, Iran)<br>(cut-off=1.1) | 5.87 (s/ca) | Not provided | None reported |
| **Gallais, 2021**<br>**(France)** | Cohort,<br>HCW | 1 | Not<br>provided | 96 | Abbott Architect SARS-CoV-2 IgG Quant II assay (Abbott,<br>Sligo, Ireland)<br>(cut-off:50AU/ml) | 2.6 log AU/ml | 7 months<br>(number of<br>days not<br>reported) | None reported |
|  |  |  |  | 96 | EDI Novel coronavirus<br>COVID-19 IgG ELISA (San<br>Diego, USA) (no cut-off<br>reported) | 1.0 OD S/CO | 7 months<br>(number of<br>days not<br>reported) |  |
| **Brehm, 2021**<br>**(Germany)** | Case<br>report,<br>HCW | 1 | B.3,<br>B.1.177 | ~6 months | Diasorin IgG Anti-S<br>(Saluggia, Italy) (cut-off: 15<br>AU/mL) | 60 AU/mL | ~4 months<br>(number of<br>days not<br>reported) | None reported |
|  |  |  |  | 210 | Indirect<br>immunofluorescence, IgG,<br>IgM, IgA | IgG 1:320<br>IgM <1:20<br>IgA <1:20 | 73 |  |
|  |  |  |  | 210 | Neutralization Assay | Local Hamburg<br>reference<br>isolate (HH-1):<br>1:80 IC50<br>B.1.177: 1:160<br>IC50 | 73 |  |
| **Selhorst, 2020**<br>**(Belgium)** | Case<br>report,<br>HCW | 1 | V clade, G<br>clade | 105 | Roche Total anti-N (Basel,<br>Switzerland) (cut-off: ≥1) | 102 cut-off/<br>index | 80 | None reported |
|  |  |  |  | 94 | PRNT/neutralization assay;<br>2019-nCoV-Italy-INMI1;<br>NT50 | NT<sub>50</sub> 200 | 91 |  |
| **Munivenkatappa, 2021 (India)** | Case report, HCW | 1 | Not provided | 76 days | ELISA (LDT), IgG, anti-RBD (no cut-off provided) | Ratio of positive to negative: 4.14 | 31 days | None reported |
|  |  |  |  | 76 days | ELISA (LDT), IgG, anti-N (no cut-off provided) | Ratio of positive to negative: 8.57 | 31 days | None reported |
|  |  |  |  | 76 days | PRNT/Neutralization assay, no details provided | Positive (no quantitative result given) | 31 days |  |
| **Krutikov, 2021 (England)** | Cohort, staff and residents in LTC | 14 | Not provided | Not provided | Mesoscale Diagnostics (MSD) IgG, anti-S (Rockville, USA) (no cut-off provided) | Range: 78-137840 AU/mL | Range: 12-132 | Cox regression showed antibody-negative staff and residents at baseline had increased risk of PCR+ infection than those antibody-positive at baseline (aHR range: 0.08 (95% CI 0.03, 0.23) -0.39 (95% CI 0.19, 0.82)) |
|  |  |  |  | Not provided | Mesoscale Diagnostics (MSD) IgG, anti-N (Rockville, USA) (no cut-off provided) | Range: 137–222308 AU/ml;<br>Median antibody levels of 101527 (95% CI 18393, 161580) AU/mL for cases, and<br>26326 (95% CI<br>14378, 59633)<br>AU/mL for<br>controls. | Range: 12-132 | No statistically significant difference between antibody levels of individuals re-infected and those not<br>(p=0.544) |
| **Wilkins, 2021**<br>**(USA)** | Cohort<br>study,<br>HCW | 8 | Not<br>provided | Not provided | Abbott ARCHITECT i2000SR<br>Immunoassay system, IgG,<br>anti-N (Sligo, Ireland) (cut-off: $\geq 1.4$) | Range: 1.92-6.01 Index<br>Value | Range: 95-212 | None reported |

Two studies compared antibody levels between individuals who were re-infected and those who were not. Krutikov et al. did not find a statistically significant difference in anti-N IgG titres (reported as the log_10_ IgG (AU/ml)) between those who were re-infected compared to those who were not (p=0.544) but did show that individuals who were antibody-negative at the start of the study were at greater risk of infection during the study period than those who were antibody-positive (26). Lumley and colleagues used Poisson regression to compare the incidence rate of infection between seropositive and seronegative individuals (2), and found that individuals who were anti-S positive were less likely to be infected compared to those who were anti-S negative (incidence rate ratio (IRR) of 0.11 (95% CI 0.03, 0.44)). Similar findings were observed using anti-N antibody (IRR = 0.11 (95% CI 0.03, 0.45)). Analysis of the association between continuous antibody concentrations and incidence was also statistically significant for both antibodies (p<0.001) (2).

### Studies reporting antibody measures related to breakthrough infection or VE

We included 11 studies describing breakthrough SARS-CoV-2 infection. These included two case reports (41, 42), one cohort study (40), two case-control studies (34, 43), and two studies that re-analyzed antibody data from a clinical trial (37, 44). Five in silico studies utilized statistical methods to explore the association between antibody levels and VE (35–39). The populations studied were either clinical trials or other vaccine study participants (35–39, 44) or healthcare workers (34, 40–43). Three studies reported results in WHO IS units (binding antibody units (BAU)/mL) (37, 38, 42), while the rest used units that were not comparable to each other.

Of the 11 studies describing breakthrough infection, six studies provided individual or aggregate humoral measures (34, 40–44), four studies used statistical modelling to explore associations between VE and antibody levels (35, 36, 38, 39), and one study included both humoral measures and statistical modelling (37) (Tables 4 and 5). Five studies (34, 41–44) reported the lineage of the breakthrough infection, and two modeling studies include VOCs in their analysis (35, 38).

**Table 4:**
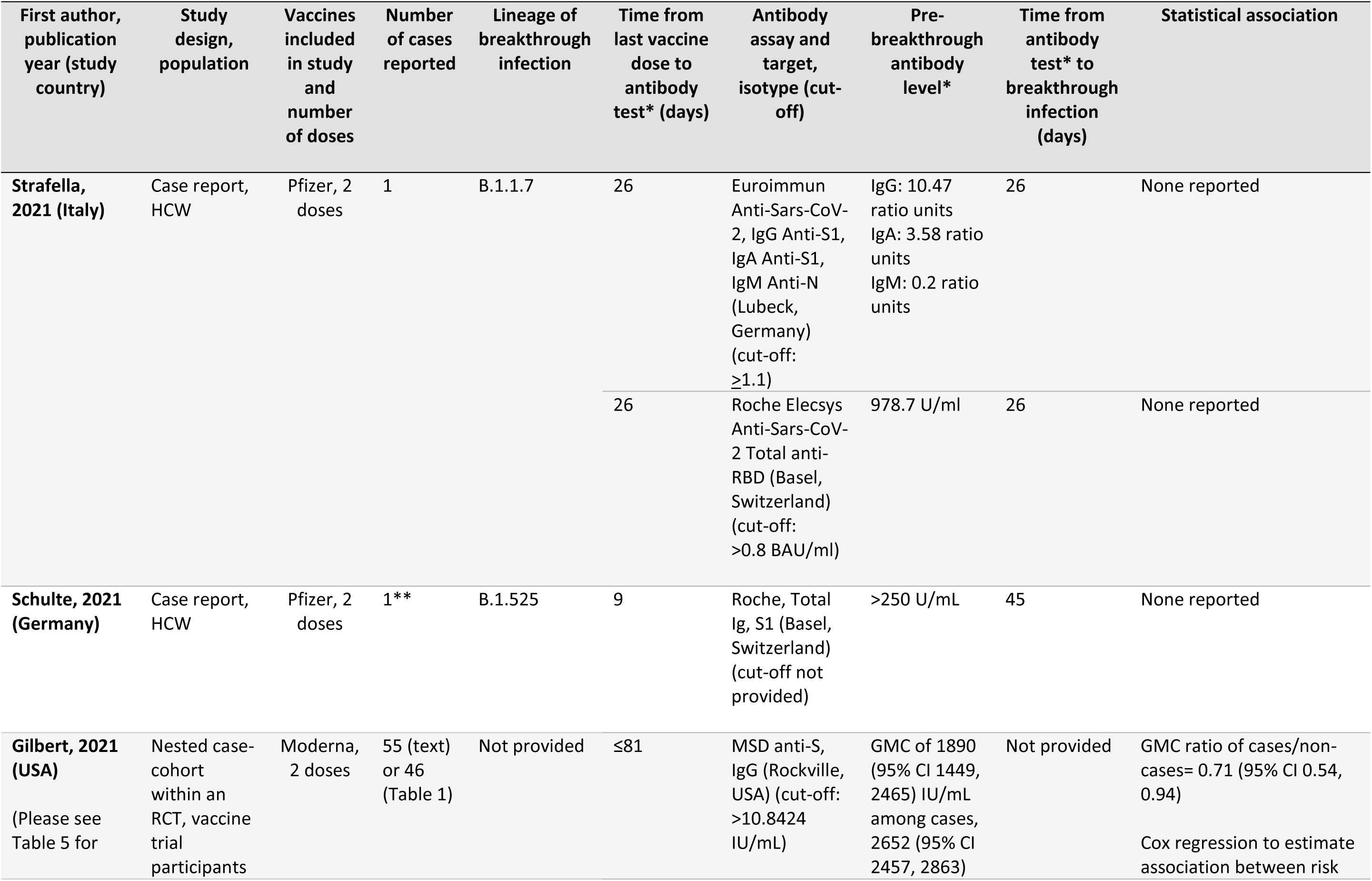
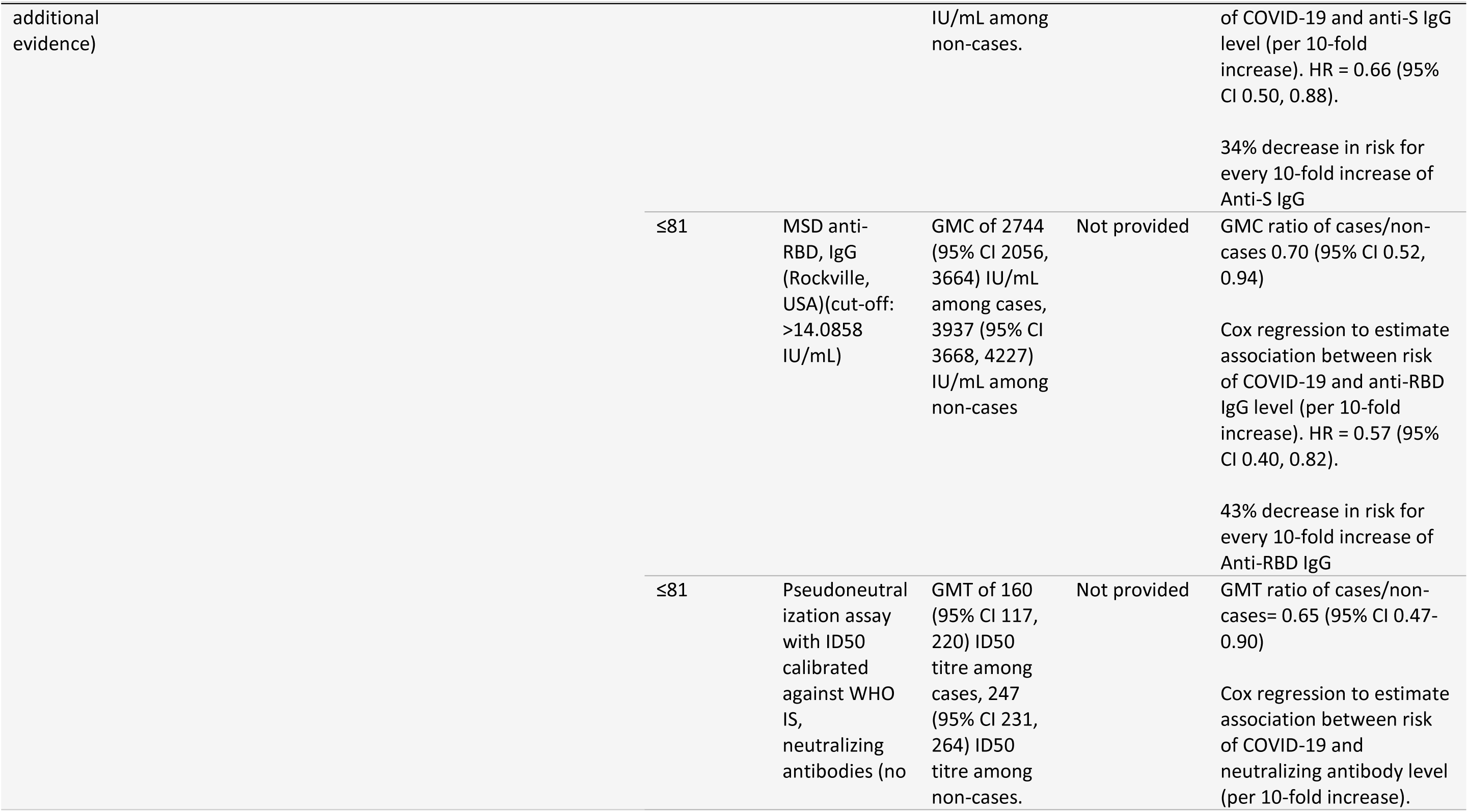
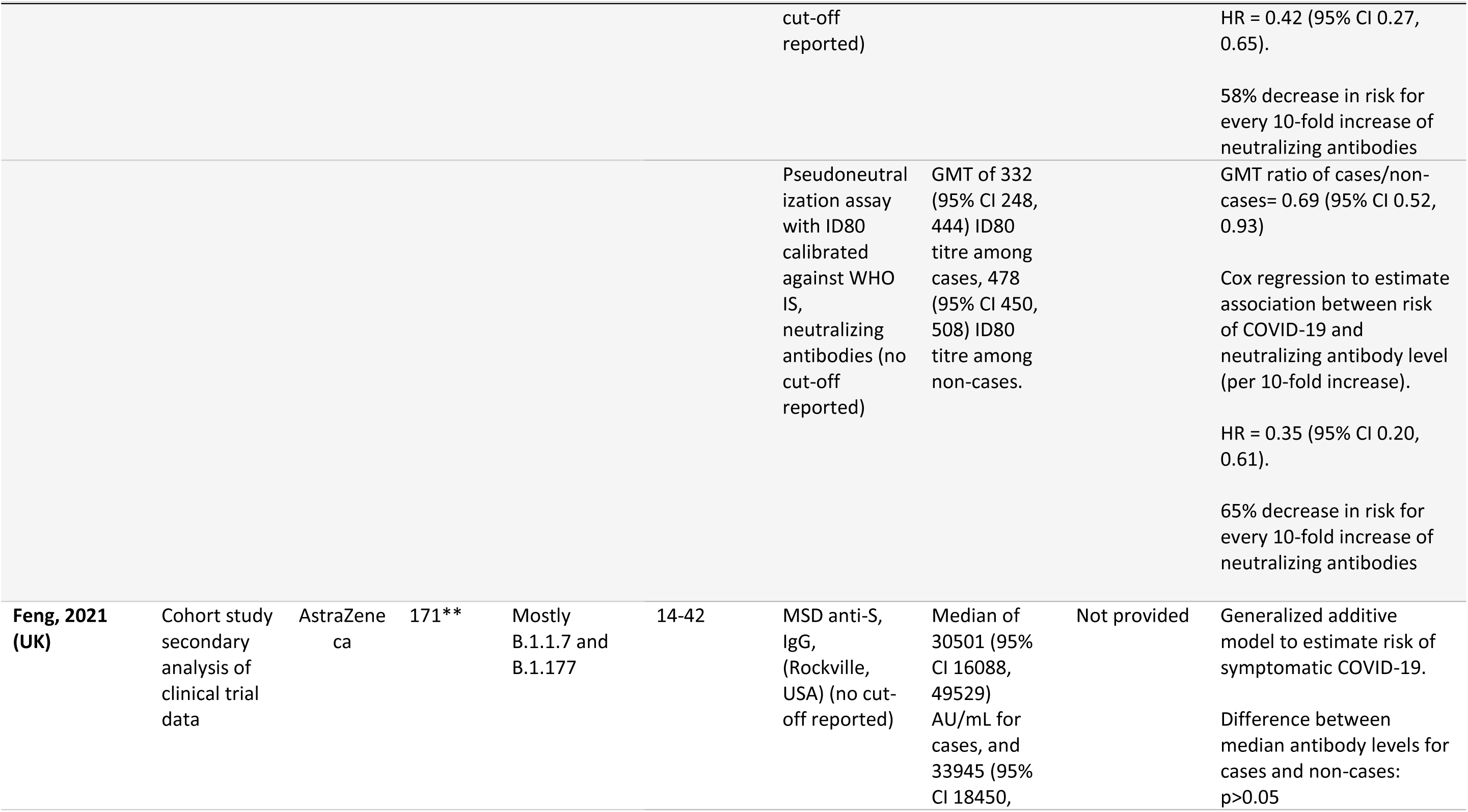
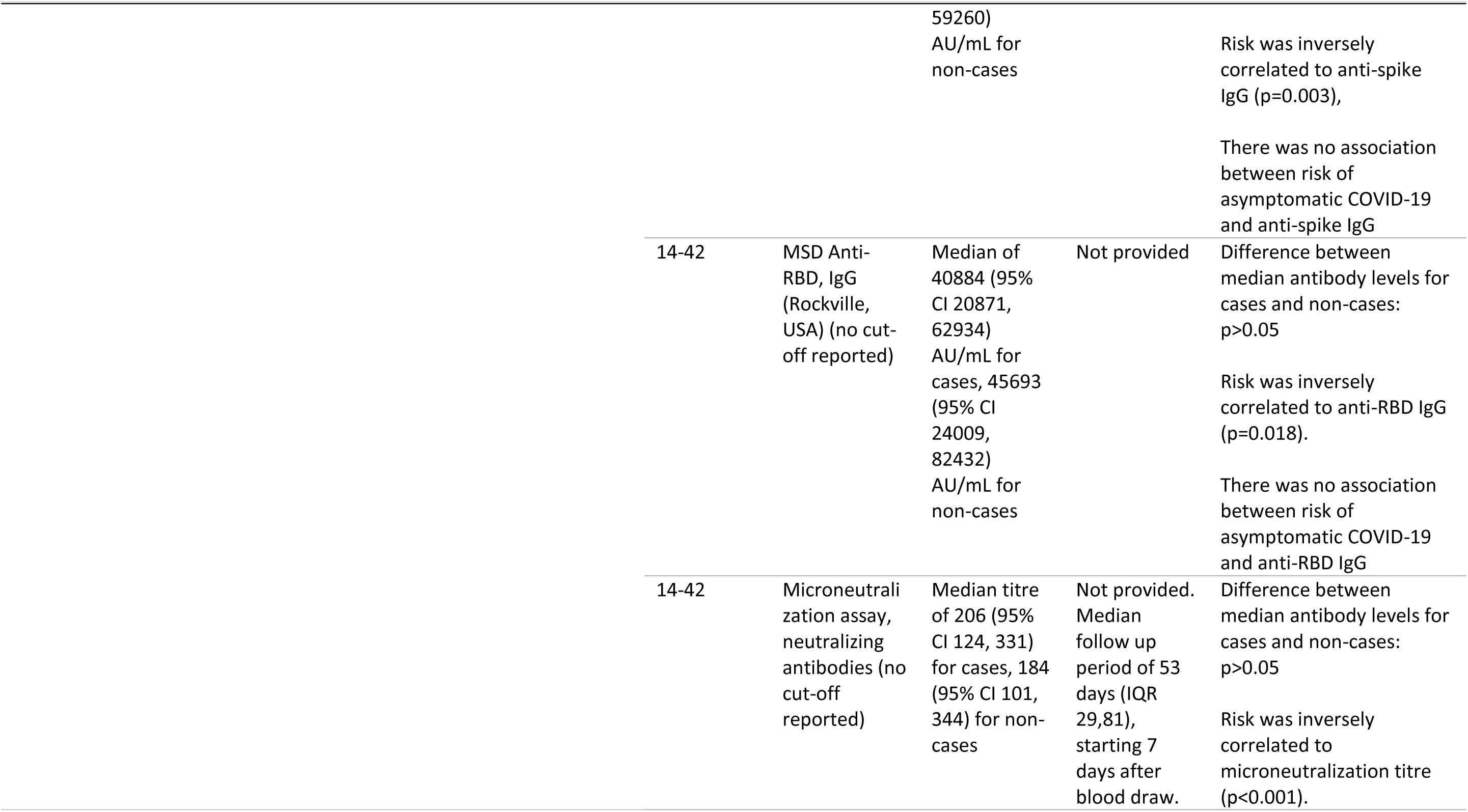
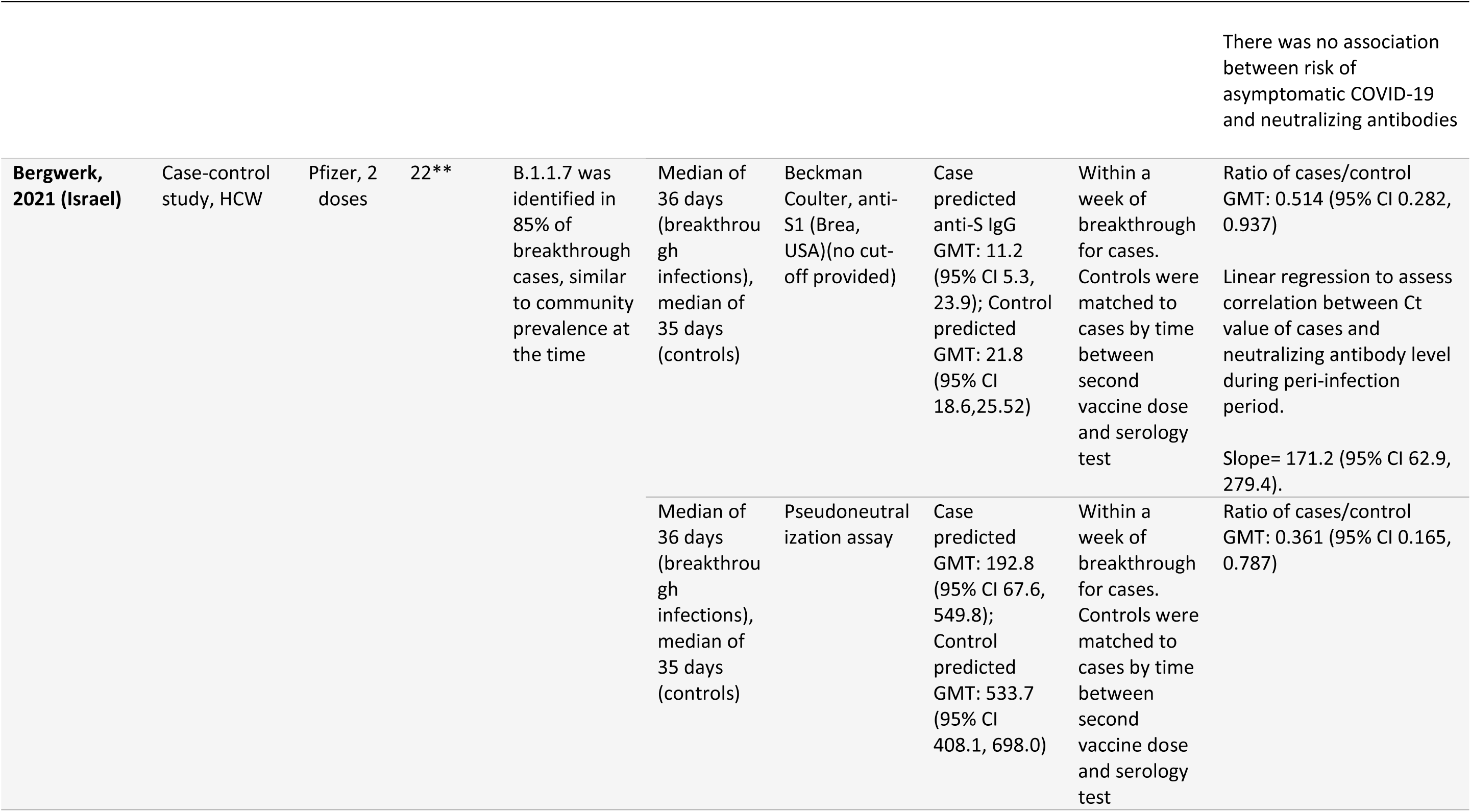
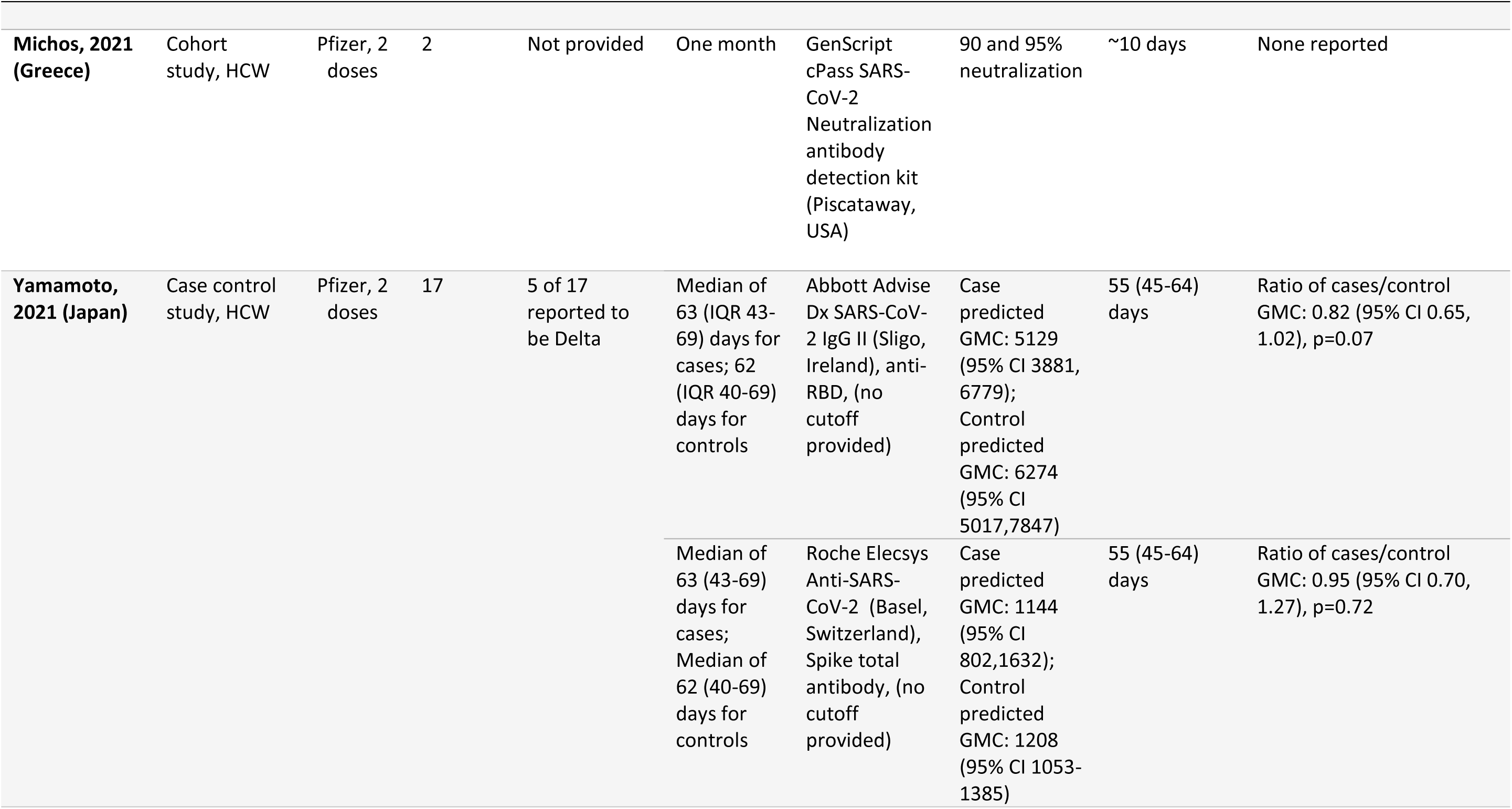
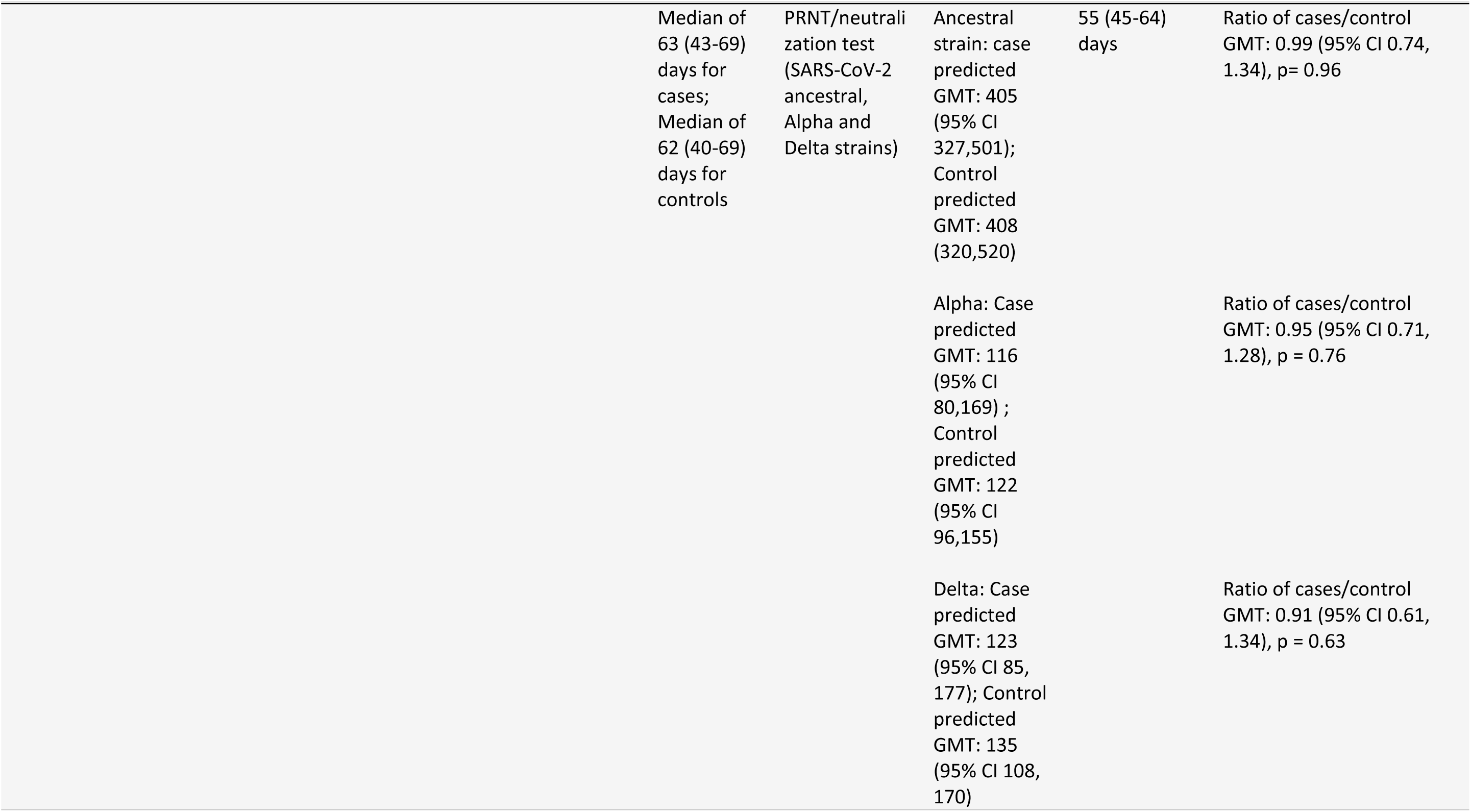
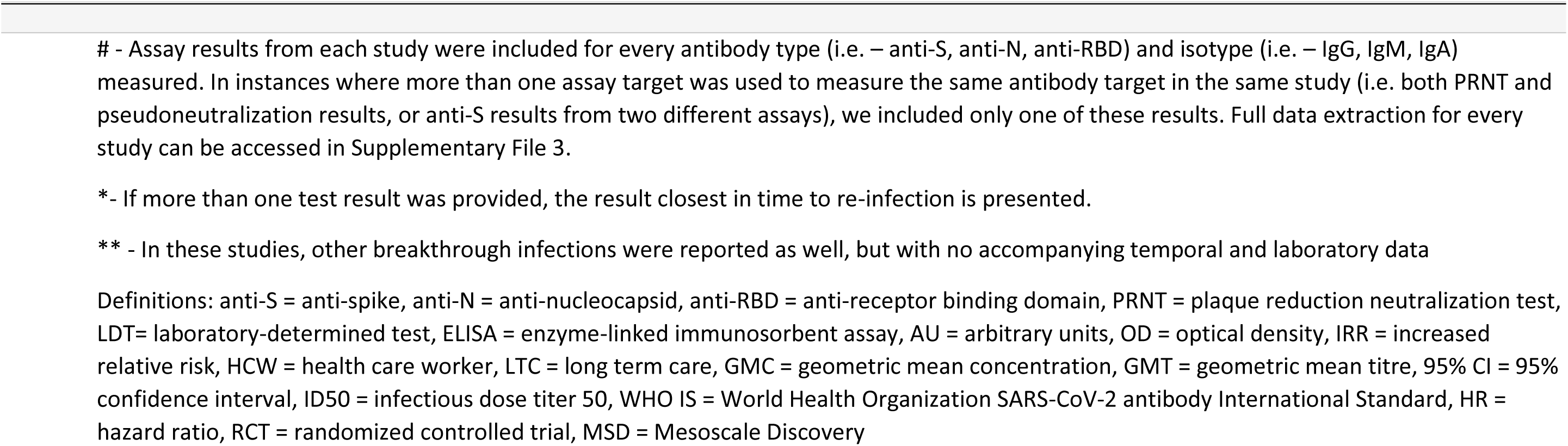
Articles describing breakthrough following SARS-CoV-2 infection along with individual or aggregate humoral measures^#^

**Table 5:**
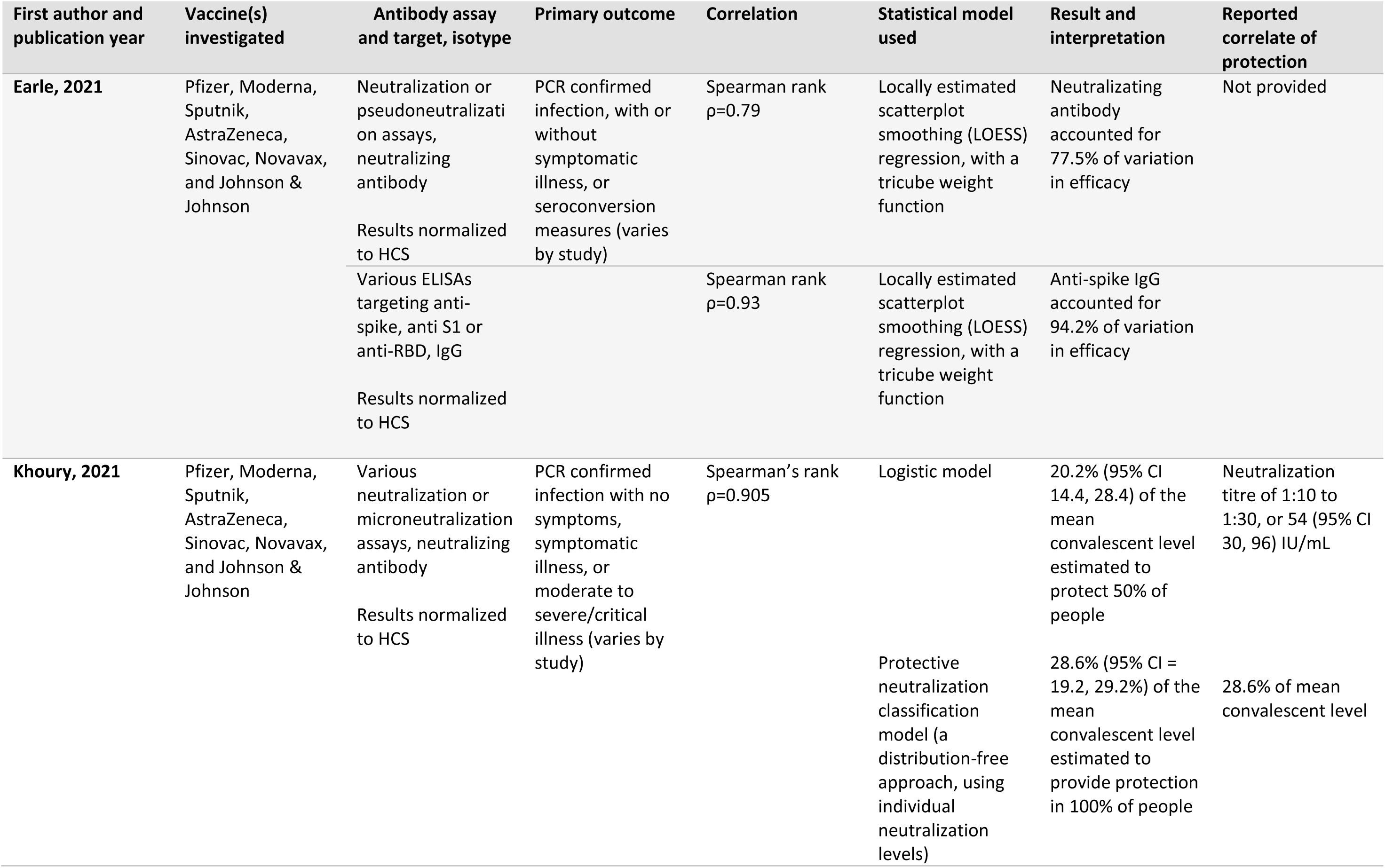
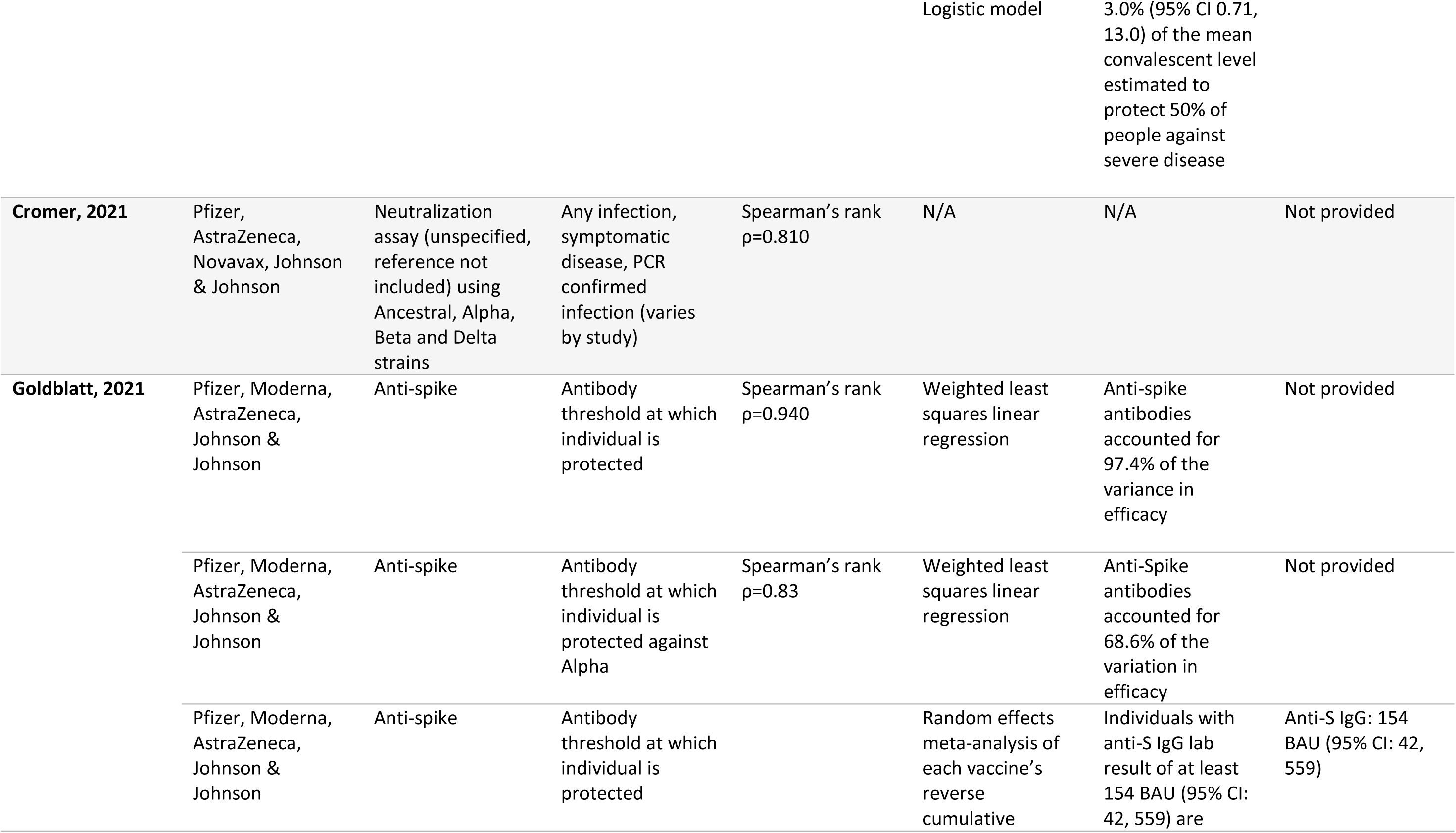
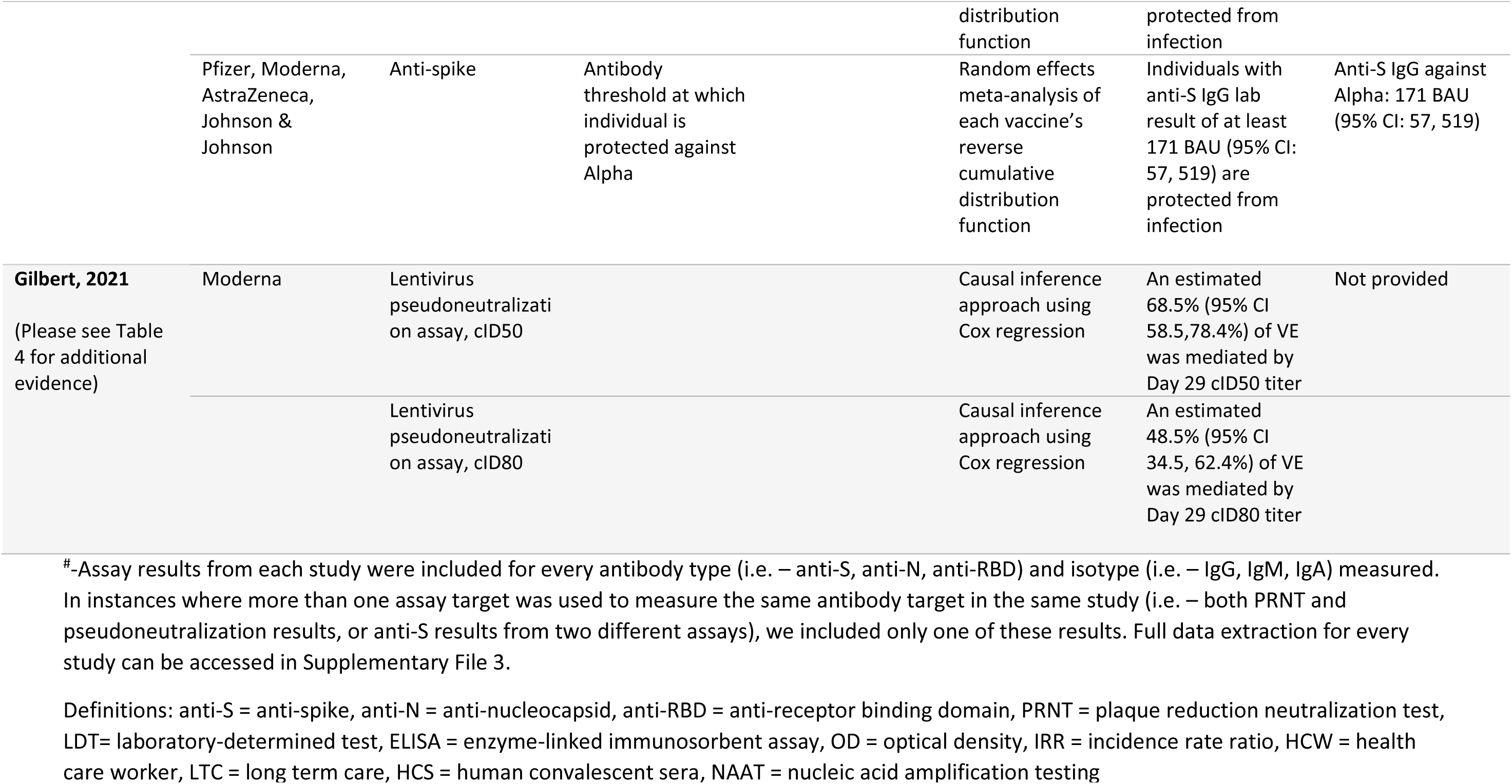
Articles describing statistical modelling to explore associations between VE and antibody levels^#^

#### Studies describing breakthrough infections following SARS-CoV-2 vaccination

Seven of the 11 studies provided individual or aggregate antibody levels following one (40) or two doses of COVID-19 vaccine, including BNT162b2 (Pfizer-BioNTech) (34, 40–43) mRNA-1273 (Moderna) (37) and ChAdOx1 nCoV-19 (AstraZeneca) (44) (Table 4). Depending on the study, specimens were collected between nine (41) and 109 days (37) after administration of the second vaccine dose. Antibody levels were assessed using a variety of commercial serology assays and/or neutralization assays. The time interval between specimen collection for pre-breakthrough antibody levels and breakthrough infection was not always reported. Five studies reported the viral lineage responsible for breakthrough or re-infection, including three studies reporting Alpha (B.1.1.7) (34, 42, 44), one reporting B.1.525 (41), and one reporting Delta (B.1.617.2) (43).

Four of the six studies compared aggregate antibody levels between cases and non-cases. Gilbert et al. calculated geometric mean concentration (GMC) ratios of cases to non-cases, which ranged from 0.57 (95% CI 0.39, 0.84) to 0.71 (95% CI 0.54, 0.94), depending on antibody target and sampling interval (37). Using Cox regression, the authors found statistically significant associations between increasing antibody levels and decreasing risk of COVID-19. Bergwerk et al. applied generalizing estimating equations to predict antibody levels and generate GMT ratios of cases to non-cases. For neutralizing antibodies, these ranged from a case-to-control ratio of 0.15 (95% CI, 0.04, 0.55) at peak values (within the first month after the second vaccine dose) to case-to-control ratio of 0.36 (95% CI 0.17, 0.79) by the week before breakthrough infection (34). Using linear regression, this study demonstrated a statistically significant correlation between cycle threshold (Ct) value of cases and neutralizing antibody level, suggesting an inverse relationship between antibody level and viral load. Feng and colleagues did not find a statistically significant difference between median antibody levels of cases and non-cases, regardless of the antibody assay used (44). However, using a generalized additive model, infection risk was found to be inversely correlated to antibody levels. This result was statistically significant for symptomatic but not asymptomatic COVID-19. Yamamoto et al. found no statistically significant difference in post-vaccination neutralization levels between healthcare workers who experienced a breakthrough infection and matched controls during the Delta wave in Japan (43). The authors found that neutralizing titres were lower against Alpha and Delta variants than the wild-type virus, but were comparable between cases and controls.

#### Studies reporting associations between antibody levels and VE

Five of the 10 breakthrough studies described correlations between antibody levels and VE against BNT162b2 (35, 36, 38, 39), mRNA-1273 (36–39), ChAdOx1 nCoV-19 (35, 36, 38, 39), Ad26.COV2.S (Janssen/ Johnson and Johnson) (35, 36, 38, 39), NVX-CoV2373 (Novavax) (35, 36, 39), CoronaVac (SinoVac) (36, 39), and rAd26+S+rAd5-S (Gamaleya Research Institute) (36, 39) vaccine. These studies re-analyzed clinical trial and other vaccine studies, and as such the VE outcomes of interest varied across the severity spectrum, ranging from asymptomatic PCR confirmed infection to severe disease. The studies generated correlations using either neutralizing antibody levels, derived through plaque reduction neutralization tests (PRNT) or microneutralization assays, or IgG levels measured through ELISAs.

Three of five studies (35, 36, 39) reported correlation coefficients for the relationship between neutralizing antibodies and VE ranging from 0.79 to 0.96. Two studies (36, 38) reported correlation coefficients of 0.82 to 0.94 to describe the relationship between anti-Spike IgG and VE. Since serology and neutralization assays were not calibrated to a common standard, three studies (35, 36, 39) normalized antibody concentrations against convalescent sera used in their respective clinical trials, and reported antibody concentrations as a ratio of the antibody concentration/convalescent serum concentration. The remaining two studies (37, 38) provided results using the WHO IS.

Using different statistical methods, three studies (36–38) attempted to quantitate the contribution of antibodies to VE measures. Earle et al. incorporated data from seven vaccine clinical trials and reported that neutralizing antibodies accounted for 77.5% to 84.4% of VE (36). Gilbert et al. focused on mRNA-1273 clinical trial data and reported that neutralizing antibodies accounted for 48.5% (95% CI 34.5, 62.4%) to 68.5% (95% CI 58.5, 78.4%) of VE (37). This approach was also taken to estimate the effect of anti-S antibodies, with Earle and colleagues finding that anti-S antibody accounts for 91.3% to 94.2% (no CIs provided) of variation in efficacy (36). Goldblatt et al., using data from a convenience sample of individuals vaccinated with BNT162b2, mRNA-1273, ChAdOx1 nCoV-19 or Ad26.COV2.S, reported that anti-S antibodies account for 68.6% to 97.4% (no CIs provided) of variation in efficacy (38).

Two studies estimated a SARS-CoV-2 threshold of protection. Goldblatt et al. calculated protective thresholds in WHO IS units for ancestral strain SARS-CoV-2 and Alpha (B.1.1.7) of 154 (95% CI 42, 559) and 171 (95% CI 57, 519) anti-S binding antibody units (BAU/mL), respectively. These were generated using a random effects meta-analytic approach using BNT162b2, mRNA-1273, ChAdOx1 nCoV-19 or Ad26.COV2.S clinical trial data. The analyses also included reverse cumulative distribution functions to estimate vaccine-specific thresholds of protection. Since thresholds calculated from two doses of mRNA vaccine were extremely high and did not overlap with other calculated thresholds, the authors also generated an anti-S threshold that excluded them (60 (95% CI 35, 102) BAU/mL). Khoury and colleagues used a protective neutralization classification model to estimate the antibody concentration resulting in 100% protection, which they estimated to be 28.6% (95% CI 19.2–29.2%) of the mean convalescent antibody level (39). The authors also applied a logistic model to calculate the 50% protective neutralization level, which estimates the antibody titre at which 50% of individuals are protected from infection, and is similar to the protective dose 50% that is sometimes used for influenza virus (46). The 50% protective neutralization level was found to be 20.2% (95% CI 14.4, 28.4) of the mean convalescent antibody level for symptomatic disease (corresponding to a neutralization titre of between 1:10 to 1:30 in most assays), which the authors estimate corresponds to 54 (95% CI 30–96) international units (IU)/ml. For severe disease, the 50% threshold was estimated to be only 3% (95% CI 0.71, 13.0%) of the mean convalescent level.

### Quality assessment

Studies were assessed for quality after full-text screening (Supplementary Table 2). Quality assessment was based on NIH NHLBI criteria (19), which centers on adequate description and transparency of methods, inclusion/exclusion criteria, and definitions. The criteria also includes an assessment of whether outcome variables were reported equally across all study participants. We excluded studies that did not adequately measure antibody levels or were missing information as to when antibody levels were obtained relative to infection, or had missing data or unclear methods related to antibody testing (47–51). Of the included studies, we noted that only five reported peak antibody levels at 30-60 days post infection or vaccination, the time period which would provide the most insight on peak antibody levels (31, 40, 42–44). Only seven studies reported antibody levels immediately prior to (within 30 days) re-infection or breakthrough (2, 26, 27, 31, 33, 40, 42), and only seven studies reporting SARS-CoV-2 lineage (22, 29, 34, 41–44).

## Discussion

The studies included in this review provided mixed evidence regarding a SARS-CoV-2 CoP, with a lack of standardization between laboratory methodology, assay targets, and sampling time points complicating comparisons and interpretation. Studies examining the relationship between antibody levels and VE presented high correlation coefficients, despite utilizing diverse data that included several vaccines and a variety of assays, VE endpoints and populations (35, 36, 38, 39). The robust correlations despite data heterogeneity support the concept of an anti-S antibody or neutralizing antibody CoP. Furthermore, several studies that explored differences in GMTs between cases and non-cases (34, 37) or associations between antibody levels and viral load with infection incidence or risk (2, 34, 37, 44), found statistically significant differences and associations. Taken together, these findings further support an antibody target as a potential correlate. However, while most studies that present aggregate measures support the existence of a humoral CoP, some individual-level data included in our review provided contradictory findings. Individuals described in case reports who experienced re-infection or breakthrough infection had considerable anti-S or neutralizing antibody levels pre-infection, and in some cases were at the upper limit or exceeded the limit of quantification of commercial assays (40, 41). Similarly, studies that attempted to estimate the contribution of antibody levels to VE measures (36–38) found that a substantial proportion of VE was not explained by antibody levels, suggesting that while important, anti-S or neutralizing antibodies are only one component of protection. These findings support observations from SARS-CoV-2 vaccine trial data, where one-dose vaccinated individuals are well-protected despite having very low levels of neutralizing antibodies. Consequently, these findings suggest that cellular immunity or non-neutralizing antibodies may also play a role in protection (36, 52).

From the reviewed literature, our analyses indicate that a humoral SARS-CoV-2 CoP may exist, but may be relative, such that the risk of infection is greatly reduced but not eliminated (8, 53). One analogous example of this is the influenza 50% protective dose, defined as the antibody concentration at which the risk of infection is reduced by half (9, 46). This is in contrast to a CoP that provides complete immunity (absolute correlate), as has been shown for viruses like rubella (9, 53). Khoury and colleagues provided evidence for a relative correlate in calculating a “50% protective neutralization level” across vaccine studies, and finding that lower antibody levels are required to prevent severe disease than to prevent infection (39). Estimating different thresholds by outcome is concordant with the concept of a relative threshold (9). Our findings are also in line with real-world observations where SARS-CoV-2 breakthrough cases are often mild or asymptomatic, suggesting that while there is not adequate immunity to prevent infection, there is adequate immunity to prevent symptomatic or severe disease (54–57). Furthermore, since mRNA vaccines produce high antibody levels while viral vector vaccines result in robust cellular immunity, it is also possible that the CoP following vaccination may differ by vaccine product (38, 52). The paucity of estimated quantitative thresholds therefore results in mostly indirect evidence included in our review.

Other data sources that were not eligible for inclusion in our review are supportive of a humoral CoP. For example, transfer of SARS-CoV-2 convalescent IgG to naïve rhesus macaques was found to be protective in a dose-dependent manner (5). Convalescent plasma has sometimes been found to be therapeutically effective in patients infected with SARS-CoV-2 (58), and monoclonal antibody therapy has been approved in the US for both treatment and prophylaxis (59). Although neither animal models nor manufactured monoclonal antibodies mimic the human immune response precisely, and the effectiveness of convalescent plasma therapy has been mixed (60), these data underscore the importance of humoral immunity for protection against SARS-CoV-2.

There were several limitations to the available literature for this systematic review. Many studies did not meet our inclusion criteria and pre-set definitions, which were designed to minimize bias. Our review included many different study types, including several case-reports, which generally provide a lower level of evidence and are particularly prone to bias (61, 62). There was heterogeneity in the targets that were measured, including neutralizing antibodies or antibody isotypes directed against spike (whole Spike, S1, receptor binding domain) or nucleocapsid protein. The included studies used different laboratory assays, which were generally not comparable. The WHO IS was seldom used, likely because it was not made available until late 2020. The diversity of laboratory assays and results precluded a meta-analysis of our data. To overcome the lack of calibration between laboratory assays, some studies normalized results against convalescent sera. However, since the humoral immune response to natural infection varies by age and disease severity (63), this method is not ideal for calibrating results. Most studies did not report which SARS-CoV-2 lineage was associated with the breakthrough or re-infection, with only a few studies reporting antibody levels preceding infection with a VOC. With the emergence of Omicron (B.1.1.529), the lack of Omicron-specific serological data prior to re-infection or breakthrough is unfortunate. Evidence based on *in vitro* neutralization assays suggests that, for immune responses to Omicron in individuals who have already been exposed to Ancestral SARS-CoV-2 antigens (whether through infection or vaccination), an Omicron CoP may be higher than for Ancestral SARS-CoV-2 or other VOCs, due to the reduced effectiveness of Ancestral antibodies for variant spike protein. To that point, Pfizer-BioNTech has reported a 25-fold reduction in neutralization titres against Omicron compared to Ancestral SARS-CoV-2 in in individuals vaccinated with two doses of BNT162b2 (64). Studies from South Africa and Germany report a reduction in neutralization up to 41-fold (65, 66), despite two or three doses of BNT162b2 or mRNA-1273 and previous infection. However, neutralization levels cannot be interpreted with regards to immunity in the absence of a CoP. This issue will be further complicated as the proportion of individuals with an Omicron-specific immune response due to infection, re-infection or breakthrough increases, especially if the clinical serology tools available for diagnostic purposes continue to use Ancestral SARS-CoV-2 antigens.

Since we restricted our review to evidence on a humoral CoP, we did not examine the role of cellular immunity. This is a limitation because both animal models and human studies have suggested that cellular immunity is likely integral to protection (5). Furthermore, the studies included in our review focused on systemic immunity. Since mucosal antibodies are a known element of SARS-CoV-2 immunity this was another limitation in our analysis (60). A recent study by Sheikh-Mohamed et al. supports the role of IgA in protection: breakthrough infection occurred in study participants with low levels of IgA compared to protected vaccinees, even if their levels of IgG were comparable (67). However, only three studies included in our review measured IgA levels, albeit in serum and not in mucosae (22, 29, 42). Since circulating IgA cannot be effectively transported into secretions (68), these studies cannot shed light on potential mucosal correlates of protection.

Our findings emphasize that further research into the role of humoral immunity, including non-neutralizing antibody, Fc effector functions and cellular and mucosal immunity is a priority, especially in the context of immune-evading variants like Omicron. The effect of lineage, vaccine product and the endpoint being measured (i.e. infection, symptomatic disease, severe disease) on the CoP are also essential questions. However, study designs that are best suited to assess whether a CoP exists are also quite complex and intensive. For example, human challenge studies are likely the most direct way to determine a CoP (69), but ethical issues that accompany these types of studies have limited their application (70). Finally, elucidating a CoP is directly related to raising global vaccine coverage and ending the COVID-19 pandemic. Currently, 40.5% of the world’s population has not been vaccinated against SARS-CoV-2 (71). The need to approve more vaccines is urgent, but placebo controlled trials have become difficult to perform (38). With this in mind, a temporary CoP, even if imperfect, would allow us to break through this impasse by performing non-inferiority studies to authorize new vaccine products.

Taken together, our findings suggest that humoral immunity is an integral part of protection against SARS-CoV-2, and that an antibody target is the most likely immune marker for a SARS-CoV-2 CoP. Although the evidence thus far supports the use of SARS-CoV-2 serology test results to confirm prior exposure to SARS-CoV-2, we currently do not have the tools to interpret serology with regards to protection.

Some jurisdictions have utilized serology testing in COVID-19 public health policies (72, 73), underscoring the urgency of elucidating a correlate of protection for SARS-CoV-2 to help guide public health decision making.

## Data Availability

All data in the present work are contained in the manuscript Supplementary Information

## Contributions

JP and SB conceptualised the study; JP and SO screened articles; JP, SO, JW, SB and MRG extracted data; SB wrote the original manuscript draft; all authors reviewed and edited the manuscript; more than one author accessed and verified the underlying data reported in the manuscript.

## Acknowledgements

MS is supported via salary awards from the BC Children’s Hospital Foundation, the Canadian Child Health Clinician Scientist Program and the Michael Smith Foundation for Health Research.

