## Supplementary Table 1 for "Does a humoral correlate of protection exist for SARS-CoV-2? A systematic review"

**LITERATURE SEARCH**

08/27/2021

COVID-19 antibody correlates of protection

Request prepared by: Library Services

Contact information:

Search results reporting

Databases Searched

| Database | Date searched | Records | Duplicates removed by database | Remaining |
| --- | --- | --- | --- | --- |
| MEDLINE (I) | 08/27/2021 | 1824 | 0 | 1824 |
| MEDLINE (II) | 08/27/2021 | 735 | 0 | 735 |

Results Totals

| Records source | Records |
| --- | --- |
| Records identified through database searching | 2559 |
| Duplicates removed by database | 0 |
| Duplicates removed by bibliographic management software | 101 |
| Total records after duplicates removed | 2458 |

Search strategies

MEDLINE

Ovid MEDLINE(R) ALL <1946 to August 26, 2021>

| # | Searches | Results |
| --- | --- | --- |
| 1 | COVID-19 Vaccines/ or (ncov or BNT162* or "mrna-1273").rn,nm,os,ox,ps,px,rs,rx. or ("Ad5-nCoV vaccine" or "BNT162 vaccine" or "ChAdOx1 COVID-19 vaccine" or "Covid-19 aAPC vaccine" or "lentiviral minigene vaccine of COVID-19 coronavirus" or "mRNA-1273 vaccine" or "recombinant SARS-CoV-2 vaccine NVX-cov2373" or "SARS-CoV-2 inactivated vaccines").rn. or (BNT162* or Comirnaty or CoronaVac or Covishield or Tozinameran or "mrna-1273*" or ChAdOx1* or AZD1222 or "Ad26.COV2.S" or "JNJ-78436735" or Novavax or NVX-CoV2373 or Ad5-nCoV or aAPC or "lentiviral minigene" or vaxzevria).ti,kf,kw. | 4959 |
| 2 | COVID-19/ or SARS-CoV-2/ or (Pandemics/ and Coronavirus Infections/) or "SARS-CoV-2 variants".os. or COVID-19 Testing/ or COVID-19 Serological Testing/ or ("COVID-19" or "SARS-CoV-2").nm,os,ox,ps,px,rs,rs. or ("2019 corona virus" or "2019 coronavirus" or "2019 ncov" or "corona virus 19" or "corona virus 2019" or "corona virus disease 19" or "corona virus disease 2019" or "corona virus epidemic*" or "corona virus outbreak*" or "corona virus pandemic*" or "corona virus response" or "coronavirus 19" or "coronavirus 2019" or "coronavirus disease 19" or "coronavirus disease 2019" or "coronavirus epidemic*" or "coronavirus outbreak*" or "coronavirus pandemic*" or "coronavirus response" or "covid 19" or "covid 2019" or "new corona virus" or "new coronavirus" or "novel corona virus" or "novel coronavirus" or "novel human coronavirus" or "sars coronavirus 2" or "sars cov 2" or "sars cov2" or "sars like coronavirus" or "severe acute respiratory syndrome corona virus 2" or "severe acute respiratory syndrome coronavirus 2" or "severe specific contagious pneumonia" or "wuhan corona virus" or "wuhan coronavirus" or ((pandemic* or novel or wuhan) adj3 (coronavirus* or "corona virus*" or betacoronavirus* or "beta coronavirus*" or "beta corona virus*" or pneumonia* or SARS or "severe acute respiratory syndrome")) or (pneumonia adj3 (coronavirus* or "corona virus*" or betacoronavirus* or "beta coronavirus*" or "beta corona virus*" or SARS or "severe acute respiratory syndrome")) or 2019ncov or covid or covid19 or covid2019 or ncov or sarscov2).kf,kw,ti. or (coronavirus* or "corona virus*" or betacoronavirus* or "beta coronavirus*" or "beta corona virus*" or pneumonia* or SARS or "severe acute respiratory syndrome").ti. or ("2019 corona virus" or "2019 coronavirus" or "2019 ncov" or "corona virus 19" or "corona virus 2019" or "corona virus disease 19" or "corona virus disease 2019" or "corona virus epidemic*" or "corona virus outbreak*" or "corona virus pandemic*" or "corona virus response" or "coronavirus 19" or "coronavirus 2019" or "coronavirus disease 19" or "coronavirus disease 2019" or "coronavirus epidemic*" or "coronavirus outbreak*" or "coronavirus pandemic*" or "coronavirus response" or "covid 19" or "covid 2019" or "new corona virus" or "new coronavirus" or "novel corona virus" or "novel coronavirus" or "novel human coronavirus" or "sars coronavirus 2" or "sars cov 2" or "sars cov2" or "sars like coronavirus" or "severe acute respiratory syndrome corona virus 2" or "severe acute respiratory syndrome coronavirus 2" or "severe specific contagious pneumonia" or "wuhan corona virus" or "wuhan coronavirus" or ((pandemic* or novel or wuhan) adj3 (coronavirus* or "corona virus*" or betacoronavirus* or "beta coronavirus*" or "beta corona virus*" or pneumonia* or SARS or "severe acute respiratory syndrome")) or (pneumonia adj3 (coronavirus* or "corona virus*" or betacoronavirus* or "beta coronavirus*" or "beta corona virus*" or SARS or "severe acute respiratory syndrome")) or 2019ncov or covid or covid19 or covid2019 or ncov or sarscov2).ab. /freq=2 | 251821 |
| 3 | Immunization Programs/ or Immunization Schedule/ or Immunization, Secondary/ or Immunization, Passive/ or Immunization/ or Immunogenicity, Vaccine/ or Immunotherapy, Active/ or Mass Vaccination/ or Vaccination Coverage/ or Vaccination/ or Vaccines, Attenuated/ or Vaccines, Combined/ or Vaccines, Synthetic/ or Vaccines/ or Viral Vaccines/ or COVID-19 Vaccines/ or vaccine*.rn. or (vaccin* or immunis* or immuniz* or innoculat* or inoculat* or jab* or shot* or injection* or nonimmunis* or nonimmuniz* or postvaccin* or postimmunis* or postimmuniz* or reimmunis* or reimmuniz* or revaccinat* or unimmunis* or unimmuniz* or ((one or single two or double or first or second or partial* or full) adj2 (dose* or injection* or injected or injecting)) or single-dose* or double-dose* or BNT162* or Comirnaty or CoronaVac or Covishield or Tozinameran or "mrna-1273*" or ChAdOx1* or AZD1222 or "Ad26.COV2.S" or "JNJ-78436735" or Novavax or NVX-CoV2373 or Ad5-nCoV or aAPC or "lentiviral minigene" or vaxzevria).ti,ab,kf,kw. | 1276813 |
| 4 | 2 and 3 | 25640 |
| 5 | Adaptive Immunity/ or Antibodies, Neutralizing/ or Antibodies, Viral/ or Antibody Affinity/ or Antibody Formation/ or Antibody Specificity/ or Antibodies, Monoclonal/ or Binding Sites, Antibody/ or exp Antibodies/ or exp Antigen-Antibody Reactions/ or Antibody-Dependent Enhancement/ or Antibody-Dependent Cell Cytotoxicity/ or Lymphocytes/ or exp B-Lymphocytes/ or B-Lymphocyte subsets/ or Lymphocyte Activation/ or Complement Activation/ or Th2 Cells/ or Immune System Phenomena/ or Immunity/ or Immunity, Active/ or Immunity, Humoral/ or Immunity, Mucosal/ or Immunologic Factors/ or Immunologic Surveillance/ or Immunity, Innate/ or Immunity, Heterologous/ or Immunity, Maternally-Acquired/ or Immune Sera/ or exp Immunoglobulins/ or Immunoglobulin M/ or Immunoglobulin G/ or exp Immunomodulation/ or Immunologic Memory/ or Models, Immunological/ or Cross Protection/im or Cross Reactions/im or Reinfection/im or Recurrence/im or "Severity of Illness Index"/im or Viral Load/im or Convalescence/im or Seroconversion/ or Seroepidemiologic Studies/ or Neutralization Tests/ or Enzyme-Linked Immunosorbent Assay/ or Protein Domains/im or Receptors, Virus/im or Spike Glycoprotein, Coronavirus/im or Coronavirus Nucleocapsid Proteins/im or *COVID-19 Vaccines/im or *COVID-19/im or *SARS-CoV-2/im or (antibody or antibodies or antibody-positiv* or antibody-negativ* or "complement activation" or ((humoral or humoural or adaptive or antibody-mediated) adj3 (respon* or immun*)) or immunity or "immune system*" or "immune marker*" or ((immun* or sero*) adj3 respon*) or immunolog* or immunomodulat* or B-lymphocyte* or "B-cell*").ti,ab,kf,kw. or (immun* or immunoglobulin* or IgA or IgD or IgE or IgG or IgM or sero* or sera*).ti,kf,kw. | 2782702 |
| 6 | ("correlat* of protection" or "correlat* with protection" or "immune correlat*" or "correlat* of immunity" or "correlat* of neutraliz*" or "correlat* of neutralis*" or "measur* of protection" or "correlat* of infectivity" or "protective correlat*" or ((protective or protection) adj5 (level* or threshold* or marker* or limit* or amount* or mean* or baseline or quantif*)) or ((correlat* or marker* or threshold* or indicat* or measur*) adj5 (immun* or protect*)) or ((antibody or antibodies or titer* or titre*) adj3 (correlat* or threshold*)) or (protect* adj5 (immunity or antibod* or titer* or titre*)) or ((neutraliz* or neutralis*) adj2 (level* or amount* or limit* or mean* or baseline or quantif* or titer* or titre*)) or (predict* adj3 (immun* or protect*)) or "protective immunity" or "surrogate endpoint*").ti,ab,kf,kw. or (protection or protective or protected or ((antibody or antibodies) adj2 (status or level*))).ti. | 370204 |
| 7 | (1 or 4) and 5 and 6 | 2048 |
| 8 | 2 and 5 and 6 | 3863 |
| 9 | 7 or 8 | 3869 |
| 10 | limit 9 to yr="2019 -Current" | 2589 |
| 11 | limit 10 to (english or french) | 2559 |
| 12 | Reinfection/ or Recurrence/ or Follow-Up Studies/ or ((Convalescence/ or "Severity of Illness Index"/ or Viral Load/ or Asymptomatic Infections/) and (re-infect* or reinfect*).ti,kf,kw.) | 827685 |
| 13 | (((previous* or prior or past or post or postcovid* or post-covid* or postsars* or post-sars* or after or following or recover*) adj2 infect*) or re-infect* or reinfect* or "after first infection" or "after infection").ti,ab,kf,kw. or (convalescence or convalescent).ti,kf,kw. | 135528 |
| 14 | 12 or 13 | 955646 |
| 15 | Treatment Failure/ or ((vaccin* or immunis* or immuniz*) adj3 fail*).ab,kf,kw,ti. | 39442 |
| 16 | (((breakthrough* or "break through*") adj5 (vaccin* or infection*)) or "breakthrough rate*").ab,kf,kw,ti. | 1398 |
| 17 | (("second dose" or "two doses" or "fully vaccinated") adj5 (infected or infection* or contracted or acquired or acquisition or caught or got* or transmi* or spread* or outbreak* or cluster* or communicab* or contagious* or epidemic* or occurrence or case or cases)).ab,kf,kw,ti. | 378 |
| 18 | ((postvaccin* or postimmuniz* or postimmunis* or "post-vaccin*" or "post-immuniz*" or "post-immunis*") adj5 (infected or infection* or contracted or acquired or acquisition or caught or got* or transmi* or spread* or outbreak* or cluster* or communicab* or contagious* or epidemic* or occurrence or case or cases)).ab,kf,kw,ti. | 618 |
| 19 | (((receiv* or recipient* or complet* or already or post or previously or after or following or "subsequent to" or since or had or despite or "in spite of") adj5 (immunis* or immuniz* or inoculat* or innoculat* or vaccin* or revaccinat* or reimmuniz* or reimmunis* or dose or doses) adj5 (infected or infection* or contracted or acquired or acquisition or caught or got* or transmi* or spread* or outbreak* or cluster* or communicab* or contagious* or epidemic* or occurrence or case or cases)) or ((infected or infection* or contracted or acquired or acquisition or caught or got* or transmi* or spread* or outbreak* or cluster* or communicab* or contagious* or epidemic* or occurrence or case or cases) adj5 (receiv* or recipient* or complet* or already or post or previously or after or following or "subsequent to" or since or had or despite or "in spite of") adj5 (immunis* or immuniz* or inoculat* or innoculat* or vaccin* or revaccinat* or reimmuniz* or reimmunis* or dose or doses))).ab,kf,kw,ti. | 19047 |
| 20 | ((vaccinated or immunized or immunised) adj5 (infected or infection* or contracted or acquired or acquisition or caught or got* or transmi* or spread* or outbreak* or cluster* or communicab* or contagious* or epidemic* or occurrence or case or cases)).ab,kf,kw,ti. | 8545 |
| 21 | ((vaccinated or immunized or immunised) adj10 (person or persons or people or man or men or woman or women or adult or adults or patient or patients or individual or individuals or responder or responders or practitioner* or staff or personnel or worker* or employee* or provider* or technician* or clinician* or doctor* or nurse or nurses or paramedic* or physician* or hospitalist* or pharmacist*) adj10 (infected or infection* or contracted or acquired or acquisition or caught or got* or transmi* or spread* or outbreak* or cluster* or communicab* or contagious* or epidemic* or occurrence or case or cases)).ab,kf,kw,ti. | 2764 |
| 22 | ((escape or escapes or escaped or escaping or evade or evades or evaded or evading or evasion) adj5 ("Ad26.COV2.S" or AZD1222 or BNT162* or ChAdOx1* or Comirnaty or immunis* or immuniz* or "JNJ-78436735" or "mrna-1273*" or Novavax or NVX-CoV2373 or PittCoVacc* or postvaccin* or postimmunis* or postimmuniz* or reimmunis* or reimmuniz* or revacciat* or tozinameran or vaccin*)).ab,kf,kw,ti. | 992 |
| 23 | 15 or 16 or 17 or 18 or 19 or 20 or 21 or 22 | 67744 |
| 24 | (wane or wanes or waned or waning).kf,kw,ti. | 559 |
| 25 | ((decay* or declin* or decreas* or impair* or duration or fail* or fall* or "go* down" or lack or lacks or lacked or lacking or lose or loses or losing or loss or lost or low or lower* or lesser* or poor or poorer* or "adversely affect*" or reduc*) adj5 (protect* or effectiv* or immunity or "immune respon*" or antibod* or sero*)).kf,kw,ti. and (infected or infection* or contracted or acquired or acquisition or caught or got* or transmi* or spread* or outbreak* or cluster* or communicab* or contagious* or epidemic* or occurrence or case or cases).ab,kf,kw,ti. | 6132 |
| 26 | ((decay* or declin* or decreas* or impair* or duration or fail* or fall* or "go* down" or lack or lacks or lacked or lacking or lose or loses or losing or loss or lost or low or lower* or lesser* or poor or poorer* or "adversely affect*" or reduc*) adj5 (protect* or effectiv* or immunity or "immune respon*" or antibod* or sero*)).ab,kf,kw,ti. and (infected or infection* or contracted or acquired or acquisition or caught or got* or transmi* or spread* or outbreak* or cluster* or communicab* or contagious* or epidemic* or occurrence or case or cases).kf,kw,ti. | 29587 |
| 27 | 24 or 25 or 26 | 34166 |
| 28 | (1 or 4) and (14 or 23 or 27) and (5 and 6) | 595 |
| 29 | 2 and (14 or 27) and (5 and 6) | 876 |
| 30 | 28 or 29 | 993 |
| 31 | limit 30 to yr="2019 -Current" | 739 |
| 32 | limit 31 to (english or french) | 735 |
| 33 | 11 not 32 | 1824 |

**LITERATURE SEARCH**

09/03/2021

COVID-19 antibody correlates of protection: Supplementary Databases

Request prepared by: Library Services

Contact information:

Search results reporting

Databases Searched

| Database | Date searched | Records | Duplicates removed by database | Remaining |
| --- | --- | --- | --- | --- |
| EMBASE (I) | 09/03/2021 | 1595 | 0 | 1595 |
| EMBASE (II) | 09/03/2021 | 473 | 0 | 473 |
| GLOBAL HEALTH (I) | 09/03/2021 | 803 | 0 | 803 |
| GLOBAL HEALTH (II) | 09/03/2021 | 306 | 0 | 306 |
| BIOSIS Previews (I) | 09/03/2021 | 819 | 0 | 819 |
| BIOSIS Previews (II) | 09/03/2021 | 359 | 0 | 359 |
| SCOPUS | 09/03/2021 | 692 | 0 | 692 |

Results Totals

| Records source | Records |
| --- | --- |
| Records identified through database searching | 5047 |
| Duplicates removed by database | 0 |
| Duplicates removed by bibliographic management software | 4,159 |
| Total records after duplicates removed  *(Search I combined: 766 records)*  *(Search II combined: 122 records)* | 888 |

Search strategies

EMBASE

Embase <1974 to 2021 Week 34>

| # | Searches | Results |
| --- | --- | --- |
| 1 | SARS-CoV-2 vaccine/ or ("Ad5-nCoV vaccine" or "BNT162 vaccine" or "ChAdOx1 COVID-19 vaccine" or "Covid-19 aAPC vaccine" or "lentiviral minigene vaccine of COVID-19 coronavirus" or "mRNA-1273 vaccine" or "PittCoVacc" or "recombinant SARS-CoV-2 vaccine NVX-cov2373" or "SARS-CoV-2 inactivated vaccines").rn. or (BNT162* or Comirnaty or CoronaVac or Covishield or Tozinameran or "mrna-1273*" or ChAdOx1* or AZD1222 or "Ad26.COV2.S" or "JNJ-78436735" or Novavax or NVX-CoV2373 or PittCoVacc* or Ad5-nCoV or aAPC or "lentiviral minigene" or vaxzevria).ti,kw. | 5424 |
| 2 | coronavirus disease 2019/ or Severe acute respiratory syndrome coronavirus 2/ or (pandemic/ and Coronavirus infection/) or ("2019 corona virus" or "2019 coronavirus" or "2019 ncov" or "corona virus 19" or "corona virus 2019" or "corona virus disease 19" or "corona virus disease 2019" or "corona virus epidemic*" or "corona virus outbreak*" or "corona virus pandemic*" or "corona virus response" or "coronavirus 19" or "coronavirus 2019" or "coronavirus disease 19" or "coronavirus disease 2019" or "coronavirus epidemic*" or "coronavirus outbreak*" or "coronavirus pandemic*" or "coronavirus response" or "covid 19" or "covid 2019" or "new corona virus" or "new coronavirus" or "novel corona virus" or "novel coronavirus" or "novel human coronavirus" or "sars coronavirus 2" or "sars cov 2" or "sars cov2" or "sars like coronavirus" or "severe acute respiratory syndrome corona virus 2" or "severe acute respiratory syndrome coronavirus 2" or "severe specific contagious pneumonia" or "wuhan corona virus" or "wuhan coronavirus" or ((pandemic* or novel or wuhan) adj3 (coronavirus* or "corona virus*" or betacoronavirus* or "beta coronavirus*" or "beta corona virus*" or pneumonia* or SARS or "severe acute respiratory syndrome")) or (pneumonia adj3 (coronavirus* or "corona virus*" or betacoronavirus* or "beta coronavirus*" or "beta corona virus*" or SARS or "severe acute respiratory syndrome")) or 2019ncov or covid or covid19 or covid2019 or ncov or sarscov2).kw,ti. or ("SARS-Cov-2 variant*" or VUI-202012-01 or "VOC-202012/01" or "B.1.1.7" or B117 or "B.1.351" or B1351 or "B.1.617" or B1617 or "P.1" or P1 or coronavirus* or "corona virus*" or betacoronavirus* or "beta coronavirus*" or "beta corona virus*" or pneumonia* or SARS or "severe acute respiratory syndrome").ti. or ((covid* or sars* or variant*) and (alpha or beta or gamma or delta or "variant* of concern" or "VOC")).ti. | 279851 |
| 3 | immunization/ or secondary immunization/ or vaccine immunogenicity/ or active immunotherapy/ or mass immunization/ or vaccination/ or vaccination coverage/ or vaccination refusal/ or live vaccine/ or vaccine/ or recombinant vaccine/ or virus vaccine/ or SARS-CoV-2 vaccine/ or (vaccin* or immunis* or immuniz* or innoculat* or inoculat* or jab* or shot* or injection* or nonimmunis* or nonimmuniz* or postvaccin* or postimmunis* or postimmuniz* or reimmunis* or reimmuniz* or revaccinat* or unimmunis* or unimmuniz* or ((one or single two or double or first or second or partial* or full) adj2 (dose* or injection* or injected or injecting)) or single-dose* or double-dose* or BNT162* or Comirnaty or CoronaVac or Covishield or Tozinameran or "mrna-1273*" or ChAdOx1* or AZD1222 or "Ad26.COV2.S" or "JNJ-78436735" or Novavax or NVX-CoV2373 or Ad5-nCoV or aAPC or "lentiviral minigene" or vaxzevria).ti,ab,kw. | 1572516 |
| 4 | 2 and 3 | 28195 |
| 5 | adaptive immunity/ or neutralizing antibody/ or virus antibody/ or antibody affinity/ or antibody production/ or antibody specificity/ or monoclonal antibody/ or antibody combining site/ or exp antibody/ or exp antigen antibody reaction/ or antibody dependent enhancement/ or antibody dependent cellular cytotoxicity/ or antibody response/ or lymphocyte/ or exp B lymphocyte/ or B lymphocyte subpopulation/ or lymphocyte activation/ or complement activation/ or Th2 cell/ or immunity/ or active immunization/ or humoral immunity/ or mucosal immunity/ or immunologic factor/ or immunosurveillance/ or innate immunity/ or heterologous immunity/ or passive immunization/ or antiserum/ or exp immunoglobulin/ or immunoglobulin G/ or immunological memory/ or biological model/ or cross protection/ or cross reaction/ or reinfection/ or recurrent disease/ or "severity of illness index"/ or virus load/ or convalescence/ or seroconversion/ or seroepidemiology/ or serodiagnosis/ or enzyme linked immunosorbent assay/ or protein domain/ or virus receptor/ or coronavirus spike glycoprotein/ or nucleocapsid protein/ or (antibody or antibodies or antibody-positiv* or antibody-negativ* or "complement activation" or ((humoral or humoural or adaptive or antibody-mediated) adj3 (respon* or immun*)) or immunity or "immune system*" or "immune marker*" or ((immun* or sero*) adj3 respon*) or immunolog* or immunomodulat* or B-lymphocyte* or "B-cell*").ti,kw. or (antibody or antibodies or antibody-positiv* or antibody-negativ* or "complement activation" or ((humoral or humoural or adaptive or antibody-mediated) adj3 (respon* or immun*)) or immunity or "immune system*" or "immune marker*" or ((immun* or sero*) adj3 respon*) or immunolog* or immunomodulat* or B-lymphocyte* or "B-cell*").ab. /freq=2 or (immun* or immunoglobulin* or IgA or IgD or IgE or IgG or IgM or sero* or sera*).ti,kw. | 3948248 |
| 6 | ("correlat* of protection" or "correlat* with protection" or "immune correlat*" or "correlat* of immunity" or "correlat* of neutraliz*" or "correlat* of neutralis*" or "measur* of protection" or "correlat* of infectivity" or "protective correlat*" or ((protective or protection) adj5 (level* or threshold* or marker* or limit* or amount* or mean* or baseline or quantif*)) or ((correlat* or marker* or threshold* or indicat* or measur*) adj5 (immun* or protect*)) or ((antibody or antibodies or titer* or titre*) adj3 (correlat* or threshold*)) or (protect* adj5 (immunity or antibod* or titer* or titre*)) or ((neutraliz* or neutralis*) adj2 (level* or amount* or limit* or mean* or baseline or quantif* or titer* or titre*)) or (predict* adj3 (immun* or protect*)) or "protective immunity" or "surrogate endpoint*").ti,ab,kw. or (protection or protective or protected or ((antibody or antibodies) adj2 (status or level*))).ti. | 474113 |
| 7 | (1 or 4) and 5 and 6 | 1908 |
| 8 | 2 and 5 and 6 | 4025 |
| 9 | 7 or 8 | 4038 |
| 10 | limit 9 to conference abstract | 412 |
| 11 | 9 not 10 | 3626 |
| 12 | 11 not (editorial or letter or note).pt. | 3467 |
| 13 | limit 12 to yr="2019 -Current" | 2123 |
| 14 | limit 13 to (english or french) | 2068 |
| 15 | reinfection/ or recurrent disease/ or ((follow up/ or convalescence/ or "severity of illness index"/ or virus load/ or asymptomatic infection/) and (re-infect* or reinfect*).ti,kw.) | 199957 |
| 16 | (((previous* or prior or past or post or postcovid* or post-covid* or postsars* or post-sars* or after or following or recover* or convalescen* or "after infection") adj2 infect*) or re-infect* or reinfect* or "after first infection").ti,ab,kw. | 167755 |
| 17 | 15 or 16 | 358275 |
| 18 | treatment failure/ or ((vaccin* or immunis* or immuniz*) adj3 fail*).ab,kw,ti. | 142466 |
| 19 | (((breakthrough* or "break through*") adj5 (vaccin* or infection*)) or "breakthrough rate*").ab,kw,ti. | 1940 |
| 20 | (("second dose" or "two doses" or "fully vaccinated") adj5 (infected or infection* or contracted or acquired or acquisition or caught or got* or transmi* or spread* or outbreak* or cluster* or communicab* or contagious* or epidemic* or occurrence or case or cases)).ab,kw,ti. | 476 |
| 21 | ((postvaccin* or postimmuniz* or postimmunis* or "post-vaccin*" or "post-immuniz*" or "post-immunis*") adj5 (infected or infection* or contracted or acquired or acquisition or caught or got* or transmi* or spread* or outbreak* or cluster* or communicab* or contagious* or epidemic* or occurrence or case or cases)).ab,kw,ti. | 716 |
| 22 | (((receiv* or recipient* or complet* or already or post or previously or after or following or "subsequent to" or since or had or despite or "in spite of") adj5 (immunis* or immuniz* or inoculat* or innoculat* or vaccin* or revaccinat* or reimmuniz* or reimmunis* or dose or doses) adj5 (infected or infection* or contracted or acquired or acquisition or caught or got* or transmi* or spread* or outbreak* or cluster* or communicab* or contagious* or epidemic* or occurrence or case or cases)) or ((infected or infection* or contracted or acquired or acquisition or caught or got* or transmi* or spread* or outbreak* or cluster* or communicab* or contagious* or epidemic* or occurrence or case or cases) adj5 (receiv* or recipient* or complet* or already or post or previously or after or following or "subsequent to" or since or had or despite or "in spite of") adj5 (immunis* or immuniz* or inoculat* or innoculat* or vaccin* or revaccinat* or reimmuniz* or reimmunis* or dose or doses))).ab,kw,ti. | 23864 |
| 23 | ((vaccinated or immunized or immunised) adj5 (infected or infection* or contracted or acquired or acquisition or caught or got* or transmi* or spread* or outbreak* or cluster* or communicab* or contagious* or epidemic* or occurrence or case or cases)).ab,kw,ti. | 9423 |
| 24 | ((vaccinated or immunized or immunised) adj10 (person or persons or people or man or men or woman or women or adult or adults or patient or patients or individual or individuals or responder or responders or practitioner* or staff or personnel or worker* or employee* or provider* or technician* or clinician* or doctor* or nurse or nurses or paramedic* or physician* or hospitalist* or pharmacist*) adj10 (infected or infection* or contracted or acquired or acquisition or caught or got* or transmi* or spread* or outbreak* or cluster* or communicab* or contagious* or epidemic* or occurrence or case or cases)).ab,kw,ti. | 3485 |
| 25 | ((escape or escapes or escaped or escaping or evade or evades or evaded or evading or evasion) adj5 ("Ad26.COV2.S" or AZD1222 or BNT162* or ChAdOx1* or Comirnaty or immunis* or immuniz* or "JNJ-78436735" or "mrna-1273*" or Novavax or NVX-CoV2373 or PittCoVacc* or postvaccin* or postimmunis* or postimmuniz* or reimmunis* or reimmuniz* or revacciat* or tozinameran or vaccin*)).ab,kw,ti. | 1175 |
| 26 | 18 or 19 or 20 or 21 or 22 or 23 or 24 or 25 | 176896 |
| 27 | (wane or wanes or waned or waning).kw,ti. | 646 |
| 28 | ((decay* or declin* or decreas* or impair* or duration or fail* or fall* or "go* down" or lack or lacks or lacked or lacking or lose or loses or losing or loss or lost or low or lower* or lesser* or poor or poorer* or "adversely affect*" or reduc*) adj5 (protect* or effectiv* or immunity or "immune respon*" or antibod* or sero*)).kw,ti. and (infected or infection* or contracted or acquired or acquisition or caught or got* or transmi* or spread* or outbreak* or cluster* or communicab* or contagious* or epidemic* or occurrence or case or cases).ab,kw,ti. | 8059 |
| 29 | ((decay* or declin* or decreas* or impair* or duration or fail* or fall* or "go* down" or lack or lacks or lacked or lacking or lose or loses or losing or loss or lost or low or lower* or lesser* or poor or poorer* or "adversely affect*" or reduc*) adj5 (protect* or effectiv* or immunity or "immune respon*" or antibod* or sero*)).ab,kw,ti. and (infected or infection* or contracted or acquired or acquisition or caught or got* or transmi* or spread* or outbreak* or cluster* or communicab* or contagious* or epidemic* or occurrence or case or cases).kw,ti. | 38103 |
| 30 | 27 or 28 or 29 | 44270 |
| 31 | (1 or 4) and (17 or 26 or 30) and (5 and 6) | 477 |
| 32 | 2 and (17 or 30) and (5 and 6) | 731 |
| 33 | 31 or 32 | 840 |
| 34 | limit 33 to conference abstract | 100 |
| 35 | 33 not 34 | 740 |
| 36 | 35 not (editorial or letter or note).pt. | 720 |
| 37 | limit 36 to yr="2019 -Current" | 482 |
| 38 | limit 37 to (english or french) | 473 |
| 39 | 14 not 38 | 1595 |

Global Health

Global Health <1973 to 2021 Week 35>

| # | Searches | Results |
| --- | --- | --- |
| 1 | ((vaccines/ or immunization/ or vaccination/ or synthetic vaccines/ or combined vaccines/ or vaccine development/ or candidate vaccines/ or messenger RNA/) and (ncov or BNT162* or "mrna-1273").ti,ab,hw.) or (BNT162* or Comirnaty or CoronaVac or Covishield or Tozinameran or "mrna-1273*" or ChAdOx1* or AZD1222 or "Ad26.COV2.S" or "JNJ-78436735" or Novavax or NVX-CoV2373 or Ad5-nCoV or aAPC or "lentiviral minigene" or vaxzevria).ti,ab,hw. | 590 |
| 2 | ("Severe acute respiratory syndrome coronavirus 2" or "coronavirus disease 2019" or "COVID-19" or "SARS-CoV-2").ti,hw,id. or ("2019 corona virus" or "2019 coronavirus" or "2019 ncov" or "corona virus 19" or "corona virus 2019" or "corona virus disease 19" or "corona virus disease 2019" or "corona virus epidemic*" or "corona virus outbreak*" or "corona virus pandemic*" or "corona virus response" or "coronavirus 19" or "coronavirus 2019" or "coronavirus disease 19" or "coronavirus disease 2019" or "coronavirus epidemic*" or "coronavirus outbreak*" or "coronavirus pandemic*" or "coronavirus response" or "covid 19" or "covid 2019" or "new corona virus" or "new coronavirus" or "novel corona virus" or "novel coronavirus" or "novel human coronavirus" or "sars coronavirus 2" or "sars cov 2" or "sars cov2" or "sars like coronavirus" or "severe acute respiratory syndrome corona virus 2" or "severe acute respiratory syndrome coronavirus 2" or "severe specific contagious pneumonia" or "wuhan corona virus" or "wuhan coronavirus" or ((pandemic* or novel or wuhan) adj3 (coronavirus* or "corona virus*" or betacoronavirus* or "beta coronavirus*" or "beta corona virus*" or pneumonia* or SARS or "severe acute respiratory syndrome")) or (pneumonia adj3 (coronavirus* or "corona virus*" or betacoronavirus* or "beta coronavirus*" or "beta corona virus*" or SARS or "severe acute respiratory syndrome")) or 2019ncov or covid or covid19 or covid2019 or ncov or sarscov2).ti,hw. or ("SARS-Cov-2 variant*" or VUI-202012-01 or "VOC-202012/01" or "B.1.1.7" or B117 or "B.1.351" or B1351 or "B.1.617" or B1617 or "P.1" or P1 or coronavirus* or "corona virus*" or betacoronavirus* or "beta coronavirus*" or "beta corona virus*" or pneumonia* or SARS or "severe acute respiratory syndrome").ti. or ((covid* or sars* or variant*) and (alpha or beta or gamma or delta or "variant* of concern" or "VOC")).ti. or ("2019 corona virus" or "2019 coronavirus" or "2019 ncov" or "corona virus 19" or "corona virus 2019" or "corona virus disease 19" or "corona virus disease 2019" or "corona virus epidemic*" or "corona virus outbreak*" or "corona virus pandemic*" or "corona virus response" or "coronavirus 19" or "coronavirus 2019" or "coronavirus disease 19" or "coronavirus disease 2019" or "coronavirus epidemic*" or "coronavirus outbreak*" or "coronavirus pandemic*" or "coronavirus response" or "covid 19" or "covid 2019" or "new corona virus" or "new coronavirus" or "novel corona virus" or "novel coronavirus" or "novel human coronavirus" or "sars coronavirus 2" or "sars cov 2" or "sars cov2" or "sars like coronavirus" or "severe acute respiratory syndrome corona virus 2" or "severe acute respiratory syndrome coronavirus 2" or "severe specific contagious pneumonia" or "wuhan corona virus" or "wuhan coronavirus" or ((pandemic* or novel or wuhan) adj3 (coronavirus* or "corona virus*" or betacoronavirus* or "beta coronavirus*" or "beta corona virus*" or pneumonia* or SARS or "severe acute respiratory syndrome")) or (pneumonia adj3 (coronavirus* or "corona virus*" or betacoronavirus* or "beta coronavirus*" or "beta corona virus*" or SARS or "severe acute respiratory syndrome")) or 2019ncov or covid or covid19 or covid2019 or ncov or sarscov2).ab. /freq=2 | 73545 |
| 3 | vaccines/ or immunization programmes/ or immunization/ or vaccination/ or passive immunization/ or synthetic vaccines/ or combined vaccines/ or vaccine development/ or candidate vaccines/ or (vaccin* or immunis* or immuniz* or innoculat* or inoculat* or jab* or shot* or injection* or nonimmunis* or nonimmuniz* or postvaccin* or postimmunis* or postimmuniz* or reimmunis* or reimmuniz* or revaccinat* or unimmunis* or unimmuniz* or ((one or single two or double or first or second or partial* or full) adj2 (dose* or injection* or injected or injecting)) or single-dose* or double-dose* or BNT162* or Comirnaty or CoronaVac or Covishield or Tozinameran or "mrna-1273*" or ChAdOx1* or AZD1222 or "Ad26.COV2.S" or "JNJ-78436735" or Novavax or NVX-CoV2373 or Ad5-nCoV or aAPC or "lentiviral minigene" or vaxzevria).ti,ab,hw. | 275347 |
| 4 | 2 and 3 | 8485 |
| 5 | neutralizing antibodies/ or monoclonal antibodies/ or exp antibodies/ or antibody formation/ or antibody testing/ or antigen antibody reactions/ or humoral immunity/ or immune serum/ or immunity/ or immune response/ or immune system/ or cross immunity/ or cross reaction/ or passive immunity/ or immune competence/ or immunological factors/ or immunology/ or antibody dependent cellular cytotoxicity/ or lymphocytes/ or B lymphocytes/ or lymphocyte transformation/ or complement activation/ or Th2 lymphocytes/ or exp immunoglobulins/ or reinfection/ or viral load/ or seroconversion/ or seroprevalence/ or neutralization tests/ or virus neutralization/ or ELISA/ or (antibody or antibodies or antibody-positiv* or antibody-negativ* or "complement activation" or ((humoral or humoural or adaptive or antibody-mediated) adj3 (respon* or immun*)) or immunity or "immune system*" or "immune marker*" or ((immun* or sero*) adj3 respon*) or immunolog* or immunomodulat* or B-lymphocyte* or "B-cell*").ti,ab,hw. or (immun* or immunoglobulin* or IgA or IgD or IgE or IgG or IgM or sero* or sera*).ti,hw. | 669065 |
| 6 | ("correlat* of protection" or "correlat* with protection" or "immune correlat*" or "correlat* of immunity" or "correlat* of neutraliz*" or "correlat* of neutralis*" or "measur* of protection" or "correlat* of infectivity" or "protective correlat*" or ((protective or protection) adj5 (level* or threshold* or marker* or limit* or amount* or mean* or baseline or quantif*)) or ((correlat* or marker* or threshold* or indicat* or measur*) adj5 (immun* or protect*)) or ((antibody or antibodies or titer* or titre*) adj3 (correlat* or threshold*)) or (protect* adj5 (immunity or antibod* or titer* or titre*)) or ((neutraliz* or neutralis*) adj2 (level* or amount* or limit* or mean* or baseline or quantif* or titer* or titre*)) or (predict* adj3 (immun* or protect*)) or "protective immunity" or "surrogate endpoint*").ti,ab,hw. or (protection or protective or protected or ((antibody or antibodies) adj2 (status or level*))).ti. | 84772 |
| 7 | 5 and 6 | 47062 |
| 8 | (1 or 4) and 7 | 679 |
| 9 | 2 and 7 | 1505 |
| 10 | 8 or 9 | 1518 |
| 11 | limit 10 to yr="2019 -Current" | 1135 |
| 12 | limit 11 to (english or french) | 1109 |
| 13 | reinfection/ or ((viral load/ or asymptomatic infections/) and (re-infect* or reinfect*).ti,hw.) | 2358 |
| 14 | (((previous* or prior or past or post or postcovid* or post-covid* or postsars* or post-sars* or after or following or recover* or convalescen* or "after infection") adj3 infect*) or re-infect* or reinfect* or "after first infection").ti,ab,hw. | 59349 |
| 15 | 13 or 14 | 59349 |
| 16 | treatment failure/ or ((vaccin* or immunis* or immuniz*) adj3 fail*).ti,ab,hw. | 10182 |
| 17 | (((breakthrough* or "break through*") adj5 (vaccin* or infection*)) or "breakthrough rate*").ti,ab,hw. | 711 |
| 18 | (("second dose" or "two doses" or "fully vaccinated") adj5 (infected or infection* or contracted or acquired or acquisition or caught or got* or transmi* or spread* or outbreak* or cluster* or communicab* or contagious* or epidemic* or occurrence or case or cases)).ti,ab,hw. | 189 |
| 19 | ((postvaccin* or postimmuniz* or postimmunis* or "post-vaccin*" or "post-immuniz*" or "post-immunis*") adj5 (infected or infection* or contracted or acquired or acquisition or caught or got* or transmi* or spread* or outbreak* or cluster* or communicab* or contagious* or epidemic* or occurrence or case or cases)).ti,ab,hw. | 253 |
| 20 | (((receiv* or recipient* or complet* or already or post or previously or after or following or "subsequent to" or since or had or despite or "in spite of") adj5 (immunis* or immuniz* or inoculat* or innoculat* or vaccin* or revaccinat* or reimmuniz* or reimmunis* or dose or doses) adj5 (infected or infection* or contracted or acquired or acquisition or caught or got* or transmi* or spread* or outbreak* or cluster* or communicab* or contagious* or epidemic* or occurrence or case or cases)) or ((infected or infection* or contracted or acquired or acquisition or caught or got* or transmi* or spread* or outbreak* or cluster* or communicab* or contagious* or epidemic* or occurrence or case or cases) adj5 (receiv* or recipient* or complet* or already or post or previously or after or following or "subsequent to" or since or had or despite or "in spite of") adj5 (immunis* or immuniz* or inoculat* or innoculat* or vaccin* or revaccinat* or reimmuniz* or reimmunis* or dose or doses))).ti,ab,hw. | 7993 |
| 21 | ((vaccinated or immunized or immunised) adj5 (infected or infection* or contracted or acquired or acquisition or caught or got* or transmi* or spread* or outbreak* or cluster* or communicab* or contagious* or epidemic* or occurrence or case or cases)).ti,ab,hw. | 3928 |
| 22 | ((vaccinated or immunized or immunised) adj10 (person or persons or people or man or men or woman or women or adult or adults or patient or patients or individual or individuals or responder or responders or practitioner* or staff or personnel or worker* or employee* or provider* or technician* or clinician* or doctor* or nurse or nurses or paramedic* or physician* or hospitalist* or pharmacist*) adj10 (infected or infection* or contracted or acquired or acquisition or caught or got* or transmi* or spread* or outbreak* or cluster* or communicab* or contagious* or epidemic* or occurrence or case or cases)).ti,ab,hw. | 1646 |
| 23 | ((escape or escapes or escaped or escaping or evade or evades or evaded or evading or evasion) adj5 ("Ad26.COV2.S" or AZD1222 or BNT162* or ChAdOx1* or Comirnaty or immunis* or immuniz* or "JNJ-78436735" or "mrna-1273*" or Novavax or NVX-CoV2373 or PittCoVacc* or postvaccin* or postimmunis* or postimmuniz* or reimmunis* or reimmuniz* or revacciat* or tozinameran or vaccin*)).ti,ab,hw. | 399 |
| 24 | 16 or 17 or 18 or 19 or 20 or 21 or 22 or 23 | 22285 |
| 25 | (wane or wanes or waned or waning).ti,hw. | 119 |
| 26 | ((decay* or declin* or decreas* or impair* or duration or fail* or fall* or "go* down" or lack or lacks or lacked or lacking or lose or loses or losing or loss or lost or low or lower* or lesser* or poor or poorer* or "adversely affect*" or reduc*) adj5 (protect* or effectiv* or immunity or "immune respon*" or antibod* or sero*)).ti,hw. and (infected or infection* or contracted or acquired or acquisition or caught or got* or transmi* or spread* or outbreak* or cluster* or communicab* or contagious* or epidemic* or occurrence or case or cases).ti,ab,hw. | 2831 |
| 27 | ((decay* or declin* or decreas* or impair* or duration or fail* or fall* or "go* down" or lack or lacks or lacked or lacking or lose or loses or losing or loss or lost or low or lower* or lesser* or poor or poorer* or "adversely affect*" or reduc*) adj5 (protect* or effectiv* or immunity or "immune respon*" or antibod* or sero*)).ab,ti,hw. and (infected or infection* or contracted or acquired or acquisition or caught or got* or transmi* or spread* or outbreak* or cluster* or communicab* or contagious* or epidemic* or occurrence or case or cases).hw,ti. | 34252 |
| 28 | 25 or 26 or 27 | 34783 |
| 29 | (1 or 4) and (15 or 24 or 28) and (5 and 6) | 226 |
| 30 | 2 and (15 or 28) and (5 and 6) | 373 |
| 31 | 29 or 30 | 406 |
| 32 | limit 31 to yr="2019 -Current" | 313 |
| 33 | limit 32 to (english or french) | 306 |
| 34 | 12 not 33 | 803 |

BIOSIS

BIOSIS Previews <2021 Week 01 to 2021 Week 41>

|  | Searches | Results |
| --- | --- | --- |
| 1 | (ncov or BNT162* or "mrna-1273" or "Ad5-nCoV vaccine" or "BNT162 vaccine" or "ChAdOx1 COVID-19 vaccine" or "Covid-19 aAPC vaccine" or "lentiviral minigene vaccine of COVID-19 coronavirus" or "mRNA-1273 vaccine" or "recombinant SARS-CoV-2 vaccine NVX-cov2373" or "SARS-CoV-2 inactivated vaccines").rn. or (BNT162* or Comirnaty or CoronaVac or Covishield or Tozinameran or "mrna-1273*" or ChAdOx1* or AZD1222 or "Ad26.COV2.S" or "JNJ-78436735" or Novavax or NVX-CoV2373 or Ad5-nCoV or aAPC or "lentiviral minigene" or vaxzevria).ti,tw. | 400 |
| 2 | ("Severe acute respiratory syndrome coronavirus 2" or "coronavirus disease 2019" or "COVID-19" or "SARS-CoV-2" or ("2019 corona virus" or "2019 coronavirus" or "2019 ncov" or "corona virus 19" or "corona virus 2019" or "corona virus disease 19" or "corona virus disease 2019" or "corona virus epidemic*" or "corona virus outbreak*" or "corona virus pandemic*" or "corona virus response" or "coronavirus 19" or "coronavirus 2019" or "coronavirus disease 19" or "coronavirus disease 2019" or "coronavirus epidemic*" or "coronavirus outbreak*" or "coronavirus pandemic*" or "coronavirus response" or "covid 19" or "covid 2019" or "new corona virus" or "new coronavirus" or "novel corona virus" or "novel coronavirus" or "novel human coronavirus" or "sars coronavirus 2" or "sars cov 2" or "sars cov2" or "sars like coronavirus" or "severe acute respiratory syndrome corona virus 2" or "severe acute respiratory syndrome coronavirus 2" or "severe specific contagious pneumonia" or "wuhan corona virus" or "wuhan coronavirus" or ((pandemic* or novel or wuhan) adj3 (coronavirus* or "corona virus*" or betacoronavirus* or "beta coronavirus*" or "beta corona virus*" or pneumonia* or SARS or "severe acute respiratory syndrome")) or (pneumonia adj3 (coronavirus* or "corona virus*" or betacoronavirus* or "beta coronavirus*" or "beta corona virus*" or SARS or "severe acute respiratory syndrome")) or 2019ncov or covid or covid19 or covid2019 or ncov or sarscov2)).ti,hw,mc. or ("SARS-Cov-2 variant*" or VUI-202012-01 or "VOC-202012/01" or "B.1.1.7" or B117 or "B.1.351" or B1351 or "B.1.617" or B1617 or "P.1" or P1 or coronavirus* or "corona virus*" or betacoronavirus* or "beta coronavirus*" or "beta corona virus*" or pneumonia* or SARS or "severe acute respiratory syndrome").ti. or ((covid* or sars* or variant*) and (alpha or beta or gamma or delta or "variant* of concern" or "VOC")).ti. or ("2019 corona virus" or "2019 coronavirus" or "2019 ncov" or "corona virus 19" or "corona virus 2019" or "corona virus disease 19" or "corona virus disease 2019" or "corona virus epidemic*" or "corona virus outbreak*" or "corona virus pandemic*" or "corona virus response" or "coronavirus 19" or "coronavirus 2019" or "coronavirus disease 19" or "coronavirus disease 2019" or "coronavirus epidemic*" or "coronavirus outbreak*" or "coronavirus pandemic*" or "coronavirus response" or "covid 19" or "covid 2019" or "new corona virus" or "new coronavirus" or "novel corona virus" or "novel coronavirus" or "novel human coronavirus" or "sars coronavirus 2" or "sars cov 2" or "sars cov2" or "sars like coronavirus" or "severe acute respiratory syndrome corona virus 2" or "severe acute respiratory syndrome coronavirus 2" or "severe specific contagious pneumonia" or "wuhan corona virus" or "wuhan coronavirus" or ((pandemic* or novel or wuhan) adj3 (coronavirus* or "corona virus*" or betacoronavirus* or "beta coronavirus*" or "beta corona virus*" or pneumonia* or SARS or "severe acute respiratory syndrome")) or (pneumonia adj3 (coronavirus* or "corona virus*" or betacoronavirus* or "beta coronavirus*" or "beta corona virus*" or SARS or "severe acute respiratory syndrome")) or 2019ncov or covid or covid19 or covid2019 or ncov or sarscov2).ab. /freq=2 | 41503 |
| 3 | (vaccin* or immunis* or immuniz* or innoculat* or inoculat* or jab* or shot* or injection* or nonimmunis* or nonimmuniz* or postvaccin* or postimmunis* or postimmuniz* or reimmunis* or reimmuniz* or revaccinat* or unimmunis* or unimmuniz* or ((one or single two or double or first or second or partial* or full) adj2 (dose* or injection* or injected or injecting)) or single-dose* or double-dose* or BNT162* or Comirnaty or CoronaVac or Covishield or Tozinameran or "mrna-1273*" or ChAdOx1* or AZD1222 or "Ad26.COV2.S" or "JNJ-78436735" or Novavax or NVX-CoV2373 or Ad5-nCoV or aAPC or "lentiviral minigene" or vaxzevria).ti,ab,hw,tw,mc. | 67637 |
| 4 | 2 and 3 | 6024 |
| 5 | (antibody or antibodies or antibody-positiv* or antibody-negativ* or "complement activation" or ((humoral or humoural or adaptive or antibody-mediated) adj3 (respon* or immun*)) or immunity or "immune system*" or "immune marker*" or ((immun* or sero*) adj3 respon*) or immunolog* or immunomodulat* or B-lymphocyte* or "B-cell*").ti,ab,hw,tw,mc. or (immun* or immunoglobulin* or IgA or IgD or IgE or IgG or IgM or sero* or sera*).ti,hw. | 288794 |
| 6 | ("correlat* of protection" or "correlat* with protection" or "immune correlat*" or "correlat* of immunity" or "correlat* of neutraliz*" or "correlat* of neutralis*" or "measur* of protection" or "correlat* of infectivity" or "protective correlat*" or ((protective or protection) adj5 (level* or threshold* or marker* or limit* or amount* or mean* or baseline or quantif*)) or ((correlat* or marker* or threshold* or indicat* or measur*) adj5 (immun* or protect*)) or ((antibody or antibodies or titer* or titre*) adj3 (correlat* or threshold*)) or (protect* adj5 (immunity or antibod* or titer* or titre*)) or ((neutraliz* or neutralis*) adj2 (level* or amount* or limit* or mean* or baseline or quantif* or titer* or titre*)) or (predict* adj3 (immun* or protect*)) or "protective immunity" or "surrogate endpoint*").ti,ab,hw,tw,mc. or (protection or protective or protected or ((antibody or antibodies) adj2 (status or level*))).ti. | 24995 |
| 7 | 5 and 6 | 15896 |
| 8 | (1 or 4) and 7 | 578 |
| 9 | 2 and 7 | 1229 |
| 10 | 8 or 9 | 1230 |
| 11 | limit 10 to yr="2019 -Current" | 1191 |
| 12 | limit 11 to (english or french) | 1178 |
| 13 | (((previous* or prior or past or post or postcovid* or post-covid* or postsars* or post-sars* or after or following or recover* or convalescen* or "after infection") adj3 infect*) or re-infect* or reinfect* or "after first infection").ti,ab,hw,tw,mc. | 9662 |
| 14 | ((vaccin* or immunis* or immuniz*) adj3 fail*).ti,ab,hw,tw,mc. | 227 |
| 15 | (((breakthrough* or "break through*") adj5 (vaccin* or infection*)) or "breakthrough rate*").ti,ab,hw,tw,mc. | 125 |
| 16 | (("second dose" or "two doses" or "fully vaccinated") adj5 (infected or infection* or contracted or acquired or acquisition or caught or got* or transmi* or spread* or outbreak* or cluster* or communicab* or contagious* or epidemic* or occurrence or case or cases)).ti,ab,hw,tw,mc. | 40 |
| 17 | ((postvaccin* or postimmuniz* or postimmunis* or "post-vaccin*" or "post-immuniz*" or "post-immunis*") adj5 (infected or infection* or contracted or acquired or acquisition or caught or got* or transmi* or spread* or outbreak* or cluster* or communicab* or contagious* or epidemic* or occurrence or case or cases)).ti,ab,hw,tw,mc. | 59 |
| 18 | (((receiv* or recipient* or complet* or already or post or previously or after or following or "subsequent to" or since or had or despite or "in spite of") adj5 (immunis* or immuniz* or inoculat* or innoculat* or vaccin* or revaccinat* or reimmuniz* or reimmunis* or dose or doses) adj5 (infected or infection* or contracted or acquired or acquisition or caught or got* or transmi* or spread* or outbreak* or cluster* or communicab* or contagious* or epidemic* or occurrence or case or cases)) or ((infected or infection* or contracted or acquired or acquisition or caught or got* or transmi* or spread* or outbreak* or cluster* or communicab* or contagious* or epidemic* or occurrence or case or cases) adj5 (receiv* or recipient* or complet* or already or post or previously or after or following or "subsequent to" or since or had or despite or "in spite of") adj5 (immunis* or immuniz* or inoculat* or innoculat* or vaccin* or revaccinat* or reimmuniz* or reimmunis* or dose or doses))).ti,ab,hw,tw,mc. | 1131 |
| 19 | ((vaccinated or immunized or immunised) adj5 (infected or infection* or contracted or acquired or acquisition or caught or got* or transmi* or spread* or outbreak* or cluster* or communicab* or contagious* or epidemic* or occurrence or case or cases)).ti,ab,hw,tw,mc. | 512 |
| 20 | ((vaccinated or immunized or immunised) adj10 (person or persons or people or man or men or woman or women or adult or adults or patient or patients or individual or individuals or responder or responders or practitioner* or staff or personnel or worker* or employee* or provider* or technician* or clinician* or doctor* or nurse or nurses or paramedic* or physician* or hospitalist* or pharmacist*) adj10 (infected or infection* or contracted or acquired or acquisition or caught or got* or transmi* or spread* or outbreak* or cluster* or communicab* or contagious* or epidemic* or occurrence or case or cases)).ti,ab,hw,tw,mc. | 204 |
| 21 | ((escape or escapes or escaped or escaping or evade or evades or evaded or evading or evasion) adj5 ("Ad26.COV2.S" or AZD1222 or BNT162* or ChAdOx1* or Comirnaty or immunis* or immuniz* or "JNJ-78436735" or "mrna-1273*" or Novavax or NVX-CoV2373 or PittCoVacc* or postvaccin* or postimmunis* or postimmuniz* or reimmunis* or reimmuniz* or revacciat* or tozinameran or vaccin*)).ti,ab,hw,tw,mc. | 132 |
| 22 | 14 or 15 or 16 or 17 or 18 or 19 or 20 or 21 | 2046 |
| 23 | (wane or wanes or waned or waning).ti,hw,tw. | 445 |
| 24 | ((decay* or declin* or decreas* or impair* or duration or fail* or fall* or "go* down" or lack or lacks or lacked or lacking or lose or loses or losing or loss or lost or low or lower* or lesser* or poor or poorer* or "adversely affect*" or reduc*) adj5 (protect* or effectiv* or immunity or "immune respon*" or antibod* or sero*)).ti,hw,tw. and (infected or infection* or contracted or acquired or acquisition or caught or got* or transmi* or spread* or outbreak* or cluster* or communicab* or contagious* or epidemic* or occurrence or case or cases).ti,ab,hw,tw,mc. | 9420 |
| 25 | ((decay* or declin* or decreas* or impair* or duration or fail* or fall* or "go* down" or lack or lacks or lacked or lacking or lose or loses or losing or loss or lost or low or lower* or lesser* or poor or poorer* or "adversely affect*" or reduc*) adj5 (protect* or effectiv* or immunity or "immune respon*" or antibod* or sero*)).ti,ab,hw,tw,mc. and (infected or infection* or contracted or acquired or acquisition or caught or got* or transmi* or spread* or outbreak* or cluster* or communicab* or contagious* or epidemic* or occurrence or case or cases).hw,ti,tw. | 9420 |
| 26 | 23 or 24 or 25 | 9779 |
| 27 | (1 or 4) and (13 or 22 or 26) and (5 and 6) | 202 |
| 28 | 2 and (13 or 26) and (5 and 6) | 366 |
| 29 | 27 or 28 | 379 |
| 30 | limit 29 to yr="2019 -Current" | 363 |
| 31 | limit 30 to (english or french) | 359 |
| 32 | 12 not 31 | 819 |

scopus

| # | Queries | Results |
| --- | --- | --- |
| #1 | TITLE-ABS ( "Ad5-nCoV vaccine"  OR  "BNT162 vaccine"  OR  "ChAdOx1 COVID-19 vaccine"  OR  "Covid-19 aAPC vaccine"  OR  "lentiviral minigene vaccine of COVID-19 cORonavirus"  OR  "mRNA-1273 vaccine"  OR  "recombinant SARS-CoV-2 vaccine NVX-cov2373"  OR  "SARS-CoV-2 inactivated vaccines" )  OR  TITLE-ABS ( bnt162*  OR  comirnaty  OR  coronavac  OR  covishield  OR  tozinameran  OR  "mrna-1273*"  OR  chadox1*  OR  azd1222  OR  "Ad26.COV2.S"  OR  "JNJ-78436735"  OR  novavax  OR  nvx-cov2373  OR  ad5-ncov  OR  aapc  OR  "lentiviral minigene"  OR  vaxzevria ) | 1,643 |
| #2 | ( TITLE-ABS ( "Severe acute respiratory syndrome coronavirus 2" OR "coronavirus disease 2019" OR "COVID-19" OR "SARS-CoV-2" ) ) OR ( TITLE-ABS ( "2019 corona virus" OR "2019 coronavirus" OR "2019 ncov" OR "corona virus 19" OR "corona virus 2019" OR "corona virus disease 19" OR "corona virus disease 2019" OR "corona virus epidemic*" OR "corona virus outbreak*" OR "corona virus pandemic*" OR "corona virus response" OR "coronavirus 19" OR "coronavirus 2019" OR "coronavirus disease 19" OR "coronavirus disease 2019" OR "coronavirus epidemic*" OR "coronavirus outbreak*" OR "coronavirus pandemic*" OR "coronavirus response" OR "covid 19" OR "covid 2019" OR "new corona virus" OR "new coronavirus" OR "novel corona virus" OR "novel coronavirus" OR "novel human coronavirus" OR "sars coronavirus 2" OR "sars cov 2" OR "sars cov2" OR "sars like coronavirus" OR "severe acute respiratory syndrome corona virus 2" OR "severe acute respiratory syndrome coronavirus 2" OR "severe specific contagious pneumonia" OR "wuhan corona virus" OR "wuhan coronavirus" OR ( ( pandemic* OR novel OR wuhan ) W/3 ( coronavirus* OR "corona virus*" OR betacoronavirus* OR "beta coronavirus*" OR "beta corona virus*" OR pneumonia* OR sars OR "severe acute respiratory syndrome" ) ) OR ( pneumonia W/3 ( coronavirus* OR "corona virus*" OR betacoronavirus* OR "beta coronavirus*" OR "beta corona virus*" OR sars OR "severe acute respiratory syndrome" ) ) OR 2019ncov OR covid OR covid19 OR covid2019 OR ncov OR sarscov2 ) ) OR ( TITLE ( "SARS-Cov-2 variant*" OR vui-202012-01 OR "VOC-202012/01" OR "B.1.1.7" OR b117 OR "B.1.351" OR b1351 OR "B.1.617" OR b1617 OR "P.1" OR p1 OR coronavirus* OR "corona virus*" OR betacoronavirus* OR "beta coronavirus*" OR "beta corona virus*" OR pneumonia* OR sars OR "severe acute respiratory syndrome" ) OR TITLE ( ( covid* OR sars* OR variant* ) AND ( alpha OR beta OR gamma OR delta OR "variant* of concern" OR "VOC" ) ) ) | 346,981 |
| #3 | TITLE-ABS ( ( vaccin* OR immunis* OR immuniz* OR innoculat* OR inoculat* OR jab* OR shot* OR injection* OR nonimmunis* OR nonimmuniz* OR postvaccin* OR postimmunis* OR postimmuniz* OR reimmunis* OR reimmuniz* OR revaccinat* OR unimmunis* OR unimmuniz* OR ( ( one OR single OR two OR double OR first OR second OR partial* OR full ) W/2 ( dose* OR injection* OR injected OR injecting ) ) OR single-dose* OR double-dose* OR bnt162* OR comirnaty OR coronavac OR covishield OR tozinameran OR "mrna-1273*" OR chadox1* OR azd1222 OR "Ad26.COV2.S" OR "JNJ-78436735" OR novavax OR nvx-cov2373 OR ad5-ncov OR aapc OR "lentiviral minigene" OR vaxzevria ) ) | 2,005,732 |
| #4 | #2 AND #3 | 25,288 |
| #5 | TITLE-ABS ( antibody OR antibodies OR antibody-positiv* OR antibody-negativ* OR "complement activation" OR ( ( humoral OR humoural OR adaptive OR antibody-mediated ) W/3 ( respon* OR immun* ) ) OR immunity OR "immune system*" OR "immune marker*" OR ( ( immun* OR sero* ) W/3 respon* ) OR immunolog* OR immunomodulat* OR b-lymphocyte* OR "B-cell*" ) OR TITLE ( immun* OR immunoglobulin* OR iga OR igd OR ige OR igg OR igm OR sero* OR sera* ) | 2,431,187 |
| #6 | TITLE-ABS ( "correlat* of protection" OR "correlat* with protection" OR "immune correlat*" OR "correlat* of immunity" OR "correlat* of neutraliz*" OR "correlat* of neutralis*" OR "measur* of protection" OR "correlat* of infectivity" OR "protective correlat*" OR ( ( protective OR protection ) W/5 ( level* OR threshold* OR marker* OR limit* OR amount* OR mean* OR baseline OR quantif* ) ) OR ( ( correlat* OR marker* OR threshold* OR indicat* OR measur* ) W/5 ( immun* OR protect* ) ) OR ( ( antibody OR antibodies OR titer* OR titre* ) W/3 ( correlat* OR threshold* ) ) OR ( protect* W/5 ( immunity OR antibod* OR titer* OR titre* ) ) OR ( ( neutraliz* OR neutralis* ) W/2 ( level* OR amount* OR limit* OR mean* OR baseline OR quantif* OR titer* OR titre* ) ) OR ( predict* W/3 ( immun* OR protect* ) ) OR "protective immunity" OR "surrogate endpoint*" ) OR TITLE ( protection OR protective OR protected OR ( ( antibody OR antibodies ) W/2 ( status OR level* ) ) ) | 678,074 |
| #7 | #5 AND #6 | 200,136 |
| #8 | (#1 OR #4) AND #7 | 1,855 |
| #9 | #2 AND #7 | 3,822 |
| #10 | #8 OR #9 | 3,847 |
| #11 | LANGUAGE(English) OR LANGUAGE(French) AND PUBYEAR > 2018 | 8,905,122 |
| #12 | #10 AND #11 | 2,219 |
| #13 | #12 AND NOT INDEX (MEDLINE) | 692 |

**LITERATURE SEARCH**

09/05/2021

COVID-19 antibody correlates of protection

Preprints

**Request prepared by:**	Library Services

**Contact information:**

Search results reporting

Databases searched

| Database | Date searched | Records | Duplicates removed by database | Remaining |
| --- | --- | --- | --- | --- |
| NIH iSearch COVID-19 Portfolio | 09/05/2021 |  | n/a | 512 |

Records totals

| Records source | Records |
| --- | --- |
| Records identified through database searching | 512 |
| Duplicates removed (manual de-duping) | 38 |
| Total records after duplicates removed | 474 |

Search strategies

National Institutes of Health iSearch COVID-19 Portfolio (Preprints) <https://icite.od.nih.gov/covid19/search/>

| # | Searches | Results |
| --- | --- | --- |
| 1 | (title:antibody OR title:antibodies OR title:antibody-response OR title:humoral OR title:humoural OR title:titer* OR title:titre* OR title:immune OR title:immunity OR title:"sero* response"~3 OR title:immunolog* OR title:immunomodulat* OR title:immunoglobulin* OR title:B-cell OR title:B-lymphocyte OR title:neutraliz* OR title:neutralis*) AND (title:protect* OR title:threshold* OR title:correlat*) AND PubTypes:"Preprint" | 188 |
| 2 | (title:immun* correlat* OR title:correlat* neutraliz* OR title:correlat* neutralis* OR title:"measur* of protection" OR title:"correlat* of infectivity" OR title:correlat* protect* OR title:protect* correlat* OR title:protect* threshold* OR title:protect* immunity OR title:predict* immun*) AND (title:antibody OR title:antibodies OR title:antibody-respons* OR title:humoral OR title:humoural OR title:titer* OR title:neutraliz* OR title:neutralis*) AND PubTypes:"Preprint" | 220 |
| 3 | (title:reinfect* OR title:re-infect* OR title:"after first infection" OR title:"postcovid* infect*"~5 OR title:"post-covid infect*"~5 OR title:postsars* OR title:"reocurr* infect*"~2) AND (title:protect* OR title:immun* OR title:sero* OR title:sera* OR title:antibod* OR title:titer* OR title:titre* OR title:neutraliz* OR title:neutralis* OR title:humoral OR title:humoural OR title:lymphocyte* OR title:B-cell* OR title:correlate* OR title:threshold* OR title:infectivity)  AND PubTypes:"Preprint" | 23 |
| 4 | (title:previous* OR title:prior OR title:past OR title:post OR title:after OR title:following OR title:recover* OR title:convalescen*) AND (title:infect*) AND (title:reinfect* OR title:re-infect*) AND PubTypes:"Preprint" | 7 |
| 5 | (title:postcovid OR title:post-covid OR title:postsars OR title:post-sars) AND (title:infect* OR title:reinfect* OR title:re-infect*) AND PubTypes:"Preprint" | 6 |
| 6 | (title:breakthrough* infection* OR title:"break through* infection*"~5 OR title:breakthrough* rate* OR title:breakthrough* vaccin* OR title:"break through* vaccin*"~5) AND (title:protect* OR title:immun* OR title:sero* OR title:sera* OR title:antibod* OR title:titer* OR title:titre* OR title:neutraliz* OR title:neutralis* OR title:humoral OR title:humoural OR title:lymphocyte* OR title:B-cell* OR title:correlate* OR title:threshold* OR title:infectivity) AND PubTypes:"Preprint" | 9 |
| 7 | (title:"one dose" OR title:"single vaccin*" OR title:"two dose*" OR title:"double dose*" OR title:"double vaccin*" OR title:"second dose" OR title:"two doses" OR title:"fully vaccinated" OR title:"partial* vaccin*" OR title:vaccinated OR title:immunized OR title:immunised OR title:postvaccin* OR title:postimmuniz* OR title:postimmunis* OR title:post-vaccin* OR title:post-immuniz* OR title:post-immunis*) AND (title:infect* OR title:re-infect* OR title:reinfect* OR title:"after first infection" OR title:contract* OR title:transmi* OR title:spread* OR title:outbreak* OR title:case* OR title:cluster* OR title:communicab* OR title:contagious*) AND PubTypes:"Preprint" | 35 |
| 8 | (title:escap* OR title:evad* OR title:evasion) AND (title:"Ad26.COV2.S" OR title:AZD1222 OR title:BNT162* OR title:ChAdOx1* OR title:Comirnaty OR title:immunis* OR title:immuniz* OR title:"JNJ-78436735" OR title:"mrna-1273*" OR title:Novavax OR title:NVX-CoV2373 OR title:PittCoVacc* OR title:postvaccin* OR title:postimmunis* OR title:postimmuniz* OR title:reimmunis* OR title:reimmuniz* OR title:revacciat* OR title:tozinameran OR title:vaccin*) AND PubTypes:"Preprint" | 24 |

Search results

Results attached as a CSV Excel file

**LI**

**LITERATURE SEARCH**

ERATURE SEARCH

10/05/2021

COVID-19 antibody correlates of protection

Preprints (supplemental search)

**Request prepared by:**	Library Services

**Contact information:**

Search results reporting

Databases searched

| Database | Date searched | Records | Duplicates removed by database | Remaining |
| --- | --- | --- | --- | --- |
| NIH iSearch COVID-19 Portfolio | 10/05/2021 |  | n/a | 194 |

Records totals

| Records source | Records |
| --- | --- |
| Records identified through database searching | 194 |
| Duplicates removed (manual de-duping) | 55 |
| Total records after duplicates removed | 139 |

Search strategies

National Institutes of Health iSearch COVID-19 Portfolio (Preprints) <https://icite.od.nih.gov/covid19/search/>

| # | Searches | Results |
| --- | --- | --- |
| 1 | (title:correlat* of protect*) AND PubTypes:"Preprint" | 10 |
| 2 | (abstract:"correlate of protection") OR (abstract:"correlates of protection") AND PubTypes:"Preprint" | 51 |
| 3 | (supplemental text:"correlate of protection") OR (supplemental text:"correlates of protection") AND PubTypes:"Preprint" | 20 |
| 4 | (title:correlate* of immun*) OR (title:correlate* of neutraliz*) AND PubTypes:"Preprint" | 18 |
| 5 | (title:protect* neutraliz*) OR (title:protect* immune*) OR (title:protect* threshold*) OR (title:neutraliz* threshold*) OR (title:antibod* threshold*) OR (title:titer* threshold*) OR (title:protect* level*) OR (title:optimal protection) OR (title:protect* correlate*) OR (title:immune* correlat*) AND PubTypes:"Preprint" | 95 |

Search results

Results attached as a CSV Excel file

**LITERATURE SEARCH**

12/31/2021

COVID-19 antibody correlates of protection

Preprints search update (original search run 2021 Oct 5^th^)

**Request prepared by:**	Library Services

**Contact information:**

Search results reporting

Databases searched

| Database | Date searched | Records | Duplicates removed by database | Remaining |
| --- | --- | --- | --- | --- |
| NIH iSearch COVID-19 Portfolio | 12/31/2021 | 27 | n/a | 27 |

Records totals

| Records source | Records |
| --- | --- |
| Records identified through database searching | 27 |
| Duplicates removed (manual de-duping) | 7 |
| Total records after duplicates removed | 20 |

Search strategies

National Institutes of Health iSearch COVID-19 Portfolio (Preprints) <https://icite.od.nih.gov/covid19/search/>

| # | Searches | Results |
| --- | --- | --- |
| 1 | (title:correlat* of protect*) AND PubTypes:"Preprint" AND Publication Date: 2021-10-06-* | 2 |
| 2 | (abstract:"correlate of protection") OR (abstract:"correlates of protection") AND PubTypes:"Preprint" AND Publication Date: 2021-10-06-* | 11 |
| 3 | (supplemental text:"correlate of protection") OR (supplemental text:"correlates of protection") AND PubTypes:"Preprint" AND Publication Date: 2021-10-06-* | 2 |
| 4 | (title:correlate* of immun*) OR (title:correlate* of neutraliz*) AND PubTypes:"Preprint" AND Publication Date: 2021-10-06-* | 1 |
| 5 | (title:protect* neutraliz*) OR (title:protect* immune*) OR (title:protect* threshold*) OR (title:neutraliz* threshold*) OR (title:antibod* threshold*) OR (title:titer* threshold*) OR (title:protect* level*) OR (title:optimal protection) OR (title:protect* correlate*) OR (title:immune* correlat*) AND PubTypes:"Preprint" AND Publication Date: 2021-10-06-* | 11 |

Search results

Results attached as a CSV Excel file

**LITERATURE SEARCH**

12/31/2021

COVID-19 antibody correlates of protection

Preprints search update (original search run 2021 Sept 3^rd^)

**Request prepared by:**	Library Services

**Contact information:**

Search results reporting

Databases searched

| Database | Date searched | Records | Duplicates removed by database | Remaining |
| --- | --- | --- | --- | --- |
| NIH iSearch COVID-19 Portfolio | 12/31/2021 | 161 | n/a | 161 |

Records totals

| Records source | Records |
| --- | --- |
| Records identified through database searching | 161 |
| Duplicates removed (manual de-duping) | 11 |
| Total records after duplicates removed | 150 |

Search strategies

National Institutes of Health iSearch COVID-19 Portfolio (Preprints) <https://icite.od.nih.gov/covid19/search/>

| # | Searches | Results |
| --- | --- | --- |
| 1 | (title:antibody OR title:antibodies OR title:antibody-response OR title:humoral OR title:humoural OR title:titer* OR title:titre* OR title:immune OR title:immunity OR title:"sero* response"~3 OR title:immunolog* OR title:immunomodulat* OR title:immunoglobulin* OR title:B-cell OR title:B-lymphocyte OR title:neutraliz* OR title:neutralis*) AND (title:protect* OR title:threshold* OR title:correlat*) AND PubTypes:"Preprint" AND Publication Date: 2021-09-04-* | 28 |
| 2 | (title:immun* correlat* OR title:correlat* neutraliz* OR title:correlat* neutralis* OR title:"measur* of protection" OR title:"correlat* of infectivity" OR title:correlat* protect* OR title:protect* correlat* OR title:protect* threshold* OR title:protect* immunity OR title:predict* immun*) AND (title:antibody OR title:antibodies OR title:antibody-respons* OR title:humoral OR title:humoural OR title:titer* OR title:neutraliz* OR title:neutralis*) AND PubTypes:"Preprint" AND Publication Date: 2021-09-04-* | 68 |
| 3 | (title:reinfect* OR title:re-infect* OR title:"after first infection" OR title:"postcovid* infect*"~5 OR title:"post-covid infect*"~5 OR title:postsars* OR title:"reocurr* infect*"~2) AND (title:protect* OR title:immun* OR title:sero* OR title:sera* OR title:antibod* OR title:titer* OR title:titre* OR title:neutraliz* OR title:neutralis* OR title:humoral OR title:humoural OR title:lymphocyte* OR title:B-cell* OR title:correlate* OR title:threshold* OR title:infectivity)  AND PubTypes:"Preprint" AND Publication Date: 2021-09-04-* | 3 |
| 4 | (title:previous* OR title:prior OR title:past OR title:post OR title:after OR title:following OR title:recover* OR title:convalescen*) AND (title:infect*) AND (title:reinfect* OR title:re-infect*) AND PubTypes:"Preprint" AND Publication Date: 2021-09-04-* | 0 |
| 5 | (title:postcovid OR title:post-covid OR title:postsars OR title:post-sars) AND (title:infect* OR title:reinfect* OR title:re-infect*) AND PubTypes:"Preprint" AND Publication Date: 2021-09-04-* | 2 |
| 6 | (title:breakthrough* infection* OR title:"break through* infection*"~5 OR title:breakthrough* rate* OR title:breakthrough* vaccin* OR title:"break through* vaccin*"~5) AND (title:protect* OR title:immun* OR title:sero* OR title:sera* OR title:antibod* OR title:titer* OR title:titre* OR title:neutraliz* OR title:neutralis* OR title:humoral OR title:humoural OR title:lymphocyte* OR title:B-cell* OR title:correlate* OR title:threshold* OR title:infectivity) AND PubTypes:"Preprint" AND Publication Date: 2021-09-04-* | 21 |
| 7 | (title:"one dose" OR title:"single vaccin*" OR title:"two dose*" OR title:"double dose*" OR title:"double vaccin*" OR title:"second dose" OR title:"two doses" OR title:"fully vaccinated" OR title:"partial* vaccin*" OR title:vaccinated OR title:immunized OR title:immunised OR title:postvaccin* OR title:postimmuniz* OR title:postimmunis* OR title:post-vaccin* OR title:post-immuniz* OR title:post-immunis*) AND (title:infect* OR title:re-infect* OR title:reinfect* OR title:"after first infection" OR title:contract* OR title:transmi* OR title:spread* OR title:outbreak* OR title:case* OR title:cluster* OR title:communicab* OR title:contagious*) AND PubTypes:"Preprint" AND Publication Date: 2021-09-04-* | 27 |
| 8 | (title:escap* OR title:evad* OR title:evasion) AND (title:"Ad26.COV2.S" OR title:AZD1222 OR title:BNT162* OR title:ChAdOx1* OR title:Comirnaty OR title:immunis* OR title:immuniz* OR title:"JNJ-78436735" OR title:"mrna-1273*" OR title:Novavax OR title:NVX-CoV2373 OR title:PittCoVacc* OR title:postvaccin* OR title:postimmunis* OR title:postimmuniz* OR title:reimmunis* OR title:reimmuniz* OR title:revacciat* OR title:tozinameran OR title:vaccin*) AND PubTypes:"Preprint" AND Publication Date: 2021-09-04-* | 12 |

Search results

Results attached as a CSV Excel file

**LITERATURE SEARCH**

12/31/2021

COVID-19 antibody correlates of protection (revised) – Search update: 2021 Dec 31

Request prepared by: Library Services

Contact information:

Search results reporting

Databases Searched

| Database | Date searched | Records | Duplicates removed by database | Remaining |
| --- | --- | --- | --- | --- |
| MEDLINE (I) | 12/31/2021 | 960 | 0 | 960 |
| MEDLINE (II) | 12/31/2021 | 438 | 0 | 438 |

Results Totals

| Records source | Records |
| --- | --- |
| Records identified through database searching | 1398 |
| Duplicates removed by database | 0 |
| Duplicates removed by bibliographic management software | 409 |
| Total records after duplicates removed | 989 |

Search strategies

MEDLINE

Ovid MEDLINE(R) ALL <1946 to December 30, 2021>

| # | Searches | Results |
| --- | --- | --- |
| 1 | COVID-19 Vaccines/ or (ncov or BNT162* or "mrna-1273").rn,nm,os,ox,ps,px,rs,rx. or ("Ad5-nCoV vaccine" or "BNT162 vaccine" or "ChAdOx1 COVID-19 vaccine" or "Covid-19 aAPC vaccine" or "lentiviral minigene vaccine of COVID-19 coronavirus" or "mRNA-1273 vaccine" or "recombinant SARS-CoV-2 vaccine NVX-cov2373" or "SARS-CoV-2 inactivated vaccines").rn. or (BNT162* or Comirnaty or CoronaVac or Covishield or Tozinameran or "mrna-1273*" or ChAdOx1* or AZD1222 or "Ad26.COV2.S" or "JNJ-78436735" or Novavax or NVX-CoV2373 or Ad5-nCoV or aAPC or "lentiviral minigene" or vaxzevria).ti,kf,kw. | 8487 |
| 2 | COVID-19/ or SARS-CoV-2/ or "SARS-CoV-2 variants".os. or "COVID-19 breakthrough infections".rs. or ("COVID-19" or "SARS-CoV-2").nm,os,ox,ps,px,rs,rs. or (Pandemics/ and (Coronavirus Infections/ or Betacoronavirus/ or Spike Glycoprotein, Coronavirus/ or Pneumonia, Viral/)) or exp COVID-19 Testing/ or COVID-19 Serological Testing/ or ("2019 corona virus" or "2019 coronavirus" or "2019 ncov" or "corona virus 19" or "corona virus 2019" or "corona virus disease 19" or "corona virus disease 2019" or "corona virus epidemic*" or "corona virus outbreak*" or "corona virus pandemic*" or "corona virus response" or "coronavirus 19" or "coronavirus 2019" or "coronavirus disease 19" or "coronavirus disease 2019" or "coronavirus epidemic*" or "coronavirus outbreak*" or "coronavirus pandemic*" or "coronavirus response" or "covid 19" or "covid 2019" or "new corona virus" or "new coronavirus" or "novel corona virus" or "novel coronavirus" or "novel human coronavirus" or "sars coronavirus 2" or "sars cov 2" or "sars cov2" or "sars like coronavirus" or "severe acute respiratory syndrome corona virus 2" or "severe acute respiratory syndrome coronavirus 2" or "severe specific contagious pneumonia" or "wuhan corona virus" or "wuhan coronavirus" or ((pandemic* or novel or wuhan or delta) adj3 (coronavirus* or "corona virus*" or betacoronavirus* or "beta coronavirus*" or "beta corona virus*" or pneumonia* or SARS or "severe acute respiratory syndrome")) or (pneumonia adj3 (coronavirus* or "corona virus*" or betacoronavirus* or "beta coronavirus*" or "beta corona virus*" or SARS or "severe acute respiratory syndrome")) or 2019ncov or covid or covid19 or covid2019 or ncov or sarscov2).kf,kw,ti. or (coronavirus* or "corona virus*" or betacoronavirus* or "beta coronavirus*" or "beta corona virus*" or pneumonia* or SARS or "severe acute respiratory syndrome").ti. or ("2019 corona virus" or "2019 coronavirus" or "2019 ncov" or "corona virus 19" or "corona virus 2019" or "corona virus disease 19" or "corona virus disease 2019" or "corona virus epidemic*" or "corona virus outbreak*" or "corona virus pandemic*" or "corona virus response" or "coronavirus 19" or "coronavirus 2019" or "coronavirus disease 19" or "coronavirus disease 2019" or "coronavirus epidemic*" or "coronavirus outbreak*" or "coronavirus pandemic*" or "coronavirus response" or "covid 19" or "covid 2019" or "new corona virus" or "new coronavirus" or "novel corona virus" or "novel coronavirus" or "novel human coronavirus" or "sars coronavirus 2" or "sars cov 2" or "sars cov2" or "sars like coronavirus" or "severe acute respiratory syndrome corona virus 2" or "severe acute respiratory syndrome coronavirus 2" or "severe specific contagious pneumonia" or "wuhan corona virus" or "wuhan coronavirus" or ((pandemic* or novel or wuhan or delta) adj3 (coronavirus* or "corona virus*" or betacoronavirus* or "beta coronavirus*" or "beta corona virus*" or pneumonia* or SARS or "severe acute respiratory syndrome")) or (pneumonia adj3 (coronavirus* or "corona virus*" or betacoronavirus* or "beta coronavirus*" or "beta corona virus*" or SARS or "severe acute respiratory syndrome")) or 2019ncov or covid or covid19 or covid2019 or ncov or sarscov2).ab. /freq=2 | 294391 |
| 3 | (Omicron or "B.1.1.529" or "VOC-21NOV-01" or "B.1.1.529.1" or "B.1.1.529.2" or B11529*).ab,ti,kw,kf. | 501 |
| 4 | 2 or 3 | 294776 |
| 5 | Immunization Programs/ or Immunization Schedule/ or Immunization, Secondary/ or Immunization, Passive/ or Immunization/ or Immunogenicity, Vaccine/ or Immunotherapy, Active/ or Mass Vaccination/ or Vaccination Coverage/ or Vaccination/ or Vaccines, Attenuated/ or Vaccines, Combined/ or Vaccines, Synthetic/ or Vaccines/ or Viral Vaccines/ or COVID-19 Vaccines/ or vaccine*.rn. or (vaccin* or immunis* or immuniz* or innoculat* or inoculat* or jab* or shot* or injection* or nonimmunis* or nonimmuniz* or postvaccin* or postimmunis* or postimmuniz* or reimmunis* or reimmuniz* or revaccinat* or unimmunis* or unimmuniz* or ((one or single two or double or first or second or partial* or full) adj2 (dose* or injection* or injected or injecting)) or single-dose* or double-dose* or BNT162* or Comirnaty or CoronaVac or Covishield or Tozinameran or "mrna-1273*" or ChAdOx1* or AZD1222 or "Ad26.COV2.S" or "JNJ-78436735" or Novavax or NVX-CoV2373 or Ad5-nCoV or aAPC or "lentiviral minigene" or vaxzevria).ti,ab,kf,kw. | 1302150 |
| 6 | 4 and 5 | 33920 |
| 7 | Adaptive Immunity/ or Antibodies, Neutralizing/ or Antibodies, Viral/ or Antibody Affinity/ or Antibody Formation/ or Antibody Specificity/ or Antibodies, Monoclonal/ or Binding Sites, Antibody/ or exp Antibodies/ or exp Antigen-Antibody Reactions/ or Antibody-Dependent Enhancement/ or Antibody-Dependent Cell Cytotoxicity/ or Lymphocytes/ or exp B-Lymphocytes/ or B-Lymphocyte subsets/ or Lymphocyte Activation/ or Complement Activation/ or Th2 Cells/ or Immune System Phenomena/ or Immunity/ or Immunity, Active/ or Immunity, Humoral/ or Immunity, Mucosal/ or Immunologic Factors/ or Immunologic Surveillance/ or Immunity, Innate/ or Immunity, Heterologous/ or Immunity, Maternally-Acquired/ or Immune Sera/ or exp Immunoglobulins/ or Immunoglobulin M/ or Immunoglobulin G/ or exp Immunomodulation/ or Immunologic Memory/ or Models, Immunological/ or Cross Protection/im or Cross Reactions/im or Reinfection/im or Recurrence/im or "Severity of Illness Index"/im or Viral Load/im or Convalescence/im or Seroconversion/ or Seroepidemiologic Studies/ or Neutralization Tests/ or Enzyme-Linked Immunosorbent Assay/ or Protein Domains/im or Receptors, Virus/im or Spike Glycoprotein, Coronavirus/im or Coronavirus Nucleocapsid Proteins/im or *COVID-19 Vaccines/im or *COVID-19/im or *SARS-CoV-2/im or (antibody or antibodies or antibody-positiv* or antibody-negativ* or "complement activation" or ((humoral or humoural or adaptive or antibody-mediated) adj3 (respon* or immun*)) or immunity or "immune system*" or "immune marker*" or ((immun* or sero*) adj3 respon*) or immunolog* or immunomodulat* or B-lymphocyte* or "B-cell*").ti,ab,kf,kw. or (immun* or immunoglobulin* or IgA or IgD or IgE or IgG or IgM or sero* or sera*).ti,kf,kw. | 2833106 |
| 8 | ("correlat* of protection" or "correlat* with protection" or "immune correlat*" or "correlat* of immunity" or "correlat* of neutraliz*" or "correlat* of neutralis*" or "measur* of protection" or "correlat* of infectivity" or "protective correlat*" or ((protective or protection) adj5 (level* or threshold* or marker* or limit* or amount* or mean* or baseline or quantif*)) or ((correlat* or marker* or threshold* or indicat* or measur*) adj5 (immun* or protect*)) or ((antibody or antibodies or titer* or titre*) adj3 (correlat* or threshold*)) or (protect* adj5 (immunity or antibod* or titer* or titre*)) or ((neutraliz* or neutralis*) adj2 (level* or amount* or limit* or mean* or baseline or quantif* or titer* or titre*)) or (predict* adj3 (immun* or protect*)) or "protective immunity" or "surrogate endpoint*").ti,ab,kf,kw. or (protection or protective or protected or ((antibody or antibodies) adj2 (status or level*))).ti. | 378764 |
| 9 | (1 or 6) and 7 and 8 | 2767 |
| 10 | 4 and 7 and 8 | 4896 |
| 11 | 9 or 10 | 4902 |
| 12 | limit 11 to yr="2021 -Current" | 2686 |
| 13 | (202108* or 202109* or 202110* or 202111* or 202112* or 202201*).ez. or (202108* or 202109* or 202110* or 202111* or 202112* or 202201*).dt. or (202108* or 202109* or 202110* or 202111* or 202112* or 202201* or 2021 08* or 2021 09* or 2021 10* or 2021 11* or 2021 12* or 2022 01* or 2021 aug or 2021 sep or 2021 oct or 2021 nov or 2021 dec or 2022 jan).dp. | 802962 |
| 14 | 12 and 13 | 1419 |
| 15 | limit 14 to (english or french) | 1398 |
| 16 | Reinfection/ or Recurrence/ or Follow-Up Studies/ or ((Convalescence/ or "Severity of Illness Index"/ or Viral Load/ or Asymptomatic Infections/) and (re-infect* or reinfect*).ti,kf,kw.) | 838483 |
| 17 | (((previous* or prior or past or post or postcovid* or post-covid* or postsars* or post-sars* or after or following or recover*) adj2 infect*) or re-infect* or reinfect* or "after first infection" or "after infection").ti,ab,kf,kw. or (convalescence or convalescent).ti,kf,kw. | 139085 |
| 18 | 16 or 17 | 969761 |
| 19 | Treatment Failure/ or ((vaccin* or immunis* or immuniz*) adj3 fail*).ab,kf,kw,ti. | 39893 |
| 20 | (((breakthrough* or "break through*") adj5 (vaccin* or infection*)) or "breakthrough rate*").ab,kf,kw,ti. | 1599 |
| 21 | (("second dose" or "two doses" or "fully vaccinated") adj5 (infected or infection* or contracted or acquired or acquisition or caught or got* or transmi* or spread* or outbreak* or cluster* or communicab* or contagious* or epidemic* or occurrence or case or cases)).ab,kf,kw,ti. | 481 |
| 22 | ((postvaccin* or postimmuniz* or postimmunis* or "post-vaccin*" or "post-immuniz*" or "post-immunis*") adj5 (infected or infection* or contracted or acquired or acquisition or caught or got* or transmi* or spread* or outbreak* or cluster* or communicab* or contagious* or epidemic* or occurrence or case or cases)).ab,kf,kw,ti. | 677 |
| 23 | (((receiv* or recipient* or complet* or already or post or previously or after or following or "subsequent to" or since or had or despite or "in spite of") adj5 (immunis* or immuniz* or inoculat* or innoculat* or vaccin* or revaccinat* or reimmuniz* or reimmunis* or dose or doses) adj5 (infected or infection* or contracted or acquired or acquisition or caught or got* or transmi* or spread* or outbreak* or cluster* or communicab* or contagious* or epidemic* or occurrence or case or cases)) or ((infected or infection* or contracted or acquired or acquisition or caught or got* or transmi* or spread* or outbreak* or cluster* or communicab* or contagious* or epidemic* or occurrence or case or cases) adj5 (receiv* or recipient* or complet* or already or post or previously or after or following or "subsequent to" or since or had or despite or "in spite of") adj5 (immunis* or immuniz* or inoculat* or innoculat* or vaccin* or revaccinat* or reimmuniz* or reimmunis* or dose or doses))).ab,kf,kw,ti. | 19960 |
| 24 | ((vaccinated or immunized or immunised) adj5 (infected or infection* or contracted or acquired or acquisition or caught or got* or transmi* or spread* or outbreak* or cluster* or communicab* or contagious* or epidemic* or occurrence or case or cases)).ab,kf,kw,ti. | 8870 |
| 25 | ((vaccinated or immunized or immunised) adj10 (person or persons or people or man or men or woman or women or adult or adults or patient or patients or individual or individuals or responder or responders or practitioner* or staff or personnel or worker* or employee* or provider* or technician* or clinician* or doctor* or nurse or nurses or paramedic* or physician* or hospitalist* or pharmacist*) adj10 (infected or infection* or contracted or acquired or acquisition or caught or got* or transmi* or spread* or outbreak* or cluster* or communicab* or contagious* or epidemic* or occurrence or case or cases)).ab,kf,kw,ti. | 3016 |
| 26 | ((escape or escapes or escaped or escaping or evade or evades or evaded or evading or evasion) adj5 ("Ad26.COV2.S" or AZD1222 or BNT162* or ChAdOx1* or Comirnaty or immunis* or immuniz* or "JNJ-78436735" or "mrna-1273*" or Novavax or NVX-CoV2373 or PittCoVacc* or postvaccin* or postimmunis* or postimmuniz* or reimmunis* or reimmuniz* or revacciat* or tozinameran or vaccin*)).ab,kf,kw,ti. | 1081 |
| 27 | 19 or 20 or 21 or 22 or 23 or 24 or 25 or 26 | 69543 |
| 28 | (wane or wanes or waned or waning).kf,kw,ti. | 606 |
| 29 | ((decay* or declin* or decreas* or impair* or duration or fail* or fall* or "go* down" or lack or lacks or lacked or lacking or lose or loses or losing or loss or lost or low or lower* or lesser* or poor or poorer* or "adversely affect*" or reduc*) adj5 (protect* or effectiv* or immunity or "immune respon*" or antibod* or sero*)).kf,kw,ti. and (infected or infection* or contracted or acquired or acquisition or caught or got* or transmi* or spread* or outbreak* or cluster* or communicab* or contagious* or epidemic* or occurrence or case or cases).ab,kf,kw,ti. | 6314 |
| 30 | ((decay* or declin* or decreas* or impair* or duration or fail* or fall* or "go* down" or lack or lacks or lacked or lacking or lose or loses or losing or loss or lost or low or lower* or lesser* or poor or poorer* or "adversely affect*" or reduc*) adj5 (protect* or effectiv* or immunity or "immune respon*" or antibod* or sero*)).ab,kf,kw,ti. and (infected or infection* or contracted or acquired or acquisition or caught or got* or transmi* or spread* or outbreak* or cluster* or communicab* or contagious* or epidemic* or occurrence or case or cases).kf,kw,ti. | 30642 |
| 31 | 28 or 29 or 30 | 35386 |
| 32 | (1 or 6) and (18 or 27 or 31) and (7 and 8) | 852 |
| 33 | 4 and (18 or 31) and (7 and 8) | 1144 |
| 34 | 32 or 33 | 1319 |
| 35 | limit 34 to yr="2021 -Current" | 840 |
| 36 | (202108* or 202109* or 202110* or 202111* or 202112* or 202201*).ez. or (202108* or 202109* or 202110* or 202111* or 202112* or 202201*).dt. or (202108* or 202109* or 202110* or 202111* or 202112* or 202201* or 2021 08* or 2021 09* or 2021 10* or 2021 11* or 2021 12* or 2022 01* or 2021 aug or 2021 sep or 2021 oct or 2021 nov or 2021 dec or 2022 jan).dp. | 802962 |
| 37 | 35 and 36 | 444 |
| 38 | limit 37 to (english or french) | 438 |
| 39 | 15 not 38 | 960 |

**LITERATURE SEARCH**

01/04/2022

COVID-19 antibody correlates of protection (revised): Supplementary Databases

Search update - 2022 Jan 04 (Part 1)

Request prepared by: Library Services

Contact information:

Search results reporting

Databases Searched

| Database | Date searched | Records | Duplicates removed by database | Remaining |
| --- | --- | --- | --- | --- |
| EMBASE | 01/04/2022 | 231 | 0 | 231 |
| GLOBAL HEALTH | 01/04/2022 | 424 | 0 | 424 |

Results Totals

| Records source | Records |
| --- | --- |
| Records identified through database searching | 655 |
| Duplicates removed by database | 0 |
| Duplicates removed by bibliographic management software | 521 |
| Total records after duplicates removed | 134 |

Search strategies

EMBASE

Embase <1974 to 2021 Week 52>

| # | Searches | Results |
| --- | --- | --- |
| 1 | SARS-CoV-2 vaccine/ or (ncov or BNT162* or "mrna-1273").rn,os,ox,px,rs. or ("Ad5-nCoV vaccine" or "BNT162 vaccine" or "ChAdOx1 COVID-19 vaccine" or "Covid-19 aAPC vaccine" or "lentiviral minigene vaccine of COVID-19 coronavirus" or "mRNA-1273 vaccine" or "recombinant SARS-CoV-2 vaccine NVX-cov2373" or "SARS-CoV-2 inactivated vaccines").rn. or (BNT162* or Comirnaty or CoronaVac or Covishield or Tozinameran or "mrna-1273*" or ChAdOx1* or AZD1222 or "Ad26.COV2.S" or "JNJ-78436735" or Novavax or NVX-CoV2373 or Ad5-nCoV or aAPC or "lentiviral minigene" or vaxzevria).ti,kf,kw. | 9147 |
| 2 | coronavirus disease 2019/ or Severe acute respiratory syndrome coronavirus 2/ or "SARS-CoV-2 variants".os. or "COVID-19 breakthrough infections".rs. or ("COVID-19" or "SARS-CoV-2").os,ox,px,rs,rs. or (pandemic/ and (Coronavirus infection/ or Betacoronavirus/ or coronavirus spike glycoprotein/ or virus pneumonia/)) or exp COVID-19 testing/ or COVID-19 serological testing/ or ("2019 corona virus" or "2019 coronavirus" or "2019 ncov" or "corona virus 19" or "corona virus 2019" or "corona virus disease 19" or "corona virus disease 2019" or "corona virus epidemic*" or "corona virus outbreak*" or "corona virus pandemic*" or "corona virus response" or "coronavirus 19" or "coronavirus 2019" or "coronavirus disease 19" or "coronavirus disease 2019" or "coronavirus epidemic*" or "coronavirus outbreak*" or "coronavirus pandemic*" or "coronavirus response" or "covid 19" or "covid 2019" or "new corona virus" or "new coronavirus" or "novel corona virus" or "novel coronavirus" or "novel human coronavirus" or "sars coronavirus 2" or "sars cov 2" or "sars cov2" or "sars like coronavirus" or "severe acute respiratory syndrome corona virus 2" or "severe acute respiratory syndrome coronavirus 2" or "severe specific contagious pneumonia" or "wuhan corona virus" or "wuhan coronavirus" or ((pandemic* or novel or wuhan or delta) adj3 (coronavirus* or "corona virus*" or betacoronavirus* or "beta coronavirus*" or "beta corona virus*" or pneumonia* or SARS or "severe acute respiratory syndrome")) or (pneumonia adj3 (coronavirus* or "corona virus*" or betacoronavirus* or "beta coronavirus*" or "beta corona virus*" or SARS or "severe acute respiratory syndrome")) or 2019ncov or covid or covid19 or covid2019 or ncov or sarscov2).kf,kw,ti. or (coronavirus* or "corona virus*" or betacoronavirus* or "beta coronavirus*" or "beta corona virus*" or pneumonia* or SARS or "severe acute respiratory syndrome").ti. or ("2019 corona virus" or "2019 coronavirus" or "2019 ncov" or "corona virus 19" or "corona virus 2019" or "corona virus disease 19" or "corona virus disease 2019" or "corona virus epidemic*" or "corona virus outbreak*" or "corona virus pandemic*" or "corona virus response" or "coronavirus 19" or "coronavirus 2019" or "coronavirus disease 19" or "coronavirus disease 2019" or "coronavirus epidemic*" or "coronavirus outbreak*" or "coronavirus pandemic*" or "coronavirus response" or "covid 19" or "covid 2019" or "new corona virus" or "new coronavirus" or "novel corona virus" or "novel coronavirus" or "novel human coronavirus" or "sars coronavirus 2" or "sars cov 2" or "sars cov2" or "sars like coronavirus" or "severe acute respiratory syndrome corona virus 2" or "severe acute respiratory syndrome coronavirus 2" or "severe specific contagious pneumonia" or "wuhan corona virus" or "wuhan coronavirus" or ((pandemic* or novel or wuhan or delta) adj3 (coronavirus* or "corona virus*" or betacoronavirus* or "beta coronavirus*" or "beta corona virus*" or pneumonia* or SARS or "severe acute respiratory syndrome")) or (pneumonia adj3 (coronavirus* or "corona virus*" or betacoronavirus* or "beta coronavirus*" or "beta corona virus*" or SARS or "severe acute respiratory syndrome")) or 2019ncov or covid or covid19 or covid2019 or ncov or sarscov2).ab. /freq=2 | 319718 |
| 3 | (Omicron or "B.1.1.529" or "VOC-21NOV-01" or "B.1.1.529.1" or "B.1.1.529.2" or B11529*).ab,ti,kw,kf. | 183 |
| 4 | 2 or 3 | 319871 |
| 5 | immunization/ or secondary immunization/ or vaccine immunogenicity/ or active immunotherapy/ or mass immunization/ or vaccination/ or vaccination coverage/ or vaccination refusal/ or live vaccine/ or vaccine/ or recombinant vaccine/ or virus vaccine/ or SARS-CoV-2 vaccine/ or (vaccin* or immunis* or immuniz* or innoculat* or inoculat* or jab* or shot* or injection* or nonimmunis* or nonimmuniz* or postvaccin* or postimmunis* or postimmuniz* or reimmunis* or reimmuniz* or revaccinat* or unimmunis* or unimmuniz* or ((one or single two or double or first or second or partial* or full) adj2 (dose* or injection* or injected or injecting)) or single-dose* or double-dose* or BNT162* or Comirnaty or CoronaVac or Covishield or Tozinameran or "mrna-1273*" or ChAdOx1* or AZD1222 or "Ad26.COV2.S" or "JNJ-78436735" or Novavax or NVX-CoV2373 or Ad5-nCoV or aAPC or "lentiviral minigene" or vaxzevria).ti,ab,kw. | 1597974 |
| 6 | 4 and 5 | 36945 |
| 7 | adaptive immunity/ or neutralizing antibody/ or virus antibody/ or antibody affinity/ or antibody production/ or antibody specificity/ or monoclonal antibody/ or antibody combining site/ or exp antibody/ or exp antigen antibody reaction/ or antibody dependent enhancement/ or antibody dependent cellular cytotoxicity/ or antibody response/ or lymphocyte/ or exp B lymphocyte/ or B lymphocyte subpopulation/ or lymphocyte activation/ or complement activation/ or Th2 cell/ or immunity/ or active immunization/ or humoral immunity/ or mucosal immunity/ or immunologic factor/ or immunosurveillance/ or innate immunity/ or heterologous immunity/ or passive immunization/ or antiserum/ or exp immunoglobulin/ or immunoglobulin G/ or immunological memory/ or biological model/ or cross protection/ or cross reaction/ or reinfection/ or recurrent disease/ or "severity of illness index"/ or virus load/ or convalescence/ or seroconversion/ or seroepidemiology/ or serodiagnosis/ or enzyme linked immunosorbent assay/ or protein domain/ or virus receptor/ or coronavirus spike glycoprotein/ or nucleocapsid protein/ or (antibody or antibodies or antibody-positiv* or antibody-negativ* or "complement activation" or ((humoral or humoural or adaptive or antibody-mediated) adj3 (respon* or immun*)) or immunity or "immune system*" or "immune marker*" or ((immun* or sero*) adj3 respon*) or immunolog* or immunomodulat* or B-lymphocyte* or "B-cell*").ti,kw. or (antibody or antibodies or antibody-positiv* or antibody-negativ* or "complement activation" or ((humoral or humoural or adaptive or antibody-mediated) adj3 (respon* or immun*)) or immunity or "immune system*" or "immune marker*" or ((immun* or sero*) adj3 respon*) or immunolog* or immunomodulat* or B-lymphocyte* or "B-cell*").ab. /freq=2 or (immun* or immunoglobulin* or IgA or IgD or IgE or IgG or IgM or sero* or sera*).ti,kw. | 3993818 |
| 8 | ("correlat* of protection" or "correlat* with protection" or "immune correlat*" or "correlat* of immunity" or "correlat* of neutraliz*" or "correlat* of neutralis*" or "measur* of protection" or "correlat* of infectivity" or "protective correlat*" or ((protective or protection) adj5 (level* or threshold* or marker* or limit* or amount* or mean* or baseline or quantif*)) or ((correlat* or marker* or threshold* or indicat* or measur*) adj5 (immun* or protect*)) or ((antibody or antibodies or titer* or titre*) adj3 (correlat* or threshold*)) or (protect* adj5 (immunity or antibod* or titer* or titre*)) or ((neutraliz* or neutralis*) adj2 (level* or amount* or limit* or mean* or baseline or quantif* or titer* or titre*)) or (predict* adj3 (immun* or protect*)) or "protective immunity" or "surrogate endpoint*").ti,ab,kw. or (protection or protective or protected or ((antibody or antibodies) adj2 (status or level*))).ti. | 484212 |
| 9 | 7 and 8 | 228405 |
| 10 | reinfection/ or recurrent disease/ or ((follow up/ or convalescence/ or "severity of illness index"/ or virus load/ or asymptomatic infection/) and (re-infect* or reinfect*).ti,kw.) | 204048 |
| 11 | (((previous* or prior or past or post or postcovid* or post-covid* or postsars* or post-sars* or after or following or recover* or convalescen* or "after infection") adj2 infect*) or re-infect* or reinfect* or "after first infection").ti,ab,kw. | 171733 |
| 12 | 10 or 11 | 366022 |
| 13 | treatment failure/ or ((vaccin* or immunis* or immuniz*) adj3 fail*).ab,kw,ti. | 146623 |
| 14 | (((breakthrough* or "break through*") adj5 (vaccin* or infection*)) or "breakthrough rate*").ab,kw,ti. | 2130 |
| 15 | (("second dose" or "two doses" or "fully vaccinated") adj5 (infected or infection* or contracted or acquired or acquisition or caught or got* or transmi* or spread* or outbreak* or cluster* or communicab* or contagious* or epidemic* or occurrence or case or cases)).ab,kw,ti. | 581 |
| 16 | ((postvaccin* or postimmuniz* or postimmunis* or "post-vaccin*" or "post-immuniz*" or "post-immunis*") adj5 (infected or infection* or contracted or acquired or acquisition or caught or got* or transmi* or spread* or outbreak* or cluster* or communicab* or contagious* or epidemic* or occurrence or case or cases)).ab,kw,ti. | 787 |
| 17 | (((receiv* or recipient* or complet* or already or post or previously or after or following or "subsequent to" or since or had or despite or "in spite of") adj5 (immunis* or immuniz* or inoculat* or innoculat* or vaccin* or revaccinat* or reimmuniz* or reimmunis* or dose or doses) adj5 (infected or infection* or contracted or acquired or acquisition or caught or got* or transmi* or spread* or outbreak* or cluster* or communicab* or contagious* or epidemic* or occurrence or case or cases)) or ((infected or infection* or contracted or acquired or acquisition or caught or got* or transmi* or spread* or outbreak* or cluster* or communicab* or contagious* or epidemic* or occurrence or case or cases) adj5 (receiv* or recipient* or complet* or already or post or previously or after or following or "subsequent to" or since or had or despite or "in spite of") adj5 (immunis* or immuniz* or inoculat* or innoculat* or vaccin* or revaccinat* or reimmuniz* or reimmunis* or dose or doses))).ab,kw,ti. | 24910 |
| 18 | ((vaccinated or immunized or immunised) adj5 (infected or infection* or contracted or acquired or acquisition or caught or got* or transmi* or spread* or outbreak* or cluster* or communicab* or contagious* or epidemic* or occurrence or case or cases)).ab,kw,ti. | 9738 |
| 19 | ((vaccinated or immunized or immunised) adj10 (person or persons or people or man or men or woman or women or adult or adults or patient or patients or individual or individuals or responder or responders or practitioner* or staff or personnel or worker* or employee* or provider* or technician* or clinician* or doctor* or nurse or nurses or paramedic* or physician* or hospitalist* or pharmacist*) adj10 (infected or infection* or contracted or acquired or acquisition or caught or got* or transmi* or spread* or outbreak* or cluster* or communicab* or contagious* or epidemic* or occurrence or case or cases)).ab,kw,ti. | 3761 |
| 20 | ((escape or escapes or escaped or escaping or evade or evades or evaded or evading or evasion) adj5 ("Ad26.COV2.S" or AZD1222 or BNT162* or ChAdOx1* or Comirnaty or immunis* or immuniz* or "JNJ-78436735" or "mrna-1273*" or Novavax or NVX-CoV2373 or PittCoVacc* or postvaccin* or postimmunis* or postimmuniz* or reimmunis* or reimmuniz* or revacciat* or tozinameran or vaccin*)).ab,kw,ti. | 1213 |
| 21 | 13 or 14 or 15 or 16 or 17 or 18 or 19 or 20 | 182506 |
| 22 | (wane or wanes or waned or waning).kw,ti. | 578 |
| 23 | ((decay* or declin* or decreas* or impair* or duration or fail* or fall* or "go* down" or lack or lacks or lacked or lacking or lose or loses or losing or loss or lost or low or lower* or lesser* or poor or poorer* or "adversely affect*" or reduc*) adj5 (protect* or effectiv* or immunity or "immune respon*" or antibod* or sero*)).kw,ti. and (infected or infection* or contracted or acquired or acquisition or caught or got* or transmi* or spread* or outbreak* or cluster* or communicab* or contagious* or epidemic* or occurrence or case or cases).ab,kw,ti. | 8094 |
| 24 | ((decay* or declin* or decreas* or impair* or duration or fail* or fall* or "go* down" or lack or lacks or lacked or lacking or lose or loses or losing or loss or lost or low or lower* or lesser* or poor or poorer* or "adversely affect*" or reduc*) adj5 (protect* or effectiv* or immunity or "immune respon*" or antibod* or sero*)).ab,kw,ti. and (infected or infection* or contracted or acquired or acquisition or caught or got* or transmi* or spread* or outbreak* or cluster* or communicab* or contagious* or epidemic* or occurrence or case or cases).kw,ti. | 36564 |
| 25 | 22 or 23 or 24 | 42852 |
| 26 | (1 or 6) and (12 or 21 or 25) and 9 | 700 |
| 27 | 4 and (12 or 25) and 9 | 962 |
| 28 | 26 or 27 | 1133 |
| 29 | limit 28 to conference abstract | 170 |
| 30 | 28 not 29 | 963 |
| 31 | 30 not (editorial or letter or note).pt. | 934 |
| 32 | limit 31 to yr="2021 -Current" | 552 |
| 33 | (202108* or 202109* or 202110* or 202111* or 202112* or 202201*).dd. or (summer 2021 or fall 2021 or autumn 2021 or winter 2021 or aug* 2021 or sep* 2021 or oct* 2021 or nov* 2021 or dec* 2021 or jan* 2022).dp. | 647758 |
| 34 | 32 and 33 | 232 |
| 35 | limit 34 to (english or french) | 231 |

Global Health

Global Health <1973 to 2021 Week 50>

|  | Searches | Results |
| --- | --- | --- |
| 1 | ((vaccines/ or immunization/ or vaccination/ or synthetic vaccines/ or combined vaccines/ or vaccine development/ or candidate vaccines/ or messenger RNA/) and (ncov or BNT162* or "mrna-1273").ti,ab,bt,hw,id,od,sh.) or (BNT162* or Comirnaty or CoronaVac or Covishield or Tozinameran or "mrna-1273*" or ChAdOx1* or AZD1222 or "Ad26.COV2.S" or "JNJ-78436735" or Novavax or NVX-CoV2373 or Ad5-nCoV or aAPC or "lentiviral minigene" or vaxzevria).ti,ab,bt,hw,id,od,sh. | 910 |
| 2 | ("Severe acute respiratory syndrome coronavirus 2" or "coronavirus disease 2019" or "COVID-19" or "SARS-CoV-2").ti,ab,bt,hw,id,od,sh. or ("2019 corona virus" or "2019 coronavirus" or "2019 ncov" or "corona virus 19" or "corona virus 2019" or "corona virus disease 19" or "corona virus disease 2019" or "corona virus epidemic*" or "corona virus outbreak*" or "corona virus pandemic*" or "corona virus response" or "coronavirus 19" or "coronavirus 2019" or "coronavirus disease 19" or "coronavirus disease 2019" or "coronavirus epidemic*" or "coronavirus outbreak*" or "coronavirus pandemic*" or "coronavirus response" or "covid 19" or "covid 2019" or "new corona virus" or "new coronavirus" or "novel corona virus" or "novel coronavirus" or "novel human coronavirus" or "sars coronavirus 2" or "sars cov 2" or "sars cov2" or "sars like coronavirus" or "severe acute respiratory syndrome corona virus 2" or "severe acute respiratory syndrome coronavirus 2" or "severe specific contagious pneumonia" or "wuhan corona virus" or "wuhan coronavirus" or ((pandemic* or novel or wuhan) adj3 (coronavirus* or "corona virus*" or betacoronavirus* or "beta coronavirus*" or "beta corona virus*" or pneumonia* or SARS or "severe acute respiratory syndrome")) or (pneumonia adj3 (coronavirus* or "corona virus*" or betacoronavirus* or "beta coronavirus*" or "beta corona virus*" or SARS or "severe acute respiratory syndrome")) or 2019ncov or covid or covid19 or covid2019 or ncov or sarscov2).ti,hw. or ("SARS-Cov-2 variant*" or VUI-202012-01 or "VOC-202012/01" or "B.1.1.7" or B117 or "B.1.351" or B1351 or "B.1.617" or B1617 or "P.1" or P1 or coronavirus* or "corona virus*" or betacoronavirus* or "beta coronavirus*" or "beta corona virus*" or pneumonia* or SARS or "severe acute respiratory syndrome").ti. or ((covid* or sars* or variant*) and (alpha or beta or gamma or delta or "variant* of concern" or "VOC")).ti. or ("2019 corona virus" or "2019 coronavirus" or "2019 ncov" or "corona virus 19" or "corona virus 2019" or "corona virus disease 19" or "corona virus disease 2019" or "corona virus epidemic*" or "corona virus outbreak*" or "corona virus pandemic*" or "corona virus response" or "coronavirus 19" or "coronavirus 2019" or "coronavirus disease 19" or "coronavirus disease 2019" or "coronavirus epidemic*" or "coronavirus outbreak*" or "coronavirus pandemic*" or "coronavirus response" or "covid 19" or "covid 2019" or "new corona virus" or "new coronavirus" or "novel corona virus" or "novel coronavirus" or "novel human coronavirus" or "sars coronavirus 2" or "sars cov 2" or "sars cov2" or "sars like coronavirus" or "severe acute respiratory syndrome corona virus 2" or "severe acute respiratory syndrome coronavirus 2" or "severe specific contagious pneumonia" or "wuhan corona virus" or "wuhan coronavirus" or ((pandemic* or novel or wuhan) adj3 (coronavirus* or "corona virus*" or betacoronavirus* or "beta coronavirus*" or "beta corona virus*" or pneumonia* or SARS or "severe acute respiratory syndrome")) or (pneumonia adj3 (coronavirus* or "corona virus*" or betacoronavirus* or "beta coronavirus*" or "beta corona virus*" or SARS or "severe acute respiratory syndrome")) or 2019ncov or covid or covid19 or covid2019 or ncov or sarscov2).ab. /freq=2 | 88811 |
| 3 | (Omicron or "B.1.1.529" or "VOC-21NOV-01" or "B.1.1.529.1" or "B.1.1.529.2" or B11529*).ti,ab,bt,hw,id,od,sh. | 0 |
| 4 | 2 or 3 | 88811 |
| 5 | vaccines/ or immunization programmes/ or immunization/ or vaccination/ or passive immunization/ or synthetic vaccines/ or combined vaccines/ or vaccine development/ or candidate vaccines/ or (vaccin* or immunis* or immuniz* or innoculat* or inoculat* or jab* or shot* or injection* or nonimmunis* or nonimmuniz* or postvaccin* or postimmunis* or postimmuniz* or reimmunis* or reimmuniz* or revaccinat* or unimmunis* or unimmuniz* or ((one or single two or double or first or second or partial* or full) adj2 (dose* or injection* or injected or injecting)) or single-dose* or double-dose* or BNT162* or Comirnaty or CoronaVac or Covishield or Tozinameran or "mrna-1273*" or ChAdOx1* or AZD1222 or "Ad26.COV2.S" or "JNJ-78436735" or Novavax or NVX-CoV2373 or Ad5-nCoV or aAPC or "lentiviral minigene" or vaxzevria).ti,ab,hw. | 282459 |
| 6 | 4 and 5 | 10999 |
| 7 | neutralizing antibodies/ or monoclonal antibodies/ or exp antibodies/ or antibody formation/ or antibody testing/ or antigen antibody reactions/ or humoral immunity/ or immune serum/ or immunity/ or immune response/ or immune system/ or cross immunity/ or cross reaction/ or passive immunity/ or immune competence/ or immunological factors/ or immunology/ or antibody dependent cellular cytotoxicity/ or lymphocytes/ or B lymphocytes/ or lymphocyte transformation/ or complement activation/ or Th2 lymphocytes/ or exp immunoglobulins/ or reinfection/ or viral load/ or seroconversion/ or seroprevalence/ or neutralization tests/ or virus neutralization/ or ELISA/ or (antibody or antibodies or antibody-positiv* or antibody-negativ* or "complement activation" or ((humoral or humoural or adaptive or antibody-mediated) adj3 (respon* or immun*)) or immunity or "immune system*" or "immune marker*" or ((immun* or sero*) adj3 respon*) or immunolog* or immunomodulat* or B-lymphocyte* or "B-cell*").ti,ab,bt,hw,id,od,sh. or (immun* or immunoglobulin* or IgA or IgD or IgE or IgG or IgM or sero* or sera*).ti,hw. | 684729 |
| 8 | ("correlat* of protection" or "correlat* with protection" or "immune correlat*" or "correlat* of immunity" or "correlat* of neutraliz*" or "correlat* of neutralis*" or "measur* of protection" or "correlat* of infectivity" or "protective correlat*" or ((protective or protection) adj5 (level* or threshold* or marker* or limit* or amount* or mean* or baseline or quantif*)) or ((correlat* or marker* or threshold* or indicat* or measur*) adj5 (immun* or protect*)) or ((antibody or antibodies or titer* or titre*) adj3 (correlat* or threshold*)) or (protect* adj5 (immunity or antibod* or titer* or titre*)) or ((neutraliz* or neutralis*) adj2 (level* or amount* or limit* or mean* or baseline or quantif* or titer* or titre*)) or (predict* adj3 (immun* or protect*)) or "protective immunity" or "surrogate endpoint*").ti,ab,bt,hw,id,od,sh. or (protection or protective or protected or ((antibody or antibodies) adj2 (status or level*))).ti. | 112171 |
| 9 | 7 and 8 | 72310 |
| 10 | reinfection/ or ((viral load/ or asymptomatic infections/) and (re-infect* or reinfect*).ti,hw.) | 2547 |
| 11 | (((previous* or prior or past or post or postcovid* or post-covid* or postsars* or post-sars* or after or following or recover* or convalescen* or "after infection") adj3 infect*) or re-infect* or reinfect* or "after first infection").ti,ab,bt,hw,id,od,sh. | 61095 |
| 12 | 10 or 11 | 61095 |
| 13 | treatment failure/ or ((vaccin* or immunis* or immuniz*) adj3 fail*).ti,ab,bt,hw,id,od,sh. | 10512 |
| 14 | (((breakthrough* or "break through*") adj5 (vaccin* or infection*)) or "breakthrough rate*").ti,ab,bt,hw,id,od,sh. | 771 |
| 15 | (("second dose" or "two doses" or "fully vaccinated") adj5 (infected or infection* or contracted or acquired or acquisition or caught or got* or transmi* or spread* or outbreak* or cluster* or communicab* or contagious* or epidemic* or occurrence or case or cases)).ti,ab,bt,hw,id,od,sh. | 220 |
| 16 | ((postvaccin* or postimmuniz* or postimmunis* or "post-vaccin*" or "post-immuniz*" or "post-immunis*") adj5 (infected or infection* or contracted or acquired or acquisition or caught or got* or transmi* or spread* or outbreak* or cluster* or communicab* or contagious* or epidemic* or occurrence or case or cases)).ti,ab,bt,hw,id,od,sh. | 276 |
| 17 | (((receiv* or recipient* or complet* or already or post or previously or after or following or "subsequent to" or since or had or despite or "in spite of") adj5 (immunis* or immuniz* or inoculat* or innoculat* or vaccin* or revaccinat* or reimmuniz* or reimmunis* or dose or doses) adj5 (infected or infection* or contracted or acquired or acquisition or caught or got* or transmi* or spread* or outbreak* or cluster* or communicab* or contagious* or epidemic* or occurrence or case or cases)) or ((infected or infection* or contracted or acquired or acquisition or caught or got* or transmi* or spread* or outbreak* or cluster* or communicab* or contagious* or epidemic* or occurrence or case or cases) adj5 (receiv* or recipient* or complet* or already or post or previously or after or following or "subsequent to" or since or had or despite or "in spite of") adj5 (immunis* or immuniz* or inoculat* or innoculat* or vaccin* or revaccinat* or reimmuniz* or reimmunis* or dose or doses))).ti,ab,bt,hw,id,od,sh. | 8273 |
| 18 | ((vaccinated or immunized or immunised) adj5 (infected or infection* or contracted or acquired or acquisition or caught or got* or transmi* or spread* or outbreak* or cluster* or communicab* or contagious* or epidemic* or occurrence or case or cases)).ti,ab,bt,hw,id,od,sh. | 4036 |
| 19 | ((vaccinated or immunized or immunised) adj10 (person or persons or people or man or men or woman or women or adult or adults or patient or patients or individual or individuals or responder or responders or practitioner* or staff or personnel or worker* or employee* or provider* or technician* or clinician* or doctor* or nurse or nurses or paramedic* or physician* or hospitalist* or pharmacist*) adj10 (infected or infection* or contracted or acquired or acquisition or caught or got* or transmi* or spread* or outbreak* or cluster* or communicab* or contagious* or epidemic* or occurrence or case or cases)).ti,ab,bt,hw,id,od,sh. | 1732 |
| 20 | ((escape or escapes or escaped or escaping or evade or evades or evaded or evading or evasion) adj5 ("Ad26.COV2.S" or AZD1222 or BNT162* or ChAdOx1* or Comirnaty or immunis* or immuniz* or "JNJ-78436735" or "mrna-1273*" or Novavax or NVX-CoV2373 or PittCoVacc* or postvaccin* or postimmunis* or postimmuniz* or reimmunis* or reimmuniz* or revacciat* or tozinameran or vaccin*)).ti,ab,bt,hw,id,od,sh. | 427 |
| 21 | 13 or 14 or 15 or 16 or 17 or 18 or 19 or 20 | 23037 |
| 22 | (wane or wanes or waned or waning).ti,bt,hw,id,od,sh. | 122 |
| 23 | ((decay* or declin* or decreas* or impair* or duration or fail* or fall* or "go* down" or lack or lacks or lacked or lacking or lose or loses or losing or loss or lost or low or lower* or lesser* or poor or poorer* or "adversely affect*" or reduc*) adj5 (protect* or effectiv* or immunity or "immune respon*" or antibod* or sero*)).ti,hw. and (infected or infection* or contracted or acquired or acquisition or caught or got* or transmi* or spread* or outbreak* or cluster* or communicab* or contagious* or epidemic* or occurrence or case or cases).ti,ab,bt,hw,id,od,sh. | 2939 |
| 24 | ((decay* or declin* or decreas* or impair* or duration or fail* or fall* or "go* down" or lack or lacks or lacked or lacking or lose or loses or losing or loss or lost or low or lower* or lesser* or poor or poorer* or "adversely affect*" or reduc*) adj5 (protect* or effectiv* or immunity or "immune respon*" or antibod* or sero*)).ti,ab,bt,hw,id,od,sh. and (infected or infection* or contracted or acquired or acquisition or caught or got* or transmi* or spread* or outbreak* or cluster* or communicab* or contagious* or epidemic* or occurrence or case or cases).ti,bt,hw,id,od,sh. | 36265 |
| 25 | 22 or 23 or 24 | 36807 |
| 26 | (1 or 6) and (12 or 21 or 25) and 9 | 428 |
| 27 | 4 and (12 or 25) and 9 | 677 |
| 28 | 26 or 27 | 747 |
| 29 | limit 28 to yr="2021 -Current" | 429 |
| 30 | limit 29 to (english or french) | 424 |

**LITERATURE SEARCH**

01/05/2022

COVID-19 antibody correlates of protection (revised): Supplementary Databases

Search update - 2022 Jan 05 (Part 2)

Request prepared by: Library Services

Contact information:

Search results reporting

Databases Searched

| Database | Date searched | Records | Duplicates removed by database | Remaining |
| --- | --- | --- | --- | --- |
| BIOSIS Previews | 01/05/2022 | 368 | 0 | 368 |
| SCOPUS | 01/05/2022 | 866 | 0 | 866 |

Results Totals

| Records source | Records |
| --- | --- |
| Records identified through database searching | 1,234 |
| Duplicates removed by database | 0 |
| Duplicates removed by bibliographic management software | 784 |
| Total records after duplicates removed | 150 |

Search strategies

BIOSIS

BIOSIS Previews <2021>

| # | Searches | Results |
| --- | --- | --- |
| 1 | (ncov or BNT162* or "mrna-1273" or "Ad5-nCoV vaccine" or "BNT162 vaccine" or "ChAdOx1 COVID-19 vaccine" or "Covid-19 aAPC vaccine" or "lentiviral minigene vaccine of COVID-19 coronavirus" or "mRNA-1273 vaccine" or "recombinant SARS-CoV-2 vaccine NVX-cov2373" or "SARS-CoV-2 inactivated vaccines").rn. or (BNT162* or Comirnaty or CoronaVac or Covishield or Tozinameran or "mrna-1273*" or ChAdOx1* or AZD1222 or "Ad26.COV2.S" or "JNJ-78436735" or Novavax or NVX-CoV2373 or Ad5-nCoV or aAPC or "lentiviral minigene" or vaxzevria).ti,tw. | 715 |
| 2 | ("Severe acute respiratory syndrome coronavirus 2" or "coronavirus disease 2019" or "COVID-19" or "SARS-CoV-2" or ("2019 corona virus" or "2019 coronavirus" or "2019 ncov" or "corona virus 19" or "corona virus 2019" or "corona virus disease 19" or "corona virus disease 2019" or "corona virus epidemic*" or "corona virus outbreak*" or "corona virus pandemic*" or "corona virus response" or "coronavirus 19" or "coronavirus 2019" or "coronavirus disease 19" or "coronavirus disease 2019" or "coronavirus epidemic*" or "coronavirus outbreak*" or "coronavirus pandemic*" or "coronavirus response" or "covid 19" or "covid 2019" or "new corona virus" or "new coronavirus" or "novel corona virus" or "novel coronavirus" or "novel human coronavirus" or "sars coronavirus 2" or "sars cov 2" or "sars cov2" or "sars like coronavirus" or "severe acute respiratory syndrome corona virus 2" or "severe acute respiratory syndrome coronavirus 2" or "severe specific contagious pneumonia" or "wuhan corona virus" or "wuhan coronavirus" or ((pandemic* or novel or wuhan) adj3 (coronavirus* or "corona virus*" or betacoronavirus* or "beta coronavirus*" or "beta corona virus*" or pneumonia* or SARS or "severe acute respiratory syndrome")) or (pneumonia adj3 (coronavirus* or "corona virus*" or betacoronavirus* or "beta coronavirus*" or "beta corona virus*" or SARS or "severe acute respiratory syndrome")) or 2019ncov or covid or covid19 or covid2019 or ncov or sarscov2)).ti,hw,mc. or ("SARS-Cov-2 variant*" or VUI-202012-01 or "VOC-202012/01" or "B.1.1.7" or B117 or "B.1.351" or B1351 or "B.1.617" or B1617 or "P.1" or P1 or coronavirus* or "corona virus*" or betacoronavirus* or "beta coronavirus*" or "beta corona virus*" or pneumonia* or SARS or "severe acute respiratory syndrome").ti. or ((covid* or sars* or variant*) and (alpha or beta or gamma or delta or "variant* of concern" or "VOC")).ti. or ("2019 corona virus" or "2019 coronavirus" or "2019 ncov" or "corona virus 19" or "corona virus 2019" or "corona virus disease 19" or "corona virus disease 2019" or "corona virus epidemic*" or "corona virus outbreak*" or "corona virus pandemic*" or "corona virus response" or "coronavirus 19" or "coronavirus 2019" or "coronavirus disease 19" or "coronavirus disease 2019" or "coronavirus epidemic*" or "coronavirus outbreak*" or "coronavirus pandemic*" or "coronavirus response" or "covid 19" or "covid 2019" or "new corona virus" or "new coronavirus" or "novel corona virus" or "novel coronavirus" or "novel human coronavirus" or "sars coronavirus 2" or "sars cov 2" or "sars cov2" or "sars like coronavirus" or "severe acute respiratory syndrome corona virus 2" or "severe acute respiratory syndrome coronavirus 2" or "severe specific contagious pneumonia" or "wuhan corona virus" or "wuhan coronavirus" or ((pandemic* or novel or wuhan) adj3 (coronavirus* or "corona virus*" or betacoronavirus* or "beta coronavirus*" or "beta corona virus*" or pneumonia* or SARS or "severe acute respiratory syndrome")) or (pneumonia adj3 (coronavirus* or "corona virus*" or betacoronavirus* or "beta coronavirus*" or "beta corona virus*" or SARS or "severe acute respiratory syndrome")) or 2019ncov or covid or covid19 or covid2019 or ncov or sarscov2).ab. /freq=2 | 53052 |
| 3 | (Omicron or "B.1.1.529" or "VOC-21NOV-01" or "B.1.1.529.1" or "B.1.1.529.2" or B11529*).ab,ti,kw,hw,tw,mc. | 50 |
| 4 | 2 or 3 | 53100 |
| 5 | (vaccin* or immunis* or immuniz* or innoculat* or inoculat* or jab* or shot* or injection* or nonimmunis* or nonimmuniz* or postvaccin* or postimmunis* or postimmuniz* or reimmunis* or reimmuniz* or revaccinat* or unimmunis* or unimmuniz* or ((one or single two or double or first or second or partial* or full) adj2 (dose* or injection* or injected or injecting)) or single-dose* or double-dose* or BNT162* or Comirnaty or CoronaVac or Covishield or Tozinameran or "mrna-1273*" or ChAdOx1* or AZD1222 or "Ad26.COV2.S" or "JNJ-78436735" or Novavax or NVX-CoV2373 or Ad5-nCoV or aAPC or "lentiviral minigene" or vaxzevria).ti,ab,hw,tw,mc. | 90677 |
| 6 | 4 and 5 | 8270 |
| 7 | (antibody or antibodies or antibody-positiv* or antibody-negativ* or "complement activation" or ((humoral or humoural or adaptive or antibody-mediated) adj3 (respon* or immun*)) or immunity or "immune system*" or "immune marker*" or ((immun* or sero*) adj3 respon*) or immunolog* or immunomodulat* or B-lymphocyte* or "B-cell*").ti,ab,hw,tw,mc. or (immun* or immunoglobulin* or IgA or IgD or IgE or IgG or IgM or sero* or sera*).ti,hw. | 373129 |
| 8 | ("correlat* of protection" or "correlat* with protection" or "immune correlat*" or "correlat* of immunity" or "correlat* of neutraliz*" or "correlat* of neutralis*" or "measur* of protection" or "correlat* of infectivity" or "protective correlat*" or ((protective or protection) adj5 (level* or threshold* or marker* or limit* or amount* or mean* or baseline or quantif*)) or ((correlat* or marker* or threshold* or indicat* or measur*) adj5 (immun* or protect*)) or ((antibody or antibodies or titer* or titre*) adj3 (correlat* or threshold*)) or (protect* adj5 (immunity or antibod* or titer* or titre*)) or ((neutraliz* or neutralis*) adj2 (level* or amount* or limit* or mean* or baseline or quantif* or titer* or titre*)) or (predict* adj3 (immun* or protect*)) or "protective immunity" or "surrogate endpoint*").ti,ab,hw,tw,mc. or (protection or protective or protected or ((antibody or antibodies) adj2 (status or level*))).ti. | 33075 |
| 9 | 7 and 8 | 20510 |
| 10 | (((previous* or prior or past or post or postcovid* or post-covid* or postsars* or post-sars* or after or following or recover* or convalescen* or "after infection") adj3 infect*) or re-infect* or reinfect* or "after first infection").ti,ab,hw,tw,mc. | 13256 |
| 11 | ((vaccin* or immunis* or immuniz*) adj3 fail*).ti,ab,hw,tw,mc. | 303 |
| 12 | (((breakthrough* or "break through*") adj5 (vaccin* or infection*)) or "breakthrough rate*").ti,ab,hw,tw,mc. | 181 |
| 13 | (("second dose" or "two doses" or "fully vaccinated") adj5 (infected or infection* or contracted or acquired or acquisition or caught or got* or transmi* or spread* or outbreak* or cluster* or communicab* or contagious* or epidemic* or occurrence or case or cases)).ti,ab,hw,tw,mc. | 58 |
| 14 | ((postvaccin* or postimmuniz* or postimmunis* or "post-vaccin*" or "post-immuniz*" or "post-immunis*") adj5 (infected or infection* or contracted or acquired or acquisition or caught or got* or transmi* or spread* or outbreak* or cluster* or communicab* or contagious* or epidemic* or occurrence or case or cases)).ti,ab,hw,tw,mc. | 89 |
| 15 | (((receiv* or recipient* or complet* or already or post or previously or after or following or "subsequent to" or since or had or despite or "in spite of") adj5 (immunis* or immuniz* or inoculat* or innoculat* or vaccin* or revaccinat* or reimmuniz* or reimmunis* or dose or doses) adj5 (infected or infection* or contracted or acquired or acquisition or caught or got* or transmi* or spread* or outbreak* or cluster* or communicab* or contagious* or epidemic* or occurrence or case or cases)) or ((infected or infection* or contracted or acquired or acquisition or caught or got* or transmi* or spread* or outbreak* or cluster* or communicab* or contagious* or epidemic* or occurrence or case or cases) adj5 (receiv* or recipient* or complet* or already or post or previously or after or following or "subsequent to" or since or had or despite or "in spite of") adj5 (immunis* or immuniz* or inoculat* or innoculat* or vaccin* or revaccinat* or reimmuniz* or reimmunis* or dose or doses))).ti,ab,hw,tw,mc. | 1595 |
| 16 | ((vaccinated or immunized or immunised) adj5 (infected or infection* or contracted or acquired or acquisition or caught or got* or transmi* or spread* or outbreak* or cluster* or communicab* or contagious* or epidemic* or occurrence or case or cases)).ti,ab,hw,tw,mc. | 720 |
| 17 | ((vaccinated or immunized or immunised) adj10 (person or persons or people or man or men or woman or women or adult or adults or patient or patients or individual or individuals or responder or responders or practitioner* or staff or personnel or worker* or employee* or provider* or technician* or clinician* or doctor* or nurse or nurses or paramedic* or physician* or hospitalist* or pharmacist*) adj10 (infected or infection* or contracted or acquired or acquisition or caught or got* or transmi* or spread* or outbreak* or cluster* or communicab* or contagious* or epidemic* or occurrence or case or cases)).ti,ab,hw,tw,mc. | 282 |
| 18 | ((escape or escapes or escaped or escaping or evade or evades or evaded or evading or evasion) adj5 ("Ad26.COV2.S" or AZD1222 or BNT162* or ChAdOx1* or Comirnaty or immunis* or immuniz* or "JNJ-78436735" or "mrna-1273*" or Novavax or NVX-CoV2373 or PittCoVacc* or postvaccin* or postimmunis* or postimmuniz* or reimmunis* or reimmuniz* or revacciat* or tozinameran or vaccin*)).ti,ab,hw,tw,mc. | 180 |
| 19 | 10 or 11 or 12 or 13 or 14 or 15 or 16 or 17 or 18 | 15363 |
| 20 | (wane or wanes or waned or waning).ti,hw,tw. | 627 |
| 21 | ((decay* or declin* or decreas* or impair* or duration or fail* or fall* or "go* down" or lack or lacks or lacked or lacking or lose or loses or losing or loss or lost or low or lower* or lesser* or poor or poorer* or "adversely affect*" or reduc*) adj5 (protect* or effectiv* or immunity or "immune respon*" or antibod* or sero*)).ti,hw,tw. and (infected or infection* or contracted or acquired or acquisition or caught or got* or transmi* or spread* or outbreak* or cluster* or communicab* or contagious* or epidemic* or occurrence or case or cases).ti,ab,hw,tw,mc. | 12710 |
| 22 | ((decay* or declin* or decreas* or impair* or duration or fail* or fall* or "go* down" or lack or lacks or lacked or lacking or lose or loses or losing or loss or lost or low or lower* or lesser* or poor or poorer* or "adversely affect*" or reduc*) adj5 (protect* or effectiv* or immunity or "immune respon*" or antibod* or sero*)).ti,ab,hw,tw,mc. and (infected or infection* or contracted or acquired or acquisition or caught or got* or transmi* or spread* or outbreak* or cluster* or communicab* or contagious* or epidemic* or occurrence or case or cases).hw,ti,tw. | 12710 |
| 23 | 20 or 21 or 22 | 13204 |
| 24 | (1 or 6) and (19 or 23) and 9 | 295 |
| 25 | 4 and (10 or 23) and 9 | 475 |
| 26 | 24 or 25 | 502 |
| 27 | limit 26 to yr="2021 -Current" | 370 |
| 28 | limit 27 to (english or french) | 368 |

scopus

| # | Queries | Results |
| --- | --- | --- |
| #1 | TITLE-ABS ( "Ad5-nCoV vaccine"  OR  "BNT162 vaccine"  OR  "ChAdOx1 COVID-19 vaccine"  OR  "Covid-19 aAPC vaccine"  OR  "lentiviral minigene vaccine of COVID-19 cORonavirus"  OR  "mRNA-1273 vaccine"  OR  "recombinant SARS-CoV-2 vaccine NVX-cov2373"  OR  "SARS-CoV-2 inactivated vaccines" )  OR  TITLE-ABS ( bnt162*  OR  comirnaty  OR  coronavac  OR  covishield  OR  tozinameran  OR  "mrna-1273*"  OR  chadox1*  OR  azd1222  OR  "Ad26.COV2.S"  OR  "JNJ-78436735"  OR  novavax  OR  nvx-cov2373  OR  ad5-ncov  OR  aapc  OR  "lentiviral minigene"  OR  vaxzevria ) | 2,721 |
| #2 | ( TITLE-ABS ( "Severe acute respiratory syndrome coronavirus 2" OR "coronavirus disease 2019" OR "COVID-19" OR "SARS-CoV-2" ) ) OR ( TITLE-ABS ( "2019 corona virus" OR "2019 coronavirus" OR "2019 ncov" OR "corona virus 19" OR "corona virus 2019" OR "corona virus disease 19" OR "corona virus disease 2019" OR "corona virus epidemic*" OR "corona virus outbreak*" OR "corona virus pandemic*" OR "corona virus response" OR "coronavirus 19" OR "coronavirus 2019" OR "coronavirus disease 19" OR "coronavirus disease 2019" OR "coronavirus epidemic*" OR "coronavirus outbreak*" OR "coronavirus pandemic*" OR "coronavirus response" OR "covid 19" OR "covid 2019" OR "new corona virus" OR "new coronavirus" OR "novel corona virus" OR "novel coronavirus" OR "novel human coronavirus" OR "sars coronavirus 2" OR "sars cov 2" OR "sars cov2" OR "sars like coronavirus" OR "severe acute respiratory syndrome corona virus 2" OR "severe acute respiratory syndrome coronavirus 2" OR "severe specific contagious pneumonia" OR "wuhan corona virus" OR "wuhan coronavirus" OR ( ( pandemic* OR novel OR wuhan ) W/3 ( coronavirus* OR "corona virus*" OR betacoronavirus* OR "beta coronavirus*" OR "beta corona virus*" OR pneumonia* OR sars OR "severe acute respiratory syndrome" ) ) OR ( pneumonia W/3 ( coronavirus* OR "corona virus*" OR betacoronavirus* OR "beta coronavirus*" OR "beta corona virus*" OR sars OR "severe acute respiratory syndrome" ) ) OR 2019ncov OR covid OR covid19 OR covid2019 OR ncov OR sarscov2 ) ) OR ( TITLE ( "SARS-Cov-2 variant*" OR vui-202012-01 OR "VOC-202012/01" OR "B.1.1.7" OR b117 OR "B.1.351" OR b1351 OR "B.1.617" OR b1617 OR "P.1" OR p1 OR coronavirus* OR "corona virus*" OR betacoronavirus* OR "beta coronavirus*" OR "beta corona virus*" OR pneumonia* OR sars OR "severe acute respiratory syndrome" ) OR TITLE ( ( covid* OR sars* OR variant* ) AND ( alpha OR beta OR gamma OR delta OR "variant* of concern" OR "VOC" ) ) ) | 405, 697 |
| #3 | TITLE-ABS-KEY ( omicron  OR  "B.1.1.529"  OR  "VOC-21NOV-01"  OR  "B.1.1.529.1"  OR  "B.1.1.529.2"  OR  b11529* ) | 1,745 |
| #4 | #2 OR #3 | 407,402 |
| #5 | TITLE-ABS ( ( vaccin* OR immunis* OR immuniz* OR innoculat* OR inoculat* OR jab* OR shot* OR injection* OR nonimmunis* OR nonimmuniz* OR postvaccin* OR postimmunis* OR postimmuniz* OR reimmunis* OR reimmuniz* OR revaccinat* OR unimmunis* OR unimmuniz* OR ( ( one OR single OR two OR double OR first OR second OR partial* OR full ) W/2 ( dose* OR injection* OR injected OR injecting ) ) OR single-dose* OR double-dose* OR bnt162* OR comirnaty OR coronavac OR covishield OR tozinameran OR "mrna-1273*" OR chadox1* OR azd1222 OR "Ad26.COV2.S" OR "JNJ-78436735" OR novavax OR nvx-cov2373 OR ad5-ncov OR aapc OR "lentiviral minigene" OR vaxzevria ) ) | 2,043,780 |
| #6 | #4 AND #5 | 33,299 |
| #7 | TITLE-ABS ( antibody OR antibodies OR antibody-positiv* OR antibody-negativ* OR "complement activation" OR ( ( humoral OR humoural OR adaptive OR antibody-mediated ) W/3 ( respon* OR immun* ) ) OR immunity OR "immune system*" OR "immune marker*" OR ( ( immun* OR sero* ) W/3 respon* ) OR immunolog* OR immunomodulat* OR b-lymphocyte* OR "B-cell*" ) OR TITLE ( immun* OR immunoglobulin* OR iga OR igd OR ige OR igg OR igm OR sero* OR sera* ) | 2,474,238 |
| #8 | TITLE-ABS ( "correlat* of protection" OR "correlat* with protection" OR "immune correlat*" OR "correlat* of immunity" OR "correlat* of neutraliz*" OR "correlat* of neutralis*" OR "measur* of protection" OR "correlat* of infectivity" OR "protective correlat*" OR ( ( protective OR protection ) W/5 ( level* OR threshold* OR marker* OR limit* OR amount* OR mean* OR baseline OR quantif* ) ) OR ( ( correlat* OR marker* OR threshold* OR indicat* OR measur* ) W/5 ( immun* OR protect* ) ) OR ( ( antibody OR antibodies OR titer* OR titre* ) W/3 ( correlat* OR threshold* ) ) OR ( protect* W/5 ( immunity OR antibod* OR titer* OR titre* ) ) OR ( ( neutraliz* OR neutralis* ) W/2 ( level* OR amount* OR limit* OR mean* OR baseline OR quantif* OR titer* OR titre* ) ) OR ( predict* W/3 ( immun* OR protect* ) ) OR "protective immunity" OR "surrogate endpoint*" ) OR TITLE ( protection OR protective OR protected OR ( ( antibody OR antibodies ) W/2 ( status OR level* ) ) ) | 693,727 |
| #9 | #7 AND #8 | 205,010 |
| #10 | (#1 OR #6) AND #9 | 2,480 |
| #11 | #4 AND #9 | 4,810 |
| #12 | #10 OR #11 | 4,837 |
| #13 | LANGUAGE(English) OR LANGUAGE(French) AND PUBYEAR > 2020 | 3,609,628 |
| #14 | #12 AND #13 | 2,389 |
| #15 | #14 AND NOT INDEX (MEDLINE) | 866 |
